# Structural Brain Correlates of Cognitive Function in Schizophrenia: A Meta-Analysis

**DOI:** 10.1101/2021.04.16.21255551

**Authors:** Marianne Khalil, Philippine Hollander, Delphine Raucher-Chéné, Martin Lepage, Katie M. Lavigne

**Affiliations:** Department of Psychiatry, McGill University, Montreal, Quebec, Canada; Douglas Mental Health University Institute, McGill University, Montreal, Quebec, Canada; Faculty of Psychology and Neuroscience, Maastricht University, Maastricht, Netherlands; Department of Psychiatry, University Hospital of Reims, EPSM Marne, Reims, France; Cognition, Health, and Society Laboratory (EA 6291), University of Reims, Champagne-Ardenne, Reims, France; Montreal Neurological Institute, McGill University, Montreal, Quebec, Canada

**Keywords:** brain networks, volume, cognitive domains, speed of processing, attention, memory, executive function, social cognition

## Abstract

Schizophrenia is characterized by cognitive impairments and widespread structural brain alterations (e.g., decreased volume, thickness, surface area). Brain structure-cognition associations have been extensively studied in schizophrenia, typically involving individual cognitive domains or brain regions of interest. Findings in overlapping and diffuse brain regions may point to structural alterations in large-scale brain networks. We performed a systematic review and meta-analysis examining whether brain structure-cognition associations can be explained in terms of biologically meaningful brain networks. Of 7,621 screened articles, 88 were included in a series of meta-analyses assessing publication bias, heterogeneity, and study quality. Significant associations were found between overall brain structure and eight cognitive domains (speed of processing, attention/vigilance, working/verbal/visual memory, executive function, social cognition, and verbal fluency). Brain structure within functionally defined networks (default, dorsal/ventral attention, frontoparietal, limbic, somatosensory, visual) and external structures (amygdala, hippocampus and cerebellum) typically showed associations with conceptually related cognitive domains, with higher-level domains (e.g., executive function, social cognition) associated with more networks. These findings suggest brain structure-cognition associations in schizophrenia may follow network architecture.

## 1. Introduction

Schizophrenia (SZ) is a debilitating psychiatric disorder affecting approximately 20 million individuals worldwide (James et al., 2018). SZ is primarily characterized by positive (e.g., hallucinations, delusions) and negative (e.g., reduced motivation, flattened affect) symptoms; however, another core feature of SZ involves impaired cognitive abilities (Kahn & Keefe, 2013; Kahn et al., 2015). Cognitive impairments are observed in the majority of individuals diagnosed with SZ and are present across a wide range of domains (attention, memory, executive function; Heinrichs & Zakzanis, 1998). To characterize the cognitive domains impacted by SZ and encourage novel treatments, the Measurement and Treatment Research to Improve Cognition in Schizophrenia (MATRICS) categorized cognitive measures into seven neurocognitive domains: speed of processing, attention/vigilance, working memory, verbal learning and memory, visual learning and memory, reasoning and problem solving, and social cognition (Marder, 2006; Nuechterlein et al., 2008). Cognitive impairments emerge early in the illness, typically preceding and even outlasting many of the clinical symptoms since cognitive deficits are often not remediated with pharmacological treatment (Kitchen, Rofail, Heron, & Sacco, 2012; Lepage, Bodnar, & Bowie, 2014). In high-risk groups, cognitive deficits also predict the onset of psychosis – independently of other comorbidities – making it an important risk factor for schizophrenia (Seidman et al., 2016). Cognitive performance has also been found to be a strong predictor of poor functional outcomes in SZ compared to other clinical symptoms (Lepage et al., 2014). This persistence of cognitive impairments in SZ, from the prodrome to first episode to enduring SZ, may be reflected in structural brain alterations.

Structural imaging studies of patients with SZ have consistently shown a widespread decrease in cortical gray matter volume compared to healthy controls, especially in frontal and medial temporal areas (Shenton, Dickey, Frumin, & McCarley, 2001; van Erp et al., 2018) as well as in subcortical structures including the hippocampus and amygdala (van Erp et al., 2016). Research focusing on surface area and cortical thickness, which together constitute brain volume, has found widespread cortical thinning and reduced surface area across the cortical mantle in SZ (van Erp et al., 2018). Given the extensive brain and cognitive alterations in SZ, a large literature has examined whether structural brain abnormalities are associated with cognitive deficits in SZ, culminating in several reviews and targeted meta-analyses (Antoniades et al., 2018; Antonova, Sharma, Morris, & Kumari, 2004; Crespo-Facorro, Barbadillo, Pelayo-Terán, & Rodríguez-Sánchez, 2007; Fujiwara, Yassin, & Murai, 2015; Gur, Ragland, & Gur, 1997; Kelly et al., 2019).

Briefly, deficits in **speed of processing** have been negatively correlated with temporal lobe volume and ventricular size (Antonova et al., 2004). **Attention** was correlated with structural alterations (e.g., decreased volume and grey matter density, cortical thinning) within the frontal and temporal lobes (Kelly et al., 2019) as well as with widespread reduced volume of subcortical regions and the ventricles (Antonova et al., 2004). **Working memory** was negatively correlated with frontal and temporal lobes volume and cortical thickness (Antoniades et al., 2018; Crespo-Facorro et al., 2007) while **verbal learning and memory** was positively correlated with hippocampal volume loss (Antonova et al., 2004; Kelly et al., 2019). **Visual learning and memory** correlated with hippocampal volume loss and with frontal regions and primary visual occipital regions (Antonova et al., 2004; Kelly et al., 2019). **Reasoning** and problem solving, including higher-order **executive functions**, was also associated with a reduced frontal lobe volume, more specifically the dorsolateral prefrontal cortex (Antonova et al., 2004; Gur et al., 1997). Finally, **social cognition** encompassed several structural deficits in the amygdala, prefrontal cortex, temporal and parietal lobes including reduced grey-matter and white-matter volumes (Fujiwara et al., 2015).

Previous selective reviews highlight brain structure-cognition associations within overlapping brain regions in SZ, which may point to fundamental structural alterations within large-scale brain networks. It is becoming increasingly recognized that symptoms of SZ are likely subtended by disruptions of integrated networks of brain regions rather than to damage in specific areas (Glover et al., 2012; Pettersson-Yeo, Allen, Benetti, McGuire, & Mechelli, 2011). The extensive morphometric changes in SZ, combined with the overlap of several regions associated with complex cognitive functions, reinforces the need to shift from a regional to network perspective in structural neuroimaging studies (Bassett & Sporns, 2017; Park & Friston, 2013; Uddin, Yeo, & Spreng, 2019). In fact, the term “large-scale neurocognitive networks” has been found in the literature since the early nineties based on the understanding that the connections between brain areas impact their role in certain functions (Mesulam, 1990).

Connectivity-based brain parcellations are typically derived from functional or diffusion-weighted magnetic resonance imaging (MRI; Eickhoff, Yeo, & Genon, 2018). Although it is still unclear whether anatomically-derived networks (e.g., structural covariance; Evans, 2013) follow the same topography as functional networks, examination of structural alterations within functional networks (e.g., Uddin et al., 2019; Yeo et al., 2011) has been shown to help characterize structural abnormalities in SZ (Shafiei et al., 2020) as well as associations between clinical and anatomical dimensions of SZ (Kirschner et al., 2020). Indeed, significant associations have been noted between the default mode and visual networks with a domain comprising cognitive deficits and negative symptoms, two stable hallmarks of SZ (Kirschner et al., 2020), shedding light on potential structure-function associations in SZ and its relation to clinical dimensions of the disease. Yet, few studies have examined brain structure within functional networks in terms of distinct cognitive domains and it is unclear whether brain structure-cognition associations in SZ follow functional network architecture.

## Rationale and Objectives

Despite extensive research on morphological markers of cognitive deficits in SZ, a comprehensive quantitative synthesis of the literature has yet to be performed. Thus, we systematically reviewed the structural MRI literature to date on brain structure-cognition associations in SZ and performed a series of meta-analyses to synthesize the literature on associations between brain structure (e.g., volume, cortical thickness, surface area) and MATRICS-inspired cognitive domains (speed of processing, attention/vigilance, working memory, verbal and visual learning and memory, reasoning and executive function, social cognition, and verbal fluency) in SZ. We further examined whether brain structure-cognition associations could be characterized in terms of functional network topography using the seven networks defined by Yeo et al. (2011). We expected that these networks would provide a better understanding of the complex interrelations between cognitive domains and brain structure reported in the literature.

## 2. Methods

### 2.1. Protocol and Registration

This study protocol was pre-registered on PROSPERO: https://www.crd.york.ac.uk/prospero/

(CRD42020206152). The PRISMA guidelines for systematic reviews and meta-analyses were followed (Moher, Liberati, Tetzlaff, Altman, & Group, 2009). The PRISMA checklist for the current study is provided in the supplementary material.

### 2.2. Information Sources and Search Strategy

A comprehensive literature search was conducted using OVID (MEDLINE, PsycInfo, Health and Psychosocial Instruments, and EMBASE) and EBSCO (CINAHL) databases on August 3^rd^, 2020. The following keywords were used: (schizophreni* OR psychosis) AND (cogniti* OR attention OR vigilance OR speed of processing OR processing speed OR reasoning OR problem solving OR executive function OR verbal memory OR verbal learning OR visual memory OR visual learning OR working memory) AND (brain structure* OR morphometr* OR volume OR cortical thickness OR surface area). Reference lists of selected articles were also examined for additional studies. Evidence sources were limited to peer-reviewed articles and we excluded book chapters, conference abstracts, and poster presentations manually.

### 2.3. Eligibility Criteria

Retrieved articles were screened according to the following inclusion criteria: (a) peer-reviewed article; (b) reported on individuals with a diagnosis of schizophrenia-spectrum disorder (schizophrenia, schizoaffective, and schizophreniform disorders), and (c) included direct associations between cognition and brain structure using structural magnetic resonance imaging (T1-weighted or T2-weighted MRI). Both French and English peer-reviewed articles were included. Studies including only individuals presenting a diagnosis of a non-primary psychotic disorder (e.g., bipolar disorder with psychosis, delusional disorder, major depression with psychotic features, Alzheimer’s/dementia with psychosis), child-onset schizophrenia, first-episode psychosis, or who were at-risk for developing psychosis were excluded. Studies reporting combined results of schizophrenia-spectrum patients and healthy controls were excluded but those combining SZ and first-episode psychosis, or a non-primary psychotic disorder were included if the proportion of schizophrenia-spectrum patients was larger than half the sample. We also excluded studies that grouped several cognitive tests, morphometric measures, or multiple neuroimaging modalities in their assessment of structure-cognition associations as they could not be separated for our meta-analyses. Finally, authors of studies that did not report complete data were contacted to obtain additional information.

### 2.4. Selection of Sources of Evidence

Articles retrieved from OVID and EBSCO were combined in EndNote software and duplicates were removed automatically by comparing the Author, Year, Title, and Journal fields. Duplicates based on smaller combinations of these fields were then checked manually. The remaining unique articles were randomly ordered and assigned to one of three independent raters (MK, KML, VM) to assess titles and abstracts based on the article selection criteria. All three raters screened the same 200 articles (100 at the beginning and 100 at the end of the selection process) to assess inter- and intra- rater reliability (Belur, Tompson, Thornton, & Simon, 2018). The ratings of each set of 100 articles were compared between raters and statistical agreement between was computed with the Gwet agreement coefficient (AC_1_). AC_1_ was used to control for the Kappa paradox, when low agreement between raters is due to highly similar raters (Gwet, 2008; Wongpakaran, Wongpakaran, Wedding, & Gwet, 2013). Intra-rater reliability was assessed by computing the percentage of agreement between an article’s final consensus rating and the rater’s score (Belur et al., 2018). Discrepancies and questionable articles were resolved by a consensus between raters.

### 2.5. Data Extraction

Full texts of the selected articles were retrieved, and data were extracted using a pre-developed form by two independent reviewers (MK, PH). Quality control of full extractions was done for approximately 45% of articles by another independent reviewer (CL) and two additional independent reviewers (DRC, KL) reviewed the cognitive domain and network categorizations of all reported findings. The extracted information included demographic details (sample size, diagnosis, sex ratio, and age of patients and controls, if applicable), structural MRI metric (e.g., volume, cortical thickness, surface area), structural MRI brain coverage (e.g., vertex, regions of interest (ROIs), whole brain), cognitive measures (cognitive test, cognitive score), and the direct link between the structural and cognitive measures (analysis technique, the value of effect, statistic type and significance). Cognitive tests were categorized into eight domains including the seven MATRICS domains (speed of processing (SP), attention and vigilance (ATT), working memory (WM), verbal learning and memory (VM), visual learning and memory (VisM), reasoning and problem solving, and social cognition (SC); Nuechterlein et al., 2008). We also included verbal fluency (VF) as a separate domain, as its role within speed of processing has been debated (Nuechterlein et al., 2004). Furthermore, the MATRICS domain reasoning and problem solving was renamed reasoning and executive functions (R&EF) to include tasks measuring executive functioning that were not typically part of the MATRICS categorization, as done previously (Lavigne, Sauvé, Raucher-Chéné, & Lepage, 2020; Van Rheenen & Rossell, 2014). Finally, for tasks in which a higher score indicated lower performance (e.g., Trail-making Test, Wisconsin Card Sorting Task perseverative errors), the sign of the correlation was reversed.

### 2.6. Quality Assessment of Individual Studies

In order to assess the quality of the included studies in the meta-analysis, we developed a rating system inspired by the newly implemented scales of quality assessment of arterial spin labeling fMRI studies (Sukumar, Sabesan, Anazodo, & Palaniyappan, 2020) based on recommendations in the field (Alsop et al., 2015). Thus, we extracted five parameters (segmentation/atlas, multiple comparison correction, covariates, nonsignificant results, and scanner strength) each of which were rated on a scale of 0 to 1 and then the total of the 5 ratings was used to assess article quality. We extracted the **type of segmentation** or brain atlas/coordinates used (e.g., ROI-based coordinates, manual, semi-automatic, automatic; 0 = not reported, 1 = reported) which is necessary to anatomically-define brain areas and ensure replicability. We also extracted whether the article included **correction for multiple comparisons** (0 = not reported, 1 = reported) which is important to control for the overall significance level in the case of multiple tests (Poldrack, 2017). When extracting **covariates**, if age and sex were controlled for, the article was rated 1 and if either one was controlled for, the article was rated 0.5. Controlling for age and sex as covariates is essential due to their well-established association with brain structure and cognition beyond disease-related factors (Gennatas et al., 2017). We also indicated if studies reported **nonsignificant results** (0 = not reported, 1 = reported) since this impacts publication bias (Müller et al., 2018). Finally, we extracted the **scanner strength**. If the scanner strength was larger than 1.5T, the article was rated 1, otherwise, it was rated 0. All studies that had a total of 3 or more on the previous criteria were labeled high in quality while studies below 3 were labeled low in quality.

### 2.7. Meta-Analysis

We used the Comprehensive Meta-Analysis (CMA) software (version 2.2.021, Biostat, Englewood, NJ) to perform our meta-analyses. We chose Fisher’s Z to present our results since the primary outcome involved correlations between brain regions and cognitive domains. Fisher’s Z transformations were calculated for each study from the reported statistical effect (i.e., Pearson’s correlation, Spearman’s rho, t-tests or p-values) and sample size. Meta-analyses were conducted using a random effects model, which assumes that the true effect size of each of the studies varies and is not due to sampling variance only (Hall & Rosenthal, 2018). Additionally, the software weights studies based on their sample size which is important to control for effects driven by studies with a small number of patients that may be heterogenous.

We first performed eight overall meta-analyses of the correlations between each cognitive domain and all brain structure findings (refer to supplementary figure 1 for the meta-analyses schematic process). We assessed publication bias and heterogeneity of studies at this overall level. We then performed 10 subgroup analyses for each cognitive domain to further characterize these overall effects in terms of the seven Yeo et al. (2011) brain networks and three additional brain regions (i.e., hippocampus, amygdala, cerebellum) that emerged consistently in our systematic review. We controlled for multiple comparisons using the false discovery rate (FDR), a robust control for loss of power in studies with high throughput (Benjamini & Hochberg, 1995) for all of the ten subgroup analyses in each of the eight domains. We set the FDR to 0.05 which is the alpha used in the eight general meta-analyses.

**Figure 1:**
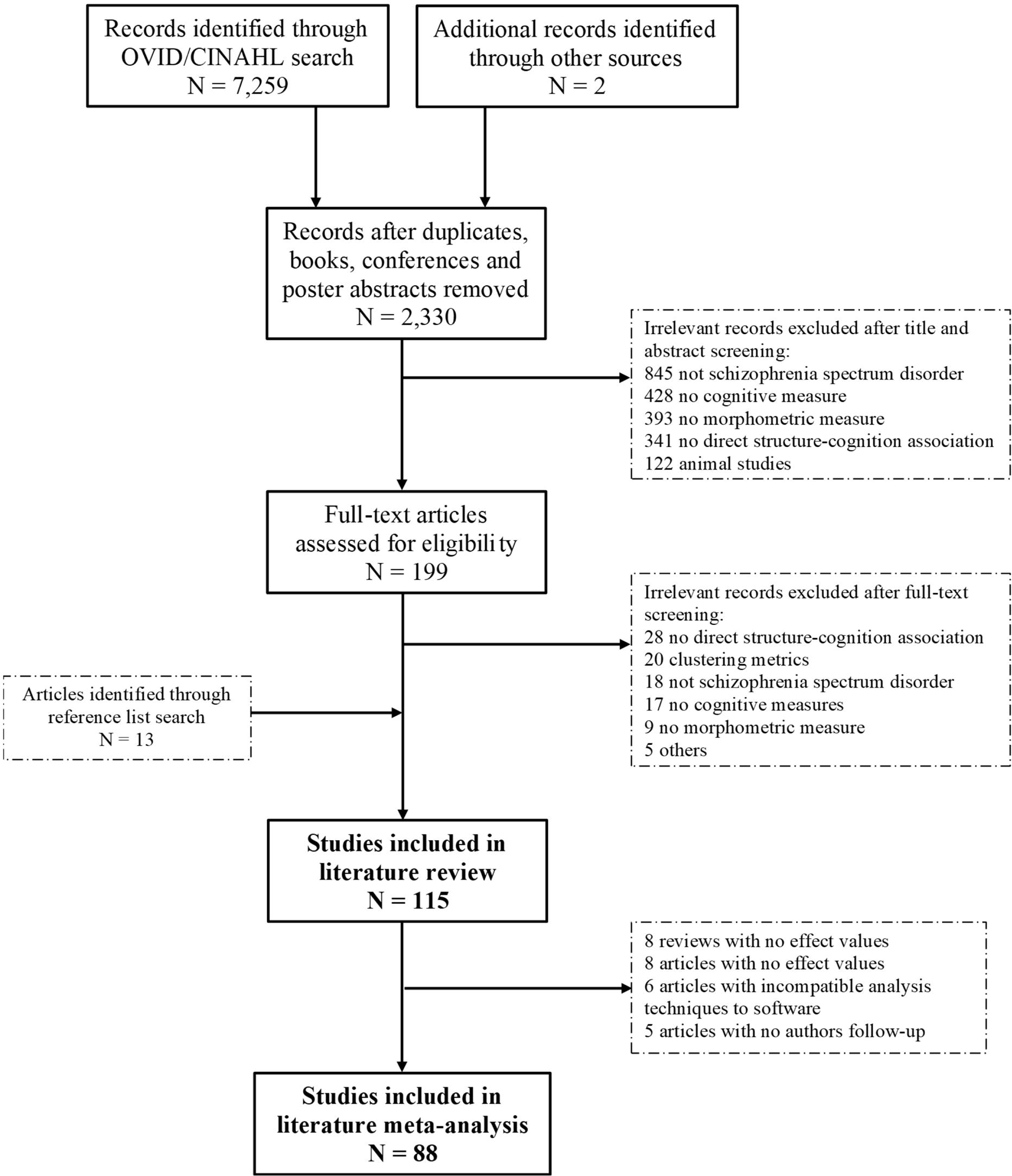
Flow chart of the article selection process following PRISMA guidelines.

#### 2.7.1. Whole Brain & Cognitive Domains

The overall meta-analyses on correlations between all reported structures and each of the cognitive domains included all the structural metrics (i.e., volume, cortical thickness, surface area). The aim of these overall analyses was to observe general trends and inform potential follow-up analyses.

##### 2.7.1.1. Risk of Bias Across Studies

We assessed risk of bias across studies for the eight overall meta-analyses since each included all the structure-cognition correlations for a given domain. Publication bias, defined as the impact that results have on the publication of a study (Easterbrook, Gopalan, Berlin, & Matthews, 1991), was assessed with two quantitative tests: Egger’s asymmetry test and the fail-safe N of Rosenthal. Given the large number of studies in each of the eight meta-analyses (see Results), qualitative assessments (e.g., visual inspection of funnel plots) were not used. Egger’s asymmetry test, a rank test, examines the significance of the correlation between effect sizes and their corresponding sampling variances and a significant result is likely evidence of publication bias (Egger, Smith, Schneider, & Minder, 1997). The fail-safe N of Rosenthal refers to the number of additional ‘negative’ studies that would be needed to increase the *p* value for a given meta-analysis to above 0.05 (Rosenthal, 1979). Usually, if the fail-safe N is larger than five times the number of studies included in the analysis plus ten then there is little evidence of publication bias (Fragkos, Tsagris, & Frangos, 2014).

We also assessed heterogeneity, the variation in outcomes between studies, with two quantitative tests: Cochran’s Q and the I2 Index. For Cochran’s Q, a p-value less than 0.1 indicates heterogeneity (Cochran, 1950). Cochran’s Q is a commonly used method but has low power when the number of studies is small and excessive power when the number of studies becomes large (Pereira, Patsopoulos, Salanti, & Ioannidis, 2010). This posed a problem for our meta-analysis due to the variation in the number of studies for each domain. Thus, we supplemented our heterogeneity assessment with the I^2^ statistic, which is not impacted by the number of studies in the meta-analysis (Pereira et al., 2010). The I2 statistic describes the percentage of variation across studies that is due to heterogeneity rather than chance with general effect size cut-offs of 25% (small), 50% (medium) and 75% (large; Higgins & Thompson, 2002).

##### 2.7.1.2. Quality Assessment

Following the overall meta-analyses, we also performed two subgroup analyses for each of the eight domains by filtering for the quality of the studies (low/high quality). We then performed, using the CMA software, a z-test comparing the mean difference between the two subgroups to determine whether if there was a significant difference between low- and high-quality studies.

### 2.8. Networks Subgroup Analyses

We performed a series of subgroup analyses to study the specific associations between brain networks and cognition. To do so, we first classified brain regions from studies using the Desikan Killiany Tourville (DKT) atlas (Klein & Tourville, 2012) parcellation method, as done in Makowski et al. (2020), into the seven Yeo et al. (2011) networks: default mode network (DMN), dorsal attention network (DAN), frontoparietal control network (FPN), limbic network (LIM), somatic network (SOM), ventral attention network (VAN) and visual network (VIS). Afterwards, using MRIcron, we overlaid the Yeo atlas of the seven networks with the structural atlases reported by other articles (MNI305, Broadman, AAL) to classify regions. If coordinates were provided, we were able to attribute a finding directly to a specific network. Consensus between authors was reached for the questionable brain regions classification. Studies which reported findings on larger brain areas (i.e., frontal lobe, temporal lobe) or unclassified structures (e.g., subcortical structures, white matter, ventricles, cerebellum, corpus callosum), were excluded from the network subgroup analyses. Given the importance of some of these regions in cognition and their prevalence in the identified literature (e.g., amygdala, hippocampus), these were assessed separately (see below) rather than excluded entirely. See Table S1 for a detailed description of the brain structures, and if applicable, their attributed network.

In CMA, we used the name of the network as a subgroup variable, laterality of the region as a differentiating variable and the name of the cognitive test as an outcome to perform seven sub-group analyses for each of the eight domains. To investigate the precise regions of the networks contributing to the correlation with cognition, we also ran more precise subgroup analyses with the name of the brain structures as a subgroup variable filtering for the brain structures with a significant network-domain association based on the previous subgroup analyses.

### 2.9. Other Structures Analyses

Specific structures (amygdala, hippocampus, overall cerebellum) were investigated with separate subgroup analyses as they have shown structural changes and cognitive correlations in the literature (Koshiyama et al., 2018). Other potentially relevant structures (i.e., thalamus, nucleus accumbens) were not included due to limited studies in the literature review. Finally, although the cerebellum has a common network parcellation to the Yeo et al. (2011) cerebral atlas (Buckner, Krienen, Castellanos, Diaz, & Yeo, 2011), studies generally reported overall correlations with the cerebellum, and we could not include it in the network analyses. Thus, we investigated these regions’ correlations with cognition in three subgroup analyses for each of the eight domains.

## 3. Results

### 3.1. Study Selection

The flowchart for article selection is displayed in Figure 1. The database search identified 7,259 articles and two additional articles were identified by co-authors. After the removal of 4,929 duplicates, books and conference abstracts, the remaining 2,330 titles and abstracts were randomly assigned to one of three raters. There were 199 articles flagged for full-text screening with the addition of 13 articles from reference lists. Although we included 115 articles in our systematic review (see Table S2), 27 were not included in the meta-analysis as they either represented review articles, did not report relevant values of effect and authors could not be reached, or used analysis techniques that were incompatible with our software (e.g., multivariate analysis). Thus, 88 original research articles were included in our meta-analyses. See Table S2 for a summary of studies included in the review and meta-analyses.

### 3.2. Raters’ Reliability

The results of the Gwet’s agreement coefficient (AC_1_) for the inter-rater reliability tests were high for both timepoints: IRR1 = 0.87 (SE = 0.03, 95% CI = [0.80 - 0.93], *p* < 0.001) and IRR2 = 0.88 (SE = 0.03, 95% CI = [0.82 - 0.95], *p* < 0.001). Additionally, the percentage of agreement between IRR1 (Rater 1: 92%; Rater 2: 89%; Rater 3: 92%) and IRR2 was high (Rater 1: 95%; Rater 2: 91%; Rater 3: 95%) and increased for all raters.

### 3.3. Whole Brain & Cognitive Domains Meta-Analyses

We performed a meta-analysis for each of the eight cognitive domains to determine if they were correlated with overall brain structures, including all structural metrics. We found a significant summary Fisher’s *z* for all domains, ranging from 0.087 (VF) to 0.669 (SC; see Figure 2; Figures S2-S9 for individual forest plots). To further explore the exceptionally strong association observed with SC relative to the other domains, we divided this domain into two sub-categories, emotion processing and theory of mind, based on the type of task reported in the article. Subgroup analyses, followed by a z-test to compare the difference of means, showed significant correlations between brain structures and both emotional processing (Fisher’s *z* = 0.778, 95% CI [0.486, 1.069]) and theory of mind (Fisher’s *z* = 0.623, 95% CI [0.432, 0.814]), with no significant differences between the two sub-domains (*p* > 0.05; see Figure 2; Figures S10-S11 for individual plots). Finally, when performing the overall meta-analyses, we found that for all cognitive domains, volume was the most reported metric for brain structures ranging between 78% (SP) to 95% (VM) of all included articles in the domains.

**Figure 2:**
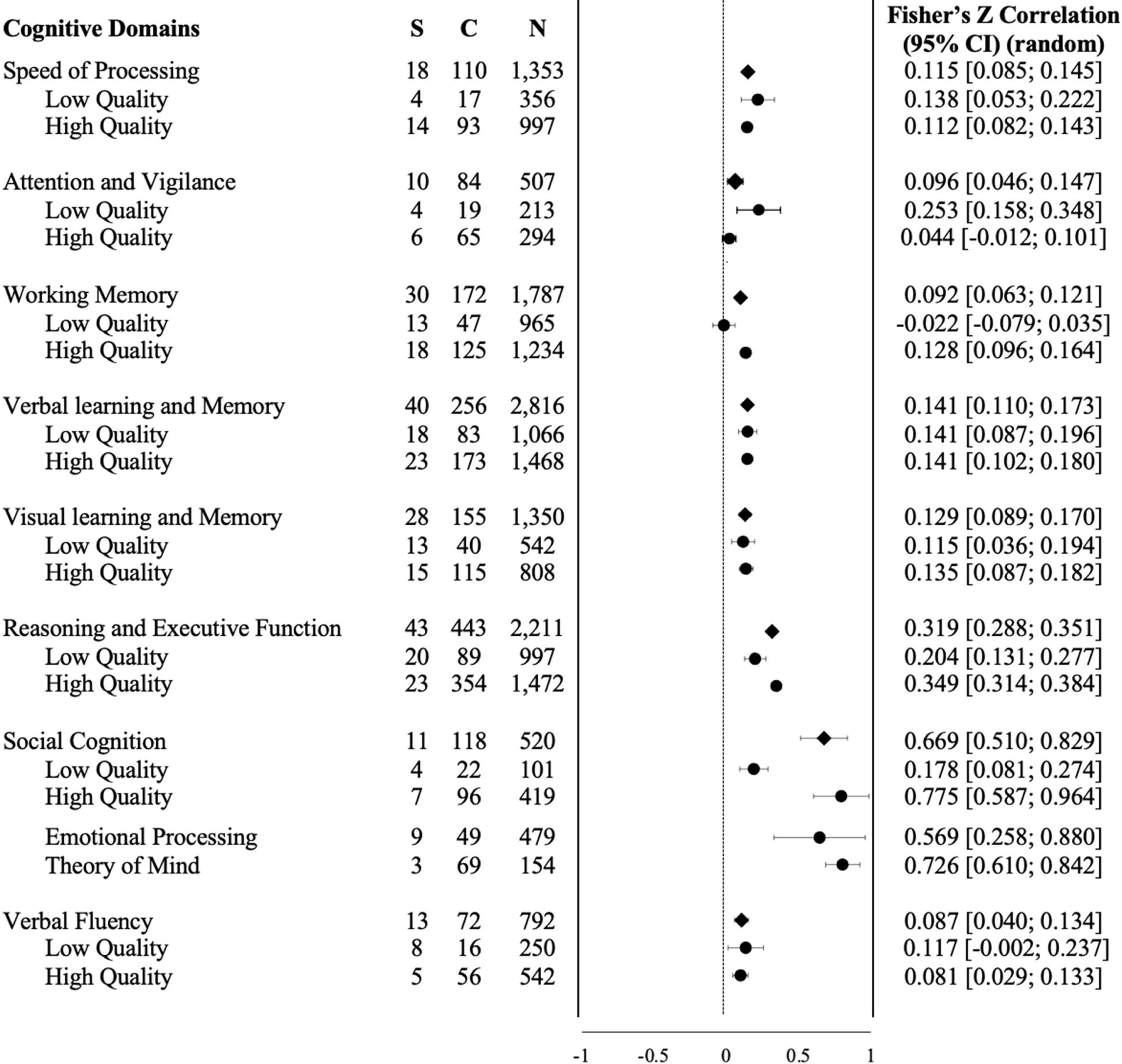
Overall correlation between each cognitive domain and overall brain structures including all structural metrics. Subgroup analyses between low- and high- quality studies are also presented for each cognitive domain. The number of studies (S), correlations (C) and patients (N) are indicated for each of correlations.

#### 3.3.1. Risk of Bias

We assessed publication bias with Egger’s Asymmetry and Fail-Safe N of Rosenthal tests (see Table 1). Egger’s Asymmetry test indicated potential publication bias (*p* < 0.05) for R&EF, SC and VF; however, no evidence of bias was observed with the fail-safe N of Rosenthal, which was above the cut-off (five times the number of studies included in the analysis plus ten) for all cognitive domains. Furthermore, the Cochran’s Q test for all domains had a p-value below 0.1 indicating heterogeneity between studies (Potvin, 2020). Similarly, the I^2^ index signalled moderate to high heterogeneity between studies for all cognitive domains (I^2^ range = 52.97 for SP to 96.80 for SC), except VF, which showed low-to-moderate heterogeneity (I^2^ = 46.00).

**Table 1:**
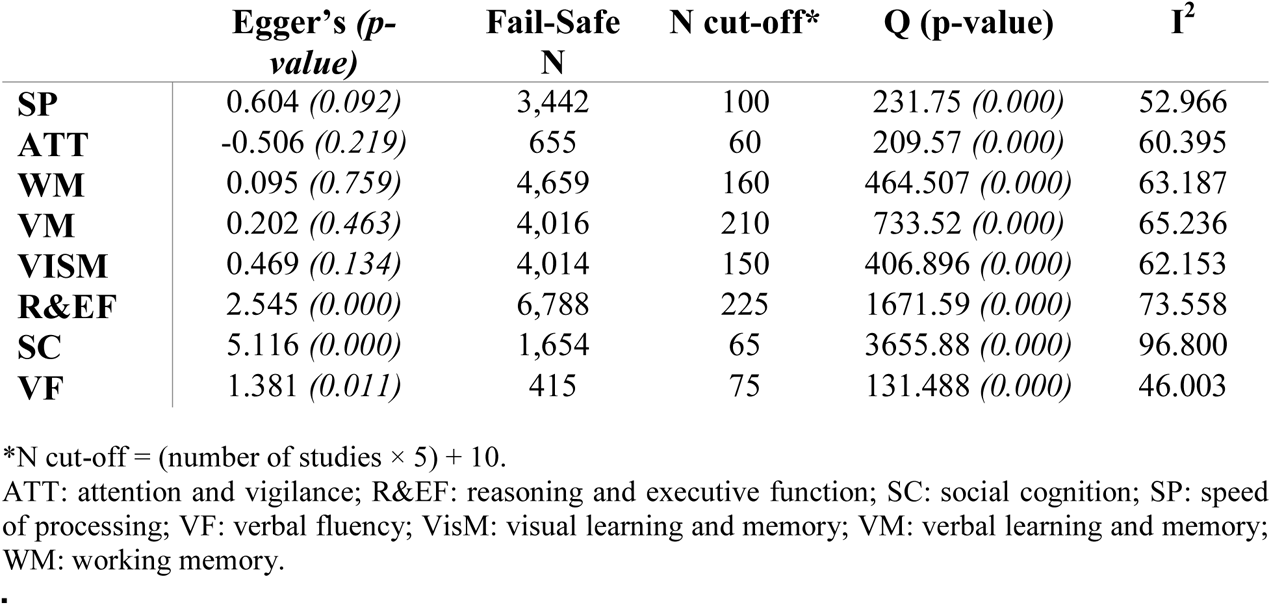
Results of publication bias tests and heterogeneity for each cognitive domain

#### 3.3.2. Quality Assessment

The rating of the included studies is displayed in the supplementary material (Table S3). Subgroup analyses followed by a test of the differences in means showed that, for ATT, low-quality studies were significantly more correlated with brain structures than those of high-quality (Z_Diff_ = -4.55, *p* < 0.05). For R&EF (Z_Diff_ = 4.62, *p* < 0.05), SC (Z_Diff_ = 2.71, *p* < 0.05) and WM (Z_Diff_ = 4.74, *p* < 0.05) high-quality studies showed a significantly larger correlation with brain structures than those of low quality. SP (Z_Diff_ = -0.581, *p* > 0.70), VF (Z_Diff_ = -0.443, *p* > 0.05), VisM (Z_Diff_ = 0.365, *p* > 0.05) and VM (Z_Diff_ = 0, *p* > 0.05) did not show evidence of significant difference between the correlations of low- and high-quality studies. The overall forest plot (Figure 2) provides the Fisher’s z correlation of each quality subgroup for comparison with the overall results of the overall meta-analyses for each domain.

### 3.4. Network Subgroup Analyses

Using brain network topography (Figure 3), we found significant (connections displayed in color) and non-significant (connections displayed in greyscale) correlations between specific networks and cognitive domains in SZ. Figure 3 also highlights associations not investigated in the literature (no connections). Additionally, we were able to observe which brain structures within each network contributed to a significant correlation with a given cognitive domain (Figure 4). The effect values of all the correlations between networks and domains as well as the brain structures and the domains are listed in Table S4 and visualized in the circle plots in Figure S12.

**Figure 3:**
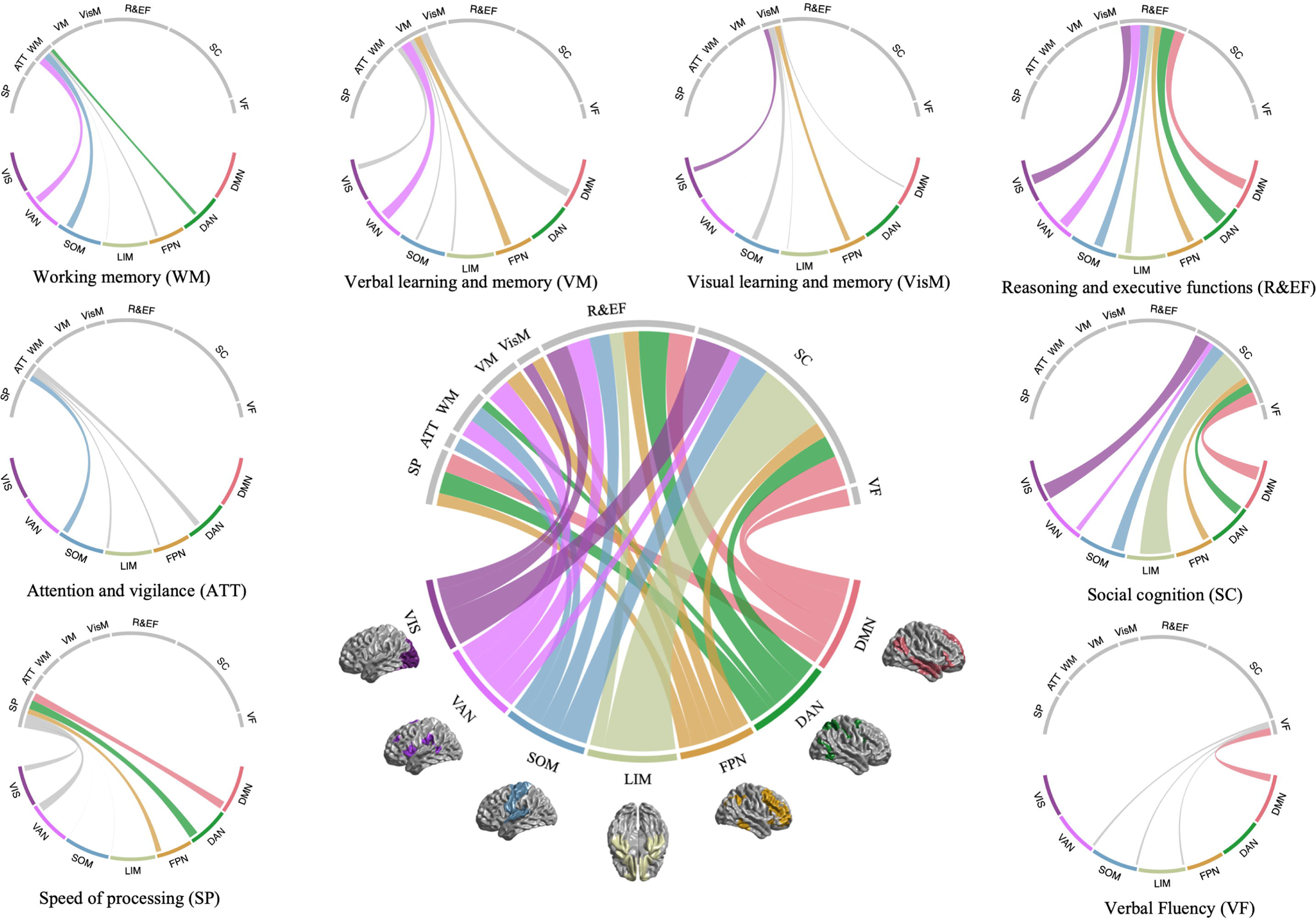
Lower middle panel: circle plot of the significant FDR-corrected correlations between the seven brain networks and the eight cognitive domains. The thickness of the link is proportional to the correlation strength (Table S4). Surrounding panels (bottom left to bottom right): significant (in colour) and non-significant (grey) summary correlations for each cognitive domain. Unexplored associations are represented as absent links between a domain and network. DMN: default-model network, DAN: dorsal attention network, FPN: frontoparietal network, LIM: limbic network, SOM: somatosensory network, VAN: ventral attention network, VIS: visual network.

**Figure 4:**
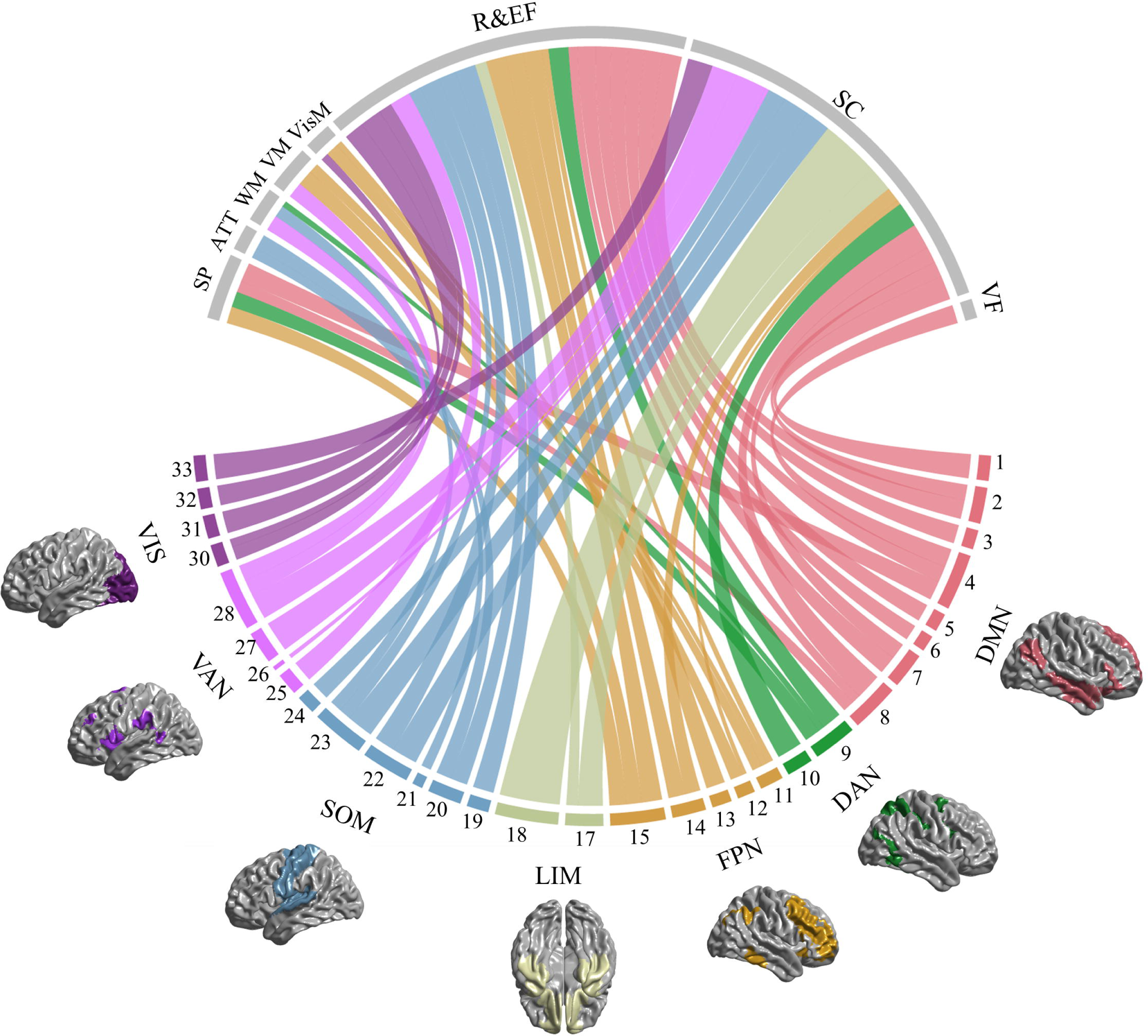
Circle plot of the FDR significant correlations between the brain regions categorized in the seven brain networks and the eight cognitive domains. The thickness of the link is proportional to the correlation strength. The colors of the brain regions correspond to the respective network. The legend and the values of the effect are in Table S4. Non-significant correlations are shown in Figure S12. ATT: attention and vigilance, DMN: default-model network, DAN: dorsal attention network, FPN: frontoparietal network, LIM: limbic network, R&EF: reasoning and executive function, SC: social cognition, SOM: somatosensory network, SP: speed of processing, VAN: ventral attention network, VF: verbal fluency, VIS: visual network, VisM: visual memory, VM: verbal memory, WM: working memory. 1: angular gyrus; 2: anterior cingulate gyrus; 3: IFG, pars orbitalis; 4: inferior parietal lobule; 5: inferior temporal gyrus; 6: lateral aspect; 7: middle temporal gyrus; 8: posterior cingulate gyrus; 9: precuneus; 10: temporal pole; 11: IFG, pars opercularis; 12: superior parietal lobule; 13: dorsolateral prefrontal cortex; 14: inferior frontal gyrus; 15: IFG, pars triangularis; 17: middle frontal gyrus; 18: entorhinal cortex; 19: gyrus rectus; 20: orbitofrontal cortex; 21: parahippocampal cortex; 22: claustrum; 23: Heschl’s gyrus; 24: planum temporale; 25: precentral gyrus; 26: superior temporal gyrus; 27: supplementary motor area; 28: BANKSTS; 30: middle cingulate gyrus; 31: superior frontal gyrus; 32: temporoparietal junction; 33: calcarine cortex

**SP** was significantly associated with three networks DMN (Fisher’s *z* = 0.508, 95% CI: [0.280; 0.737]), DAN (*z* = 0.576, [0.212; 0.940]) and FPN (*z* = 0.337, [0.106; 0.568]). For the DMN, the inferior temporal gyrus (*z* = 0.704, [0.346, 1.062]) was the strongest brain structure correlated with the domain. For the DAN, it was the inferior frontal gyrus (IFG), pars opercularis (*z* = 0.576, [0.212, 0.940]) and for the FPN, it was the middle cingulate gyrus (*z* = 1.131, [0.658, 1.605]).

The **ATT** cognitive domain was found to be significantly correlated only with the SOM network (*z* = 0.358, [0.205; 0.511]) due to Heschl’s gyrus which had the strongest significant correlation out of the other four for this domain (*z* = 0.551, [0.351, 0.751]).

**WM** was associated significantly with three networks DAN (*z* = 0.221, [0.048; 0.394]), SOM (*z* = 0.436, [0.276; 0.595]) and VAN (*z* = 0.443, [0.183; 0.702]). These network correlations were driven by the IFG pars opercularis (*z* = 0.221, [0.048, 0.394]) for the DAN, by the superior temporal gyrus (*z* = 0.436, [0.276, 0.595]) for the SOM network and by the superior frontal gyrus (*z* = 0.572, [0.202, 0.943]) for the VAN.

**VM** was correlated significantly with FPN (*z* = 0.440, [0.290; 0.590]) and VAN (*z* = 0.622, [0.253; 0.992]). It also correlated most strongly with the middle (*z* = 0.576, [0.182, 0.970]) and the superior (*z* = 0.576, [0.310, 0.843]) frontal gyri categorized in the FPN and VAN, respectively.

**VisM** was significantly associated with FPN (*z* = 0.322, [0.202; 0.443]) and VIS (*z* = 0.291, [0.072; 0.509]) networks. These network correlations were driven by inferior frontal gyrus (*z* = 0.465, [0.286, 0.645]) for the FPN and by fusiform gyrus (*z* = 0.291, [0.072, 0.509]) for the VIS.

**R&EF** was significantly associated with all seven brain networks. These network correlations were driven by the anterior cingulate gyrus (*z* = 0.694, [0.556, 0.832]) for the DMN (*z* = 0.620, [0.542; 0.699]), by the IFG pars opercularis (*z* = 0.729, [0.593, 0.864]) for the DAN (*z* = 0.729, [0.593; 0.864]), by the middle frontal gyrus (*z* = 0.741, [0.517, 0.966]) for the FPN (*z* = 0.411, [0.298; 0.523]), by the orbitofrontal cortex (*z* = 0.423, [0.269, 0.577]) for the LIM network (*z* = 0.365, [0.216; 0.513]), by the claustrum (*z* = 0.769, [0.351, 1.186]) for the SOM network (*z* = 0.537, [0.384; 0.690]), by the BANKSTS (*z* = 0.775, [0.569, 0.982]) for the VAN (*z* = 0.604, [0.511; 0.697]), and by the inferior occipital gyrus (*z* = 0.803, [0.385, 1.221]) for the VIS network (*z* = 0.592, [0.448; 0.734]).

The **SC** domain was also significantly associated with all the brain networks. These network correlations were driven by the posterior cingulate gyrus (*z* = 1.722, [0.134, 3.309]) for the DMN (*z* = 0.804, [0.356; 1.253]), the superior parietal lobule (*z* = 0.929, [0.467, 1.391]) for the DAN (*z* = 0.929, [0.467; 1.391]), the IFG pars triangularis (*z* = 0.640, [0.056, 1.224]) for the FPN (*z* = 0.411, [0.237; 0.585]), the parahippocampal gyrus (*z* = 2.215, [0.585, 3.845]) for the LIM network (*z* = 1.892, [0.547; 3.236]), the superior temporal gyrus (*z* = 0.963, [0.637, 1.290]) for the SOM network (*z* = 0.784, [0.545; 1.024]), the middle cingulate gyrus (*z* = 1.131, [0.658, 1.605]) for the VAN (*z* = 0.315, [0.214; 0.415]), and the middle occipital gyrus (*z* = 0.914, [0.648, 1.181]) for the VIS network (*z* = 0.914, [0.648; 1.181]).

Finally, the **VF** domain was only correlated significantly with DMN (*z* = 0.447, [0.088; 0.806]) mostly driven by the strong association with the inferior temporal gyrus (*z* = 0.678, [0.172, 1.184]).

### 3.5. Structures Subgroup Analyses

Selected structures commonly emerging in the literature review included the amygdala, the cerebellum and the hippocampus (Figure 5). **SP** was significantly associated with the hippocampus (*z* = 0.130, [0.080, 0.180]) and the amygdala (*z* = 0.204, [0.141, 0.267]) while **ATT** was only significantly associated with the cerebellum (*z* = 0.109, [0.030, 0.188]). **WM** was found to be significantly correlated with all three structures (amygdala: *z* = 0.136, [0.081, 0.191]; cerebellum: *z* = 0.800, [0.141, 1.078]; hippocampus: *z* = 0.159, [0.115, 0.202]). **VM** was found to be associated significantly with the hippocampus (*z* = 0.234, [0.181, 0.287]) and **VisM** was found to be correlated significantly with the cerebellum (*z* = 1.042, [0.816, 1.268]) and the hippocampus (*z* = 0.141, [0.063, 0.219]). The **R&EF** domain was significantly correlated with the hippocampus (*z* = 0.190, [0.111, 0.268]) and the amygdala (*z* = 0.169, [0.075, 0.263]). **SC** was investigated in relation to the hippocampus (*z* = 1.263, [0.801, 1.725]) and the amygdala (*z* = 0.596, [0.351, 0.841]) and found significant for both. Finally, **VF** was significantly associated with the cerebellum (*z* = 0.201, [0.135, 0.267]).

**Figure 5:**
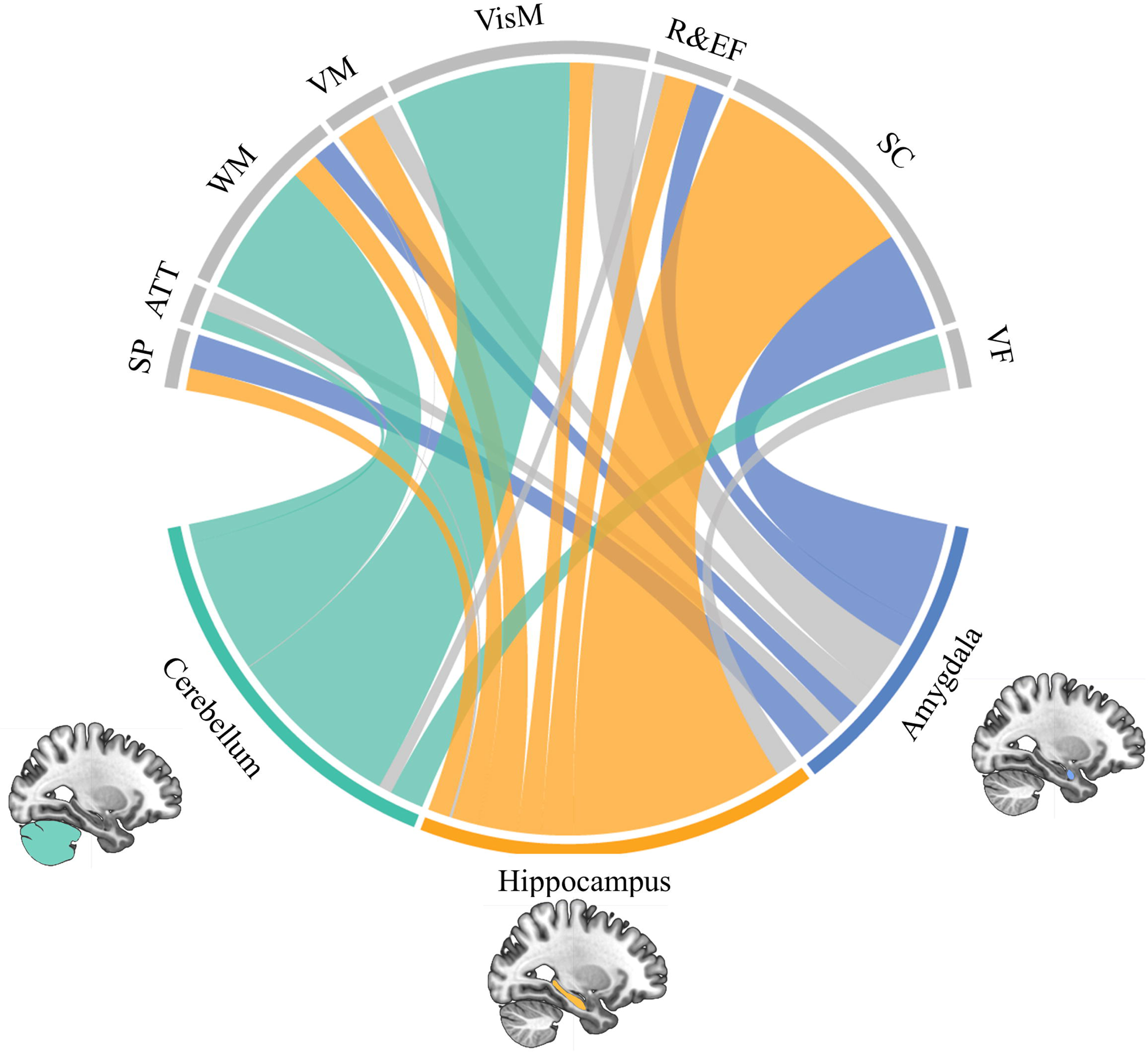
Circle plot of the correlations between three structures not categorized in networks and the eight cognitive domains. The thickness of the link is proportional to the correlation strength (Table S6). The grey links are nonsignificant with FDR. ATT: attention and vigilance, R&EF: reasoning and executive function, SC: social cognition, SP: speed of processing, VF: verbal fluency, VisM: visual memory, VM: verbal memory, WM: working memory.

## 4. Discussion

The aim of this study was to comprehensively synthesize the literature to date on associations between brain structure and cognition in SZ as well as examine whether these could be better understood from a network perspective. We identified 115 articles investigating brain-cognition associations in SZ, 88 of which were included in our meta-analyses. Overall, we found that all cognitive domains were generally associated with brain structure in schizophrenia. Brain network subgroup analyses provided deeper insight into these associations from a structure-function perspective and point to potential hub regions associated with distinct cognitive functions. Notably, we observed that higher-level cognitive processes (e.g., reasoning and executive function, social cognition) tend to be associated with a greater number of networks compared to other cognitive domains. We also identified specific network-cognition associations lacking in the literature that would be expected based on the network’s functional characteristics. Thus, leveraging functional network topography allowed us to better characterize brain structure-cognition associations in schizophrenia, suggesting that brain structural correlates of cognition may follow network architecture.

### 4.1. Whole Brain & Cognitive Domains Meta-Analyses

The overall set of meta-analyses yielded significant correlations between brain structure and all eight cognitive domains. The strongest overall brain-cognition associations were with SP, VM, VisM, R&EF and SC. These domains are also those with the most associated studies, which might reflect research trends in the field or more knowledge on structural regions associated with them than other domains. We detected publication bias for three domains (R&EF, SC and VF). We thus acknowledge that this partiality in the literature might impact our results for these domains. Additionally, heterogeneity was present for all eight domains, which was expected due to the variability of methodologies across studies and our inclusion of multiple metrics and brain regions. Interestingly, VF was the only domain to show low heterogeneity, which might be partially due to this domain being measured by only two cognitive tests with little variation in scoring (letter and category fluency). The use of the random effects model in all meta-analyses to estimate the correlation between brain structure and cognitive domains was employed to remediate this and other potential sources of variation between studies (Lipsey & Wilson, 2001).

We also assessed the quality of the studies included in each of the eight meta-analyses and expected that the results in high-quality studies would differ significantly from those of low-quality. This was the case for ATT, R&EF, SC and WM, suggesting that methodological differences relating to brain segmentation/atlas, multiple comparisons, covariates, nonsignificant results, and scanner strength have a strong impact on the overall correlation with brain structure for at least these domains. Surprisingly, larger effects were observed for low- relative to high-quality studies for ATT, which warrants further investigation. The remaining domains (SP, VM, VisM, and VF) did not show a significant difference between low- and high-quality studies, which could indicate that brain structure associations with these domains are less affected by these criteria, that there is a certain homogeneity in the way these domains have been assessed to date, or perhaps that the strength of the effects eclipse the confounding impact of the quality criteria. The subsequent network subgroup analyses aimed to address the additive effects of the overall analysis, which could hide more subtle correlations, and provide a targeted view of brain structure-cognition associations in schizophrenia.

### 4.2. Cognition and Networks

Leveraging functional brain network architecture provided new insights concerning network-cognition associations in schizophrenia, revealing significant correlations between all cognitive domains and at least one brain network. Follow-up analyses allowed us to pinpoint within-network regions contributing most strongly to these network-cognition associations. Overall, we observed that associations between brain structure and cognition can be understood in terms of brain network architecture and key brain regions, which are interpreted by cognitive domain below.

#### 4.2.1. Speed of Processing

This domain was significantly associated with three networks: the DMN correlation was driven by inferior temporal gyrus, the DAN by IFG, pars opercularis, and the FPN by middle cingulate gyrus. These results are consistent with recent functional and effective connectivity findings indicating that digit symbol coding (the task most commonly employed within this domain) involves interactions between frontoparietal and inferior temporal/frontal systems involved in goal- and stimulus-directed behaviour (Silva et al., 2019). While the emergence of the DMN in the current study was unexpected, this network’s effect was driven primarily by the inferior temporal gyrus, which has been designated a number-form area comprising a "symbolic number processing network" along with several other regions observed herein (e.g., inferior frontal gyrus; Yeo, Wilkey, & Price, 2017). The current findings suggest that these associations with speed of processing in healthy individuals may extend to SZ and that structural alterations within these regions contribute to speed of processing deficits in SZ. Concerning other structures, SP was significantly correlated with both the amygdala and the hippocampus involved in learning and memory (Andersen, Morris, Amaral, Bliss, & O’Keefe, 2006). This is in line with behavioral research indicating links between memory and processing speed in schizophrenia as well as the potential use of memory-related strategies to perform SP tasks (Brébion, Amador, Smith, & Gorman, 1998; Knowles et al., 2015). Finally, this domain did not show significant differences between low- and high- quality studies in the overall meta-analysis which could be due to high heterogeneity in neuroimaging methodology between studies. Previous meta-analyses pointed to the high variability in effect sizes of studies with coding tasks compared to other tasks of SP (Knowles, David, & Reichenberg, 2010).

#### 4.2.2. Attention and Vigilance

ATT was only significantly correlated with the SOM network, driven by Heschl’s gyrus. The planum temporale, close to the temporo-parietal junction and part of the SOM network, was also significantly correlated with ATT supporting previous reviews’ findings (Antonova et al., 2004; Lesh, Niendam, Minzenberg, & Carter, 2011). This is likely due to several studies using auditory continuous performance tests to assess attention. We also expected DAN and VAN to significantly correlate with this domain, but surprisingly, associations between ATT and regions within these attentional networks were rarely assessed in the literature. On one hand, the DAN showed a non-significant correlation, possibly due to the fact that only two associations with one specific brain region (IFG pars opercularis) were examined in the literature. On the other hand, studies did not report any correlations with brain regions categorized within the VAN. Thus, more research is necessary to determine whether structural associations with attention and vigilance in schizophrenia are limited to primary sensory regions or are also reflected in attention-related networks.

We also observed a significant correlation between this domain and the cerebellum, recently known for its integrative cognitive functions (Schmahmann, 2019). Indeed, theories of cerebellar dysfunction in schizophrenia point to impaired coordinated integration of stimuli, also known as cognitive dysmetria (Andreasen, Paradiso, & O’Leary, 1998; Schmahmann, 2019). Finally, this domain contained the lowest number of studies and a high heterogeneity which points to an important gap in the literature and may indicate high variability in the tasks used to assess attention, which has been previously noted to lead to opposing results (Hoonakker, Doignon-Camus, & Bonnefond, 2017).

#### 4.2.3. Working Memory

This domain was significantly associated with three networks: the DAN driven by the IFG opercularis, the SOM driven by the superior temporal gyrus, and the VAN by the superior frontal gyrus. This emergence of attentional networks (DAN, VAN) is not unexpected given that most tasks assessing WM require goal- and stimulus-directed processing (Moreau & Champagne-Lavau, 2014; Nuechterlein et al., 2004). Contribution of the superior temporal gyrus, part of the SOM, may reflect the auditory nature of several WM tasks (e.g., traditional digit span). No other network survived FDR correction although FPN and LIM were both found to have high numbers of correlations in the literature (21 and 14 respectively). Functional studies have pointed to an association between the FPN, more specifically the dorsolateral prefrontal cortex, and the DMN with WM in SZ (Godwin, Ji, Kandala, & Mamah, 2017; Kelly et al., 2019). The lack of association with the FPN is striking, particularly given the importance of the dorsolateral prefrontal cortex in WM via region of interest studies. Notably, the superior frontal gyrus, which did emerge as driving the association between the DAN and WM, overlaps anatomically with the dorsolateral prefrontal cortex. Other possibilities include the DLPFC association with WM being primarily of a functional nature or being obscured due to WM task variability. The lack of structural studies examining brain regions categorized in the DMN (none found in our review) indicates directions for future research. Another expected correlation of WM that we found was with the cerebellum, thought to be one of the important regions of working memory (Vandervert, Schimpf, & Liu, 2007). Finally, due to its function in learning and memory (Andersen et al., 2006), it was also expected that the hippocampus would be significantly correlated with WM which was supported by our findings.

#### 4.2.4. Verbal Learning and Memory

VM was significantly associated with two networks, the FPN driven by the dorsolateral and middle frontal gyrus, and the VAN driven by the superior frontal gyrus. These findings support previous evidence indicating an association between VM and thinning of the frontal cortex in SZ patients (Antonova et al., 2004; Guimond, Chakravarty, Bergeron-Gagnon, Patel, & Lepage, 2016) as well as a reduced volume of prefrontal regions (Antonova et al., 2004). As expected, due to its importance in memory, the hippocampus was also significantly associated with VM as shown previously (Antoniades et al., 2018; Guimond et al., 2016). However, the amygdala, usually involved in the emotional tagging of memories, did not significantly correlate with VM. This was not due to a lack of research investigating the association but is consistent with the general absence of emotional content in the tasks typically used to assess VM. Finally, the LIM, which includes parahippocampus and entorhinal cortex (input and output regions for the hippocampus, respectively; Foster, Kennedy, Hoagey, and Rodrigue (2019)) was correlated with VM but did not retain significance following FDR correction. This may point to subnetworks or modules of LIM preferentially related to VM.

#### 4.2.5. Visual Learning and Memory

VisM was significantly associated with the FPN, due to a strong correlation with the dorsolateral prefrontal cortex, and the VIS network, driven by a significant association with the fusiform gyrus. We expected the VisM-VIS association since this network is specific to visual processing and has been observed previously (Antonova et al., 2004; Kelly et al., 2019). The particular association between the fusiform gyrus, known for its involvement in facial recognition (Weiner & Zilles, 2016), and VisM may be due to studies assessing VisM with facial recognition tasks. Previous reviews also pointed to the prefrontal cortex as a processing and integration area for visual information (Antonova et al., 2004; Kelly et al., 2019), which is supported by the current findings. Four other networks were investigated in this domain but were nonsignificant. Although this effect can be explained by a small number of correlations for three of the networks (DMN, SOM, VAN), the LIM was assessed over it is not the case for the LIM network which has 17 associations in the literature. Thus, it could be that the tasks assessing VisM have no emotional or reward component that would impact memory encoding or retrieval. This could also be the case for the nonsignificant association with the amygdala. In contrast, the cerebellum, implicated in facial recognition (Andreasen & Pierson, 2008), and the hippocampus, involved in memory, was significantly associated with VisM.

#### 4.2.6. Reasoning and Executive Function

This domain, which is considered a higher-level domain encompassing several cognitive processes (e.g., SP, ATT; Nuechterlein et al., 2004), was significantly associated with all seven networks. Some of the strongest contributing regions to these findings included anterior cingulate cortex (DMN), IFG pars opercularis (DAN), middle frontal gyrus (FPN), orbitofrontal cortex (LIM), claustrum (SOM), BANKSTS (VAN), and inferior occipital lobe (VIS). The widespread brain regions associated with the domain likely reflects the many different processes engaged during reasoning and executive function tasks and the interrelation of this domain with other cognition domains, as has been noted in previous reviews (Antonova et al., 2004; Rüsch et al., 2008). Contrary to the previous domains, evidence of publication bias was found for R&EF at the overall level which had very few reported nonsignificant correlations. R&EF was also found to be significantly correlated with the hippocampus and the amygdala, centres of memory and emotional processing. As mentioned, this domain is multidimensional and was thus expected to recruit those subcortical regions (Rüsch et al., 2008). However, the cerebellum did not pass FDR correction which was surprising. Indeed, we would expect that as an integrative hub of cognitive processes, the cerebellum would be engaged during problem solving and executive functions which utilize multiple domains. Indeed, the “error detection” and cognitive coordination functions of the cerebellum would be thought to be important for R&EF (Schmahmann, 2019).

#### 4.2.7. Social Cognition

Like R&EF, SC was also associated with all networks, confirming previous studies about these two domains potentially sharing neural correlates due to their higher-order nature (Moreau & Champagne-Lavau, 2014). However, the regions driving these network associations differed between the two cognitive domains. One pertinent example is the LIM, for which reasoning and executive function was associated with orbitofrontal cortex, while social cognition was associated with the parahippocampal gyrus, a region known for its encoding and retrieval function in memory and its proximity to the hippocampus (Diederen et al., 2010). Other SC- network correlations were driven by posterior cingulate gyrus (DMN), superior parietal lobule (DAN), IFG pars triangularis (FPN), superior temporal gyrus (SOM), middle cingulate gyrus (VAN), and middle occipital gyrus (VIS). These findings may point to distinct subnetworks subserving REAS&EF and SC in schizophrenia.

The extensive structural abnormalities observed are consistent with previous reviews and studies identifying multiple subdomains of SC and related structures (Buck, Healey, Gagen, Roberts, & Penn, 2016; Fujiwara et al., 2015; Green, Horan, & Lee, 2015). SC also contained the strongest correlation with overall brain regions relative to the other domains. Clear divisions of tasks assessing SC allowed us to divide the domain into emotion processing and theory of mind, which are among the most frequently studied in schizophrenia (Savla, Vella, Armstrong, Penn, & Twamley, 2013), finding that both domains contributed to these overall findings. These strong correlations may be additional evidence of the publication bias found in this domain. Concerning other structures, the amygdala, responsible for emotional processing, and the hippocampus, involved in the memory component of some tasks, were significantly correlated with SC. The high heterogeneity of this domain is also evidenced by the significant difference between the low- and high- quality studies.

#### 4.2.8. Verbal Fluency

VF was associated only with the DMN driven by the inferior temporal gyrus, which is known for its function in auditory and speech processing (Takahashi et al., 2011). Verbal fluency measures are frequently related to or combined within SP and ATT (Nuechterlein et al., 2004; Ojeda et al., 2010), and SP also showed an association with the DMN and particularly the inferior temporal gyrus; however, overlapping associations between these domains in other networks could not be determined due to few associations reported in the literature. The hippocampus was non-significant, but the cerebellum showed a significant association with VF. As mentioned, the cerebellum is involved in speed of processing and integration of information. It would be expected to have a strong correlation with VF specially since previous studies have found that VF is related to SP in healthy controls (Ojeda et al., 2010). Publication bias present in this domain as well as non-significant differences between low- and high- quality studies could be evidence that there are a lot of similarities in methodologies between included studies.

### 4.3. Strengths & Limitations

The current meta-analysis brings together the vast literature on brain structure and cognition in SZ in an effort to provide a reliable reference for the field and inform on gaps in the literature. We assessed the risk of bias for studies in the literature for each of the eight domains as well as the quality of the studies based on several criteria relevant to the field. The use of the random effects model and the quality assessment also remediated the high heterogeneity found in domains. Additionally, leveraging networks to investigate structure-cognition associations is an approach that brings the previous literature up to the current state of the field and provides interesting future research directions. Visualizing both the networks and the associated brain regions provides a multidimensional understanding of the associations between brain structure and cognition.

However, there were some limitations to this study. First, meta-analyses are prone to diluted summary effects when combining multiple studies with different methodologies. Though our overall meta-analyses combined all identified brain regions and metrics per domain, the emergence of significant effects across all domains speaks to the strength of the observed associations in schizophrenia. In addition, our network subgroup analyses aimed to address this heterogeneity in a way that incorporates biologically meaningful brain architecture. Second, we performed a large number of subgroup analyses to pinpoint brain structure-cognition associations across domains and networks. To remediate this, we included corrections for multiple comparisons and report only corrected results. Also, due to the nature of the network analyses we wanted to perform, we excluded general regions (i.e., lobes) or other structures (i.e., ventricles, thalamus, nucleus accumbens), which may hold relevant associations with cognitive domains. We may also consider additional criteria that could distinguish low- and high- quality studies. For example, previous domain-specific meta-analyses found that covariates including medication duration, IQ and the duration of memory recall based on different tests and the tasks themselves may impact results (Antoniades et al., 2018; Knowles et al., 2010). These and other potential factors were not consistently assessed in the literature and could not be included in the current study. Finally, there was a substantial bias in the morphological metrics reported by studies. There is a very large literature on volume changes in relation to cognition in schizophrenia but relatively few investigations about other metrics including cortical thickness, surface area and density. The partiality of the literature limits our understanding of the neurobiological underpinnings of cognitive deficits in SZ and should be explored in future research.

### 4.4. Conclusion

The current findings suggest strong associations between the most commonly assessed cognitive domains and overall brain structure in schizophrenia and provide insight into network associations and potential hub regions within those networks contributing to specific cognitive domains. We found that the number of networks associated with a given domain was often indicative of the complexity of that domain. We also identified multiple associations between the amygdala, the hippocampus and the cerebellum with cognition in SZ. The use of functional networks as a map for topographical studies yielded novel results and shed light on gaps in the literature concerning certain brain structures and biases that should be considered for some domains. We looked at structure within networks but investigating connectivity and between-network associations could be equally, if not more, important given the overlap between cognitive domains and associated brain networks. Additionally, future studies should focus on determining which of cortical thickness or surface area drives the volumetric changes robustly reported in the literature. More research in this area is also necessary with first-episode psychosis or at-risk populations in order to determine whether these structural and network associations with cognition are present in early stages of psychosis, which would provide a deeper understanding of the neurodevelopmental aspects of the disorder.

## Data Availability

All available data is in the supplementary material submitted

## Acknowledgements

The authors would like to acknowledge the contribution of Vanessa McGrory (VM) and Cai Li (CL) during the article selection process and the quality control of data extraction. We would also like to thank Geneviève Sauvé for her invaluable help during the analysis of the results, Gabrielle Pochiet for her help during the setting of the meta-analysis and Élisabeth Thibaudeau for her insight into social cognition.

