## Supplementary Material for "Structural Brain Correlates of Cognitive Function in Schizophrenia: A Meta-Analysis"

RUNNING HEAD: Brain Structure and Cognition in Schizophrenia

**SUPPLEMENTARY MATERIAL**

Marianne Khalil^1,2^, Philippine Hollander^2,3^, Delphine Raucher-Chéné^1,2,4,5^, Martin Lepage^1,2^, Katie M. Lavigne^1,2,6*^

¹ Department of Psychiatry, McGill University, Montreal, Quebec, Canada

² Douglas Mental Health University Institute, McGill University, Montreal, Quebec, Canada

^3^ Faculty of Psychology and Neuroscience, Maastricht University, Maastricht, Netherlands

^4^ Department of Psychiatry, University Hospital of Reims, EPSM Marne, Reims, France

^5^ Cognition, Health, and Society Laboratory (EA 6291), University of Reims, Champagne-Ardenne, Reims, France

^6^ Montreal Neurological Institute, McGill University, Montreal, Quebec, Canada

*Corresponding Author:

**Katie M. Lavigne,** F1128.1, 6875 boulevard Lasalle, Frank B. Common Pavilion, Douglas Mental Health University Institute, Montreal, QC H4H 1R3, Canada. Phone: 1- 514-761-6131 x 3384.

### **PRISMA Checklist (2020)**

| **Section and Topic** | **Item #** | **Checklist item** | **Location where item is reported** |
| --- | --- | --- | --- |
| **TITLE** | | |  |
| Title | 1 | Identify the report as a systematic review. | p. 1 |
| **ABSTRACT** | | |  |
| Abstract | 2 | See the PRISMA 2020 for Abstracts checklist. | p. 2 |
| **INTRODUCTION** | | |  |
| Rationale | 3 | Describe the rationale for the review in the context of existing knowledge. | p. 6 |
| Objectives | 4 | Provide an explicit statement of the objective(s) or question(s) the review addresses. | p. 6 |
| **METHODS** | | |  |
| Eligibility criteria | 5 | Specify the inclusion and exclusion criteria for the review and how studies were grouped for the syntheses. | p. 7 (2.3) |
| Information sources | 6 | Specify all databases, registers, websites, organisations, reference lists and other sources searched or consulted to identify studies. Specify the date when each source was last searched or consulted. | p. 6 (2.2) |
| Search strategy | 7 | Present the full search strategies for all databases, registers and websites, including any filters and limits used. | p. 6 (2.2) |
| Selection process | 8 | Specify the methods used to decide whether a study met the inclusion criteria of the review, including how many reviewers screened each record and each report retrieved, whether they worked independently, and if applicable, details of automation tools used in the process. | p. 8 (2.4) |
| Data collection process | 9 | Specify the methods used to collect data from reports, including how many reviewers collected data from each report, whether they worked independently, any processes for obtaining or confirming data from study investigators, and if applicable, details of automation tools used in the process. | p. 8 (2.5) |
| Data items | 10a | List and define all outcomes for which data were sought. Specify whether all results that were compatible with each outcome domain in each study were sought (e.g. for all measures, time points, analyses), and if not, the methods used to decide which results to collect. | p. 8 (2.5) |
|  | 10b | List and define all other variables for which data were sought (e.g. participant and intervention characteristics, funding sources). Describe any assumptions made about any missing or unclear information. | p. 8 (2.5) |
| Study risk of bias assessment | 11 | Specify the methods used to assess risk of bias in the included studies, including details of the tool(s) used, how many reviewers assessed each study and whether they worked independently, and if applicable, details of automation tools used in the process. | p. 11 (2.7.1.1) |
| Effect measures | 12 | Specify for each outcome the effect measure(s) (e.g. risk ratio, mean difference) used in the synthesis or presentation of results. | p.10-13 (2.7) |
| Synthesis methods | 13a | Describe the processes used to decide which studies were eligible for each synthesis (e.g. tabulating the study intervention characteristics and comparing against the planned groups for each synthesis (item #5)). | p. 7 (2.3) |
|  | 13b | Describe any methods required to prepare the data for presentation or synthesis, such as handling of missing summary statistics, or data conversions. | p.10-13 (2.7) |
|  | 13c | Describe any methods used to tabulate or visually display results of individual studies and syntheses. | p.10-13 (2.7) |
|  | 13d | Describe any methods used to synthesize results and provide a rationale for the choice(s). If meta-analysis was performed, describe the model(s), method(s) to identify the presence and extent of statistical heterogeneity, and software package(s) used. | p.10-13 (2.7) |
|  | 13e | Describe any methods used to explore possible causes of heterogeneity among study results (e.g. subgroup analysis, meta-regression). | p. 11 (2.7.1.1) |
|  | 13f | Describe any sensitivity analyses conducted to assess robustness of the synthesized results. | p. 9 (2.6) + p. 12 (2.7.1.2) |
| Reporting bias assessment | 14 | Describe any methods used to assess risk of bias due to missing results in a synthesis (arising from reporting biases). | - |
| Certainty assessment | 15 | Describe any methods used to assess certainty (or confidence) in the body of evidence for an outcome. | p. 10 (2.7) |
| **RESULTS** | | |  |
| Study selection | 16a | Describe the results of the search and selection process, from the number of records identified in the search to the number of studies included in the review, ideally using a flow diagram. | p. 14 (3.1) |
|  | 16b | Cite studies that might appear to meet the inclusion criteria, but which were excluded, and explain why they were excluded. | Table S2 |
| Study characteristics | 17 | Cite each included study and present its characteristics. | Table S2 |
| Risk of bias in studies | 18 | Present assessments of risk of bias for each included study. | p. 15 (3.3.1) |
| Results of individual studies | 19 | For all outcomes, present, for each study: (a) summary statistics for each group (where appropriate) and (b) an effect estimate and its precision (e.g. confidence/credible interval), ideally using structured tables or plots. | Table S2 |
| Results of syntheses | 20a | For each synthesis, briefly summarise the characteristics and risk of bias among contributing studies. | p. 15 (3.3.1) |
|  | 20b | Present results of all statistical syntheses conducted. If meta-analysis was done, present for each the summary estimate and its precision (e.g. confidence/credible interval) and measures of statistical heterogeneity. If comparing groups, describe the direction of the effect. | p. 14-19 |
|  | 20c | Present results of all investigations of possible causes of heterogeneity among study results. | p. 15 (3.3.1) |
|  | 20d | Present results of all sensitivity analyses conducted to assess the robustness of the synthesized results. | p. 15 (3.3.2) |
| Reporting biases | 21 | Present assessments of risk of bias due to missing results (arising from reporting biases) for each synthesis assessed. | - |
| Certainty of evidence | 22 | Present assessments of certainty (or confidence) in the body of evidence for each outcome assessed. | p. 15 (3.3.2) |
| **DISCUSSION** | | |  |
| Discussion | 23a | Provide a general interpretation of the results in the context of other evidence. | p. 19-27 |
|  | 23b | Discuss any limitations of the evidence included in the review. | p. 27 (4.3) |
|  | 23c | Discuss any limitations of the review processes used. | p. 27 (4.3) |
|  | 23d | Discuss implications of the results for practice, policy, and future research. | p. 28 (4.4) |
| **OTHER INFORMATION** | | |  |
| Registration and protocol | 24a | Provide registration information for the review, including register name and registration number, or state that the review was not registered. | p. 6 (2.1) |
|  | 24b | Indicate where the review protocol can be accessed, or state that a protocol was not prepared. | p. 6 (2.1) |
|  | 24c | Describe and explain any amendments to information provided at registration or in the protocol. | p. 7 (2.3) |
| Support | 25 | Describe sources of financial or non-financial support for the review, and the role of the funders or sponsors in the review. | p. 30 |
| Competing interests | 26 | Declare any competing interests of review authors. | p. 30 |

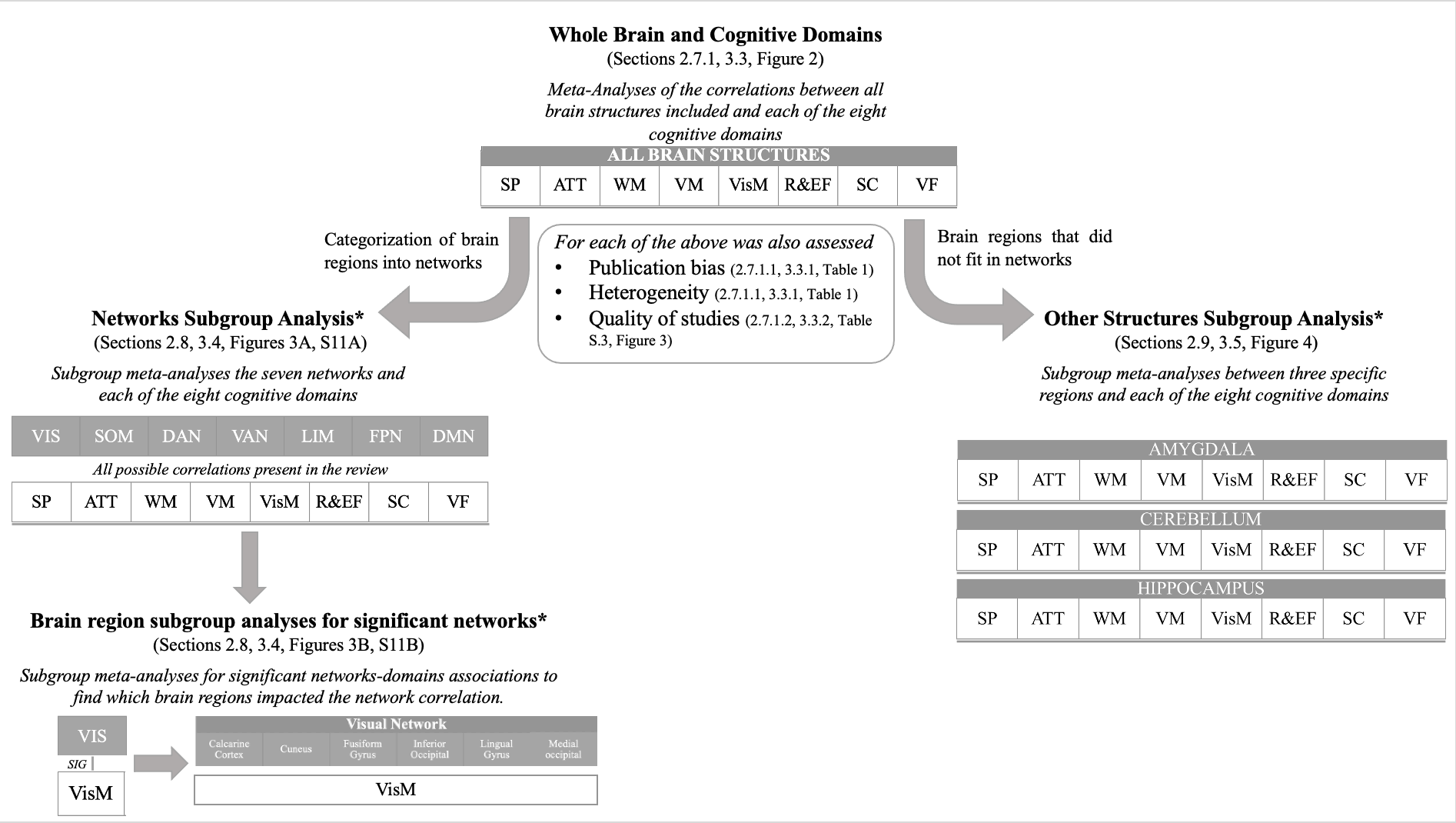

### **Figure S1.** Schematic process of the meta-analyses performed on the included data.

The sections and figures refer to where the methods or the results are discussed in the manuscript.

*These correlations were corrected for multiple comparisons using the false discovery rate (FDR) of 0.05.

### **Table S1.** Reported brain regions and the final categorization into brain structures or networks.

| **Final reported structures** | **Network**  **(if applicable)** | **Atlas/Segmentation**  **(if applicable)** |
| --- | --- | --- |
| Amygdala | - | - |
| Angular gyrus | DMN | WFU PickAtlas |
| Anterior cingulate gyrus | DMN | AUT |
| Anterior cingulate gyrus | DMN | DSK |
| Anterior cingulate gyrus | DMN | FreeSurfer Software |
| Anterior cingulate gyrus | DMN | SPM5 |
| Anterior cingulate gyrus | DMN | WFU PickAtlas |
| BANKSTS | VEN | DSK |
| Calcarine cortex | VIS | DSK |
| Caudate | - | - |
| Cerebellum | - | - |
| Claustrum | SOM | MNI |
| Corpus callosum | - | - |
| Cuneus | VIS | AUT |
| Dorsolateral prefrontal cortex | FPN | Broadmann |
| Entorhinal cortex | LIM | Broadmann |
| Frontal lobe | - | - |
| Fronto-temporal lobe | - | - |
| Fusiform gyrus | VIS | AUT |
| Fusiform gyrus | VIS | Broadmann |
| Fusiform gyrus | VIS | MAN |
| Grey matter | - | - |
| Gyrus rectus | LIM | AUT |
| Heschl's gyrus | SOM | Broadmann |
| Hippocampus | - | - |
| Inferior frontal gyrus | FPN | Broadman |
| Inferior frontal gyrus | FPN | DSK |
| Inferior frontal gyrus | FPN | WFU PickAtlas |
| IFG pars opercularis | DOR | Broadmann |
| IFG pars orbitalis | DMN | Broadmann |
| IFG pars triangularis | FPN | Broadmann |
| Inferior occipital | VIS | Broadmann |
| Inferior parietal lobule | DMN | AUT |
| Inferior parietal lobule | DMN | Broadmann |
| Inferior parietal lobule | DMN | MNI |
| Inferior parietal lobule | DMN | WFU PickAtlas |
| Inferior temporal gyrus | DMN | Broadmann |
| Inferior temporal gyrus | DMN | DKT |
| Insula | VEN | DKT |
| Insula | VEN | WFU PickAtlas |

| **Final reported structures** | **Network**  **(if applicable)** | **Atlas/Segmentation**  **(if applicable)** |
| --- | --- | --- |
| Lateral occipital | DMN | DKT |
| Lingual gyrus | VIS | Broadmann |
| Medial frontal cortex | FPN | AUT |
| Medial frontal cortex | FPN | Broadmann |
| Medial frontal cortex | FPN | MAN |
| Medial occipital lobe | VIS | AUT |
| Middle cingulate gyrus | VEN | AUT |
| Middle cingulate gyrus | VEN | WFU PickAtlas |
| Middle frontal gyrus | FPN | AUT |
| Middle frontal gyrus | FPN | Broadmann |
| Middle frontal gyrus | FPN | DSK |
| Middle occipital lobe | VIS | AUT |
| Middle temporal gyrus | DMN | AUT |
| Middle temporal gyrus | DMN | Broadmann |
| Nucleus accumbens | - | - |
| Occipital lobe | VIS | AUT |
| Operculum | VEN | Broadmann |
| Orbitofrontal cortex | LIM | AUT |
| Orbitofrontal cortex | LIM | TAL |
| Orbitofrontal cortex | LIM | WFU PickAtlas |
| Pallidum | - | - |
| Parahippocampal gyrus | LIM | AUT |
| Parietal lobe | - | - |
| Parieto-occipital lobe | - | - |
| Pituitary | - | - |
| Planum temporale | SOM | Broadmann |
| Posterior cingulate gyrus | DMN | AUT |
| Posterior cingulate gyrus | DMN | Broadmann |
| Precentral gyrus | SOM | DKT |
| Precuneus | DMN | MNI |
| Prefrontal cortex | - | - |
| Presubiculum | - | - |
| Putamen | - | - |
| Striatum | - | - |
| Subcortex | - | - |
| Subiculum | - | - |
| Superior frontal gyrus | VEN | AUT |
| Superior frontal gyrus | VEN | Broadmann |
| Superior parietal lobule | DOR | DKT |
| Superior temporal gyrus | SOM | AUT |
| Superior temporal gyrus | SOM | Broadmann |
| Supplementary motor area | SOM | MNI |
| Supramarginal gyrus | FPC | DKT |
| Temporal lobe | - | - |
| **Final reported structures** | **Network**  **(if applicable)** | **Atlas/Segmentation**  **(if applicable)** |
| Temporal pole | DMN | Broadmann |
| Temporal pole | DMN | MAN |
| Temporal pole | DMN | WFU PickAtlas |
| Temporal horns | - | - |
| Temporoparietal | - | - |
| Temporoparietal junction | VEN | - |
| Thalamus | - | - |
| Ventricles | - | - |
| White matter | - | - |
| Whole brain | - | - |

**For the networks:** DMN: Default mode network, DOR: dorsal attention network, FPN: frontoparietal network, LIM: limbic network, SOM: somatomotor network, VEN: ventral attention network, VIS: visual network. **For the atlases and segmentation methods**: AUT: automatic segmentation with a software, Broadmann: Broadmann areas, DKT: Desikian-Killiany Atlas, DSK: Destrieux cortical Atlas, FreeSurfer: Freesurfer software for brain mapping, MAN: manual segmentation based on reported criteria, MNI: Montreal Neurological Institute atlas, SPM5: Statistical Parametric Mapping (5th version), TAL: Talairach Atlas, WFU PickAtlas: Wake Forest University statist

### **Table S2.** Description of included articles in the literature review with the cognitive domains and structural metrics reported

| **Article** | **Brain region** | **Cognitive Domain** | **Cognitive test** | **Structural measure** | **Value of Effect** | **N patients** |
| --- | --- | --- | --- | --- | --- | --- |
| Abbs et al. (2011) | prefrontal cortex  prefrontal cortex  inferior parietal  hippocampus  prefrontal cortex  prefrontal cortex  prefrontal cortex  inferior parietal  hippocampus  prefrontal cortex | VM | CVLT Trial 5  CVLT Semantic Clustering  CVLT Semantic Clustering  CVLT Semantic Clustering  CVLT Serial Clustering  CVLT Trial 5  CVLT Semantic Clustering  CVLT Semantic Clustering  CVLT Semantic Clustering  CVLT Serial Clustering | Volume | r = 0.40  r = 0.09 (ns)  r = -0.04 (ns)  r = 0.32  r = 0.49  r = −0.51  r = -0.45  r = 0.38  r = 0.03 (ns)  r = -0.32 | 59  29 |
| Antonova, Sharma, Morris, and Kumari (2004) | *Review* |  | *Review* |  | *Review* |  |
| Antonova et al. (2005) | precuneus  occipital lobe white matter  occipital lobe white matter  whole brain | VM  ATT  ATT  VF | Buschke Selective Reminding Test - consistent recall  Stroop Test - words  Stroop Test - colors  Letter and Category verbal fluency - phonological | Volume | r = 0.40  r = 0.44 (ns)  r = 0.48 (ns)  r = 0.37 | 45 |
| Antoniades et al. (2018) | *Meta-Analysis* |  | *Meta-Analysis* |  | *Meta-Analysis* |  |
| Baare et al. (1999) | total prefrontal cortex  left prefrontal cortex  right prefrontal cortex  total orbital gyrus  grey orbital gyrus  total prefrontal cortex  right prefrontal cortex  total prefrontal cortex | VM  SP | CVLT - immediate recall  CVLT - immediate recall  CVLT - immediate recall  CVLT - immediate recall  CVLT - immediate recall  WMS-R Visual reproduction - immediate  WMS-R Visual reproduction - immediate  Semantic fluency | Volume | r = 0.46 (ns)  r = 0.69  r = 0.83  r = 0.32 (ns)  r = 0.37 (ns)  r = 0.6  r = 0.76  r = 0.64 | 13 |
| Banaj et al. (2018) | left inferior parietal cortex left insula  right middle temporal cortex  left cerebellum – lobule 6 (BA18) right cerebellum –lobuleVIIb  right cerebellum – CRUS 1 | VM  VisM | Rey's 15-word Immediate Recall Rey's 15-word Immediate Recall Rey-Osterrieth Complex Figure Test – delayed recall Rey-Osterrieth Complex Figure Test – delayed recall Rey-Osterrieth Complex Figure Test – delayed recall Rey-Osterrieth Complex Figure Test – delayed recall | Volume | t = 7.32 t = 6.16 t = 7.15  t = 6.23  t = 6.27 t = 6.49 | 28 |
| Bonilha et al. (2008) | BA 9 BA 9 BA 46 | R&EF | Trail making test B Wisconsin Card Sorting Task - errors Wisconsin Card Sorting Task - failure to maintain | Volume | r = −0.70* r = −0.65* r = −0.56* | 14 |
| Bornstein, Schwarzkopf, Olson, and Nasrallah (1992) | third ventricle  third ventricle  third ventricle  lateral ventricle  third ventricle  lateral ventricle  third ventricle  third ventricle  third ventricle  third ventricle  third ventricle | R&EF  VM  WM  SP  WM | Wisconsin Card Sorting Task - category  Wisconsin Card Sorting Task - perseveration  Verbal Concept Formation Test  WMS-R Visual Memory Span - forward  WMS-R Visual Memory Span - forward  WMS-R Visual Memory Span - backward  WMS-R Visual Memory Span - backward  WMS-R Digit Span - forward  WMS-R Digit Span - backward  Trail making test A  Knox cube delay | Volume | r = -0.34  r = 0.22*  r = -0.33  r = -0.33  r = -0.30  r = -0.29  r = -0.29  r = -0.29  r = -0.28  r = 0.23  r = -0.27 | 72 |
| Caldiroli et al. (2018) | right insula  left insula  right insula  left insula  right insula  left insula  right insula  left insula  right insula  left insula  right insula  left insula  right insula  left insula  right insula  left insula  right insula  left insula  right insula  left insula  right insula  left insula | VisM  SC | BFRT score  BFRT score  DFAR percentage neutral faces  DFAR percentage neutral faces  DFAR percentage happy faces  DFAR percentage happy faces  DFAR percentage fearful faces  DFAR percentage fearful faces  DFAR percentage angry faces  DFAR percentage angry faces  DFAR percentage (all faces)  DFAR percentage (all faces)  EMT second order belief score  EMT second order belief score  EMT second order emotion score  EMT second order emotion score  EMT first order emotion score  EMT first order emotion score  EMT first order belief score  EMT first order belief score  EMT control question score  EMT control question score | Volume | t = −0.63 (ns)  t = 0.50 (ns)  t = 0.61 (ns)  t = −0.72 (ns)  t = 0.91 (ns)  t = −0.28 (ns)  t = 0.15 (ns)  t = 1.86 (ns)  t = −0.19 (ns)  t = −0.84 (ns)  t = 0.05 (ns)  t = 0.48 (ns)  t = −0.17 (ns)  t = 0.69 (ns)  t = 0.82 (ns)  t = −1.00 (ns)  t = −0.38 (ns)  t = 1.51 (ns)  t = −0.04 (ns)  t = 0.14 (ns)  t = 0.25 (ns)  t = −1.01 (ns) | 133 |
| Crespo-Facorro, Barbadillo, Pelayo-Teran, and Rodriguez-Sanchez (2007) | *Review* |  | *Review* |  | *Review* |  |
| DeLisi et al. (1991) | left temporal lobe  right temporal lobe  left frontal lobe  right frontal lobe  left lateral ventricles  right lateral ventricles  left parahippocampal gyrus  right parahippocampal gyrus  left temporal lobe  right temporal lobe  left frontal lobe  right frontal lobe  left lateral ventricles  right lateral ventricles  left parahippocampal gyrus  right parahippocampal gyrus  left temporal lobe  right temporal lobe  left frontal lobe  right frontal lobe  left lateral ventricles  right lateral ventricles  left parahippocampal gyrus  right parahippocampal gyrus  left temporal lobe  right temporal lobe  left frontal lobe  right frontal lobe  left lateral ventricles  right lateral ventricles  left parahippocampal gyrus  right parahippocampal gyrus  left temporal lobe  right temporal lobe  left frontal lobe  right frontal lobe  left lateral ventricles  right lateral ventricles  left parahippocampal gyrus  right parahippocampal gyrus  left temporal lobe  right temporal lobe  left frontal lobe  right frontal lobe  left lateral ventricles  right lateral ventricles  left parahippocampal gyrus  right parahippocampal gyrus  left temporal lobe  right temporal lobe  left frontal lobe  right frontal lobe  left lateral ventricles  right lateral ventricles  left parahippocampal gyrus  right parahippocampal gyrus  left temporal lobe  right temporal lobe  left frontal lobe  right frontal lobe  left lateral ventricles  right lateral ventricles  left parahippocampal gyrus  right parahippocampal gyrus | VM  VisM  VM  VisM  R&EF  VF  R&EF | WMS - Logical memory  WMS - Logical memory  WMS - Logical memory  WMS - Logical memory  WMS - Logical memory  WMS - Logical memory  WMS - Logical memory  WMS - Logical memory  WMS associate learning  WMS associate learning  WMS associate learning  WMS associate learning  WMS associate learning  WMS associate learning  WMS associate learning  WMS associate learning  WMS visual reproduction  WMS visual reproduction  WMS visual reproduction  WMS visual reproduction  WMS visual reproduction  WMS visual reproduction  WMS visual reproduction  WMS visual reproduction  California Verbal Learning Test  California Verbal Learning Test  California Verbal Learning Test  California Verbal Learning Test  California Verbal Learning Test  California Verbal Learning Test  California Verbal Learning Test  California Verbal Learning Test  Benton Visual Retention Test  Benton Visual Retention Test  Benton Visual Retention Test  Benton Visual Retention Test  Benton Visual Retention Test  Benton Visual Retention Test  Benton Visual Retention Test  Benton Visual Retention Test  Wisconsin Card Sorting Test  Wisconsin Card Sorting Test  Wisconsin Card Sorting Test  Wisconsin Card Sorting Test  Wisconsin Card Sorting Test  Wisconsin Card Sorting Test  Wisconsin Card Sorting Test  Wisconsin Card Sorting Test  Controlled Oral Word Association  Controlled Oral Word Association  Controlled Oral Word Association  Controlled Oral Word Association  Controlled Oral Word Association  Controlled Oral Word Association  Controlled Oral Word Association  Controlled Oral Word Association  Trail making test B  Trail making test B  Trail making test B  Trail making test B  Trail making test B  Trail making test B  Trail making test B  Trail making test B | Volume | r = 0.10 (ns)  r = 0.06 (ns)  r = -0.11 (ns)  r = -0.17 (ns)  r = -0.15 (ns)  r = -0.04 (ns)  r = 0.33 (ns)  r = 0.31 (ns)  r = 0.07 (ns)  r = 0.04 (ns)  r = -0.11 (ns)  r = -0.19 (ns)  r = -0.08 (ns)  r = 0.09 (ns)  r = 0.45  r = 0.40  r = 0.16 (ns)  r = 0.09 (ns)  r = 0.20 (ns)  r = 0.17 (ns)  r = -0.22 (ns)  r = -0.05 (ns)  r = -0.31 (ns)  r = -0.17 (ns)  r = 0.10 (ns)  r = 0.17 (ns)  r = -0.01 (ns)  r = 0.00 (ns)  r = 0.04 (ns)  r = 0.14 (ns)  r = 0.13 (ns)  r = 0.13 (ns)  r = 0.15 (ns)  r = -0.02 (ns)  r = 0.17 (ns)  r = 0.07 (ns)  r = -0.25 (ns)  r = -0.12 (ns)  r = -0.05 (ns)  r = -0.08 (ns)  r = -0.02* (ns)  r = 0.05* (ns)  r = -0.21* (ns)  r = -0.14* (ns)  r = 0.33* (ns)  r = 0.09* (ns)  r = 0.14* (ns)  r = 0.12* (ns)  r = -0.05 (ns)  r = -0.06 (ns)  r = 0.06 (ns)  r = -0.02 (ns)  r = -0.13 (ns)  r = -0.07 (ns)  r = -0.04 (ns)  r = -0.09 (ns)  r = -0.11* (ns)  r = -0.11* (ns)  r = 0.05* (ns)  r = 0.05* (ns)  r = 0.23* (ns)  r = 0.29* (ns)  r = 0.01* (ns)  r = 0.00 (ns) | 45 |
| Dickey et al. (2007) | right hippocampus  left hippocampus  right hippocampus  left hippocampus  right hippocampus  left hippocampus  right hippocampus  left hippocampus  right hippocampus  left hippocampus | WM  VM | Delayed Alternation test - total errors  Delayed Alternation test - total errors  Delayed Alternation test - perseverations  Delayed Alternation test - perseverations  Delayed Alternation test - failure to maintain  Delayed Alternation test - failure to maintain  WMS-R Logical Memory 1  WMS-R Logical Memory 1  CVLT - total words  CVLT - total words | Volume | ρ = 0.098 (ns)  ρ = −0.528  ρ = 0.218 (ns)  ρ = −0.435  ρ = −0.191 (ns)  ρ = −0.468  ρ = −0.262 (ns)  ρ = −0.104 (ns)  ρ = −0.205 (ns)  ρ = 0.112 (ns) | 20 |
| Edgar et al. (2012) | left Heschel's gyrus  left planum temporale  left lateral aspect  right Heschl's gyrus  right planum temporale  right lateral aspect  left inferior orbital cortex  left inferior triangularis  left middle frontal gyrus  left inferior opercular gyrus  right inferior opercular cortex  right middle frontal gyrus  right inferior orbital  right inferior triangularis | ATT | TMT A + Connors' Continuous Performance Test  TMT A + Connors' Continuous Performance Test  TMT A + Connors' Continuous Performance Test  TMT A + Connors' Continuous Performance Test  TMT A + Connors' Continuous Performance Test  TMT A + Connors' Continuous Performance Test  TMT A + Connors' Continuous Performance Test  TMT A + Connors' Continuous Performance Test  TMT A + Connors' Continuous Performance Test  TMT A + Connors' Continuous Performance Test  TMT A + Connors' Continuous Performance Test  TMT A + Connors' Continuous Performance Test  TMT A + Connors' Continuous Performance Test  TMT A + Connors' Continuous Performance Test | Cortical Thickness | r = 0.46  r = 0.34 (ns)  r = 0.26 (ns)  r = 0.54  r = 0.36 (ns)  r = 0.05 (ns)  r = 0.52  r = 0.31  r = 0.3  r = 0.18 (ns)  r = 0.48  r = 0.33  r = 0.18 (ns)  r = 0.25 (ns) | 51 |
| Ehrlich et al. (2012) | right middle/sup temporal gyrus  left hippocampus  right hippocampus  left hippocampus  right hippocampus  left hippocampus  right hippocampus  left hippocampus  right hippocampus | WM  VM  VisM  VM | WAIS letter number sequencing - recall  WAIS letter number sequencing - recall  WAIS letter number sequencing - recall  Hopkins Verbal Learning Test-Revised - learning  Hopkins Verbal Learning Test-Revised - learning  Benton Visual Retention Test  Benton Visual Retention Test  WMS - Logical memory immediate recall  WMS - Logical memory immediate recall | Thickness  Volume | r = 0.391  t = 2.386  t = 1.951  t = 2.951  t = 1.764  t = 2.123  t = 1.014  t = 2.517  t = 2.038 | 131 |
| Exner, Boucsein, Degner, Irle, and Weniger (2004) | right amygdala  right amygdala | VisM  SC | WMS-R Visual paired associates immediate recall  ERT Emotion recognition | Volume | r = 0.832  r = 0.612 | 16 |
| Exner et al. (2008) | right hippocampus  right hippocampus | VisM | WMS-R Visual Reproduction 1  WMS-R Visual Reproduction 1 | Volume | r = 0.54  r = 0.23 | 14  7 |
| Francis et al. (2016) | posterior splenium  anterior splenium  posterior splenium  anterior splenium  posterior splenium  anterior splenium  posterior splenium  anterior splenium  posterior splenium  anterior splenium  posterior splenium  anterior splenium  posterior splenium  anterior splenium  posterior splenium  anterior splenium  posterior splenium  anterior splenium  posterior splenium  anterior splenium | VM  WM  VF  SP  R&EF  VM  WM  VF  SP  R&EF | BACS verbal memory  BACS verbal memory  BACS digit sequencing  BACS digit sequencing  BACS verbal fluency  BACS verbal fluency  BACS symbol coding  BACS symbol coding  BACS tower test  BACS tower test  BACS verbal memory  BACS verbal memory  BACS digit sequencing  BACS digit sequencing  BACS verbal fluency  BACS verbal fluency  BACS symbol coding  BACS symbol coding  BACS tower test  BACS tower test | Volume | r = 0.12 (ns)  r = 0.13 (ns)  r = 0.13 (ns)  r = 0.05 (ns)  r = 0.12 (ns)  r = 0.08 (ns)  r = 0.14 (ns)  r = 0.13 (ns)  r = 0.09 (ns)  r = 0.07 (ns)  r = 0.02 (ns)  r = 0.10 (ns)  r = -0.06 (ns)  r = -0.06 (ns)  r = -0.001 (ns)  r = -0.001 (ns)  r = -0.10 (ns)  r = -0.10 (ns)  r = 0.03 (ns)  r = 0.03 (ns) | 224  142 |
| Frascarelli et al. (2015) | left medial frontal gyrus  left medial frontal gyrus  left medial frontal gyrus  left medial frontal gyrus | R&EF | WCST Perseverative responses  WCST Perseverative errors  WCST Perseverative responses  WCST Perseverative errors | Volume | R² = 0.018 (ns)  R² = 0.187 (ns)  R² = 0.079 (ns)  R² = 0.297 (ns) | 18  15 |
| Fujiwara et al. (2007) | left cingulate sulcus  left anterior cingulate cortex  right anterior cingulate cortex  left anterior cingulate cortex  right anterior cingulate cortex  right anterior cingulate cortex | SC | Perception of Affect Tasks (PAT) - Subtask 1  Perception of Affect Tasks (PAT) - Subtask 2  Perception of Affect Tasks (PAT) - Subtask 3  Perception of Affect Tasks (PAT) - Subtask 3  Perception of Affect Tasks (PAT) - Subtask 4  Perception of Affect Tasks (PAT) - Subtask 4 | Volume | t = 2.899  r = 0.399  r = 0.415  r = 0.433  r = 0.48  r = 0.446 | 26 |
| Fujiwara, Yassin, and Murai (2015) | *Review* |  | *Review* |  | *Review* |  |
| Garlinghouse, Roth, Isquith, Flashman, and Saykin (2010) | left frontal  right frontal  left temporal  right temporal  left parietal  right parietal  total intracranial | WM | WAIS-III Digit span backwards  WAIS-III Digit span backwards  WAIS-III Digit span backwards  WAIS-III Digit span backwards  WAIS-III Digit span backwards  WAIS-III Digit span backwards  WAIS-III Digit span backwards | Volume | r = −0.40  r = −0.28 (ns)  r = −0.45  r = −0.46  r = 0.17 (ns)  r = 0.03 (ns)  r = 0.25 (ns) | 29 |
| (Goghari, MacDonald, & Sponheim, 2014) | bilateral superior prefrontal  left inferior prefrontal  right inferior prefrontal  bilateral superior prefrontal  left inferior prefrontal  right inferior prefrontal | WM | Spatial working memory task (hold)  Spatial working memory task (hold)  Spatial working memory task (hold)  Spatial working memory task (flip)  Spatial working memory task (flip)  Spatial working memory task (flip) | Volume | r = 0.46 (ns)  r = 0.33 (ns)  r = 0.11 (ns)  r = 0.57  r = 0.56  r = 0.33 (ns) | 17 |
| Goldberg, Torrey, Berman, and Weinberger (1994) | left hippocampus  right hippocampus  lateral ventricles  third ventricle  left hippocampus  right hippocampus  lateral ventricles  third ventricle  left hippocampus  right hippocampus  lateral ventricles  third ventricle  left hippocampus  right hippocampus  lateral ventricles  third ventricle  left hippocampus  right hippocampus  lateral ventricles  third ventricle  left hippocampus  right hippocampus  lateral ventricles  third ventricle  left hippocampus  right hippocampus  lateral ventricles  third ventricle  left hippocampus  right hippocampus  lateral ventricles  third ventricle  left hippocampus  right hippocampus  lateral ventricles  third ventricle | ATT  VM  VisM  VM  SP  R&EF  VF | Continuous Performance Test  Continuous Performance Test  Continuous Performance Test  Continuous Performance Test  Stroop Color Test  Stroop Color Test  Stroop Color Test  Stroop Color Test  Logical memory  Logical memory  Logical memory  Logical memory  Visual reproduction  Visual reproduction  Visual reproduction  Visual reproduction  Paired association  Paired association  Paired association  Paired association  TMT A  TMT A  TMT A  TMT A  WCST - categories  WCST - categories  WCST - categories  WCST - categories  WCST - perseverative errors  WCST - perseverative errors  WCST - perseverative errors  WCST - perseverative errors  Verbal fluency test  Verbal fluency test  Verbal fluency test  Verbal fluency test | Volume | r = 0.009 (ns)  r = 0.18 (ns)  r = -0.17 (ns)  r = -0.28 (ns)  r = -0.16 (ns)  r = -0.15 (ns)  r = 0.34 (ns)  r = -0.13 (ns)  r = 0.57  r = 0.05 (ns)  r = -0.18 (ns)  r = -0.18 (ns)  r = -0.19 (ns)  r = -0.05 (ns)  r = -0.19 (ns)  r = -0.02 (ns)  r = -0.45 (ns)  r = -0.5 (ns)  r = 0.28 (ns)  r = 0.1 (ns)  r = -0.12 (ns)  r = -0.1 (ns)  r = 0.08 (ns)  r = -0.08 (ns)  r = -0.06 (ns)  r = -0.25 (ns)  r = 0.39 (ns)  r = -0.33 (ns)  r = 0.29* (ns)  r = 0.49* (ns)  r = -0.52*  r = 0.03* (ns)  r = -0.32 (ns)  r = -0.2 (ns)  r = 0.27 (ns)  r = 0.33 (ns) | 15 |
| Goldstein et al. (2011) | right BA44  right BA45 | WM | CANTAB - spatio-visual working memory  CANTAB - spatio-visual working memory | Volume | r = 0.04 (ns)  r = -0.02 (ns) | 29 |
| R. E. Gur et al. (1998) | *Some results missing* |  |  |  |  |  |
| R. C. Gur, Ragland, and Gur (1997) | *Review* |  | *Review* |  | *Review* |  |
| R. E. Gur, Turetsky, Bilker, and Gur (1999) | grey matter  grey matter  grey matter  grey matter  grey matter  grey matter  grey matter  grey matter | R&EF  VM  VisM  R&EF | Neuropsychological tests - abstraction**  Neuropsychological tests - abstraction**  Neuropsychological tests - verbal memory***  Neuropsychological tests - verbal memory***  Neuropsychological tests - spatial memory ****  Neuropsychological tests - spatial memory ****  Neuropsychological tests - spatial abilities *****  Neuropsychological tests - spatial abilities ***** | Volume | r = 0.15 (ns)  r = 0.2 (ns)  r = 0.41  r = 0.36  r = 0.33  r = -0.04 (ns)  r = 0.42  r = 0.13 (ns) | 75  55  75  55  75  55  75  55 |
| R. E. Gur, Cowell, et al. (2000) | orbitomedial prefrontal | VM | Composite score verbal****** | Volume | r = 0.49 | 30 |
| R. E. Gur, Turetsky, et al. (2000) | hippocampus  hippocampus | VM | Neurocognitive battery - verbal memory***  Neurocognitive battery - verbal memory*** | Volume | r = 0.35  r = 0.26 | 58  42 |
| Habets et al. (2008) | *Some missing results* |  |  |  |  |  |
| Hanford, Pinnock, Hall, and Heinrichs (2019) | *Some results are missing* |  |  |  |  |  |
| Hartberg, Sundet, Rimol, Haukvik, Lange, Nesvag, Dale, et al. (2011) | left temporal pole  left transverse temporal  left pars opercularis  right fusiform  left transverse temporal | SP  WM  R&EF | WAIS-III Digit Symbol  WAIS-III Digit Symbol  WAIS-III Digit Span  D-KEFS Color-Word Inhibition  D-KEFS Color-Word Inhibition | Cortical Thickness | r = 0.27 (ns)  r = 0.28  r = 0.24 (ns)  r = -0.25 (ns)  r = -0.24 (ns) | 117 |
| Hartberg, Sundet, Rimol, Haukvik, Lange, Nesvag, Melle, et al. (2011) | Left putamen Right putamen Left putamen  Right putamen Left putamen  Right putamen Left putamen  Right putamen | VM  WM  R&EF | CVLT - list A1-A5 CVLT - list A1-A6 WAIS-III Digit Span WAIS-III Digit Span  D-KEFS Color-Word interference test D-KEFS Color-Word interference test D-KEFS Verbal Fluency - category switching D-KEFS Verbal Fluency - category switching | Volume | r = -0.26 r = -0.28  r = -0.28 r = -0.27 r = 0.3 r = 0.25 (ns) r = -0.31  r = -0.27 | 117 |
| Herold et al. (2015) | Left hippocampus  Right hippocampus  Left anterior hippocampus  Left posterior hippocampus  Right anterior hippocampus  Right posterior hippocampus  Left hippocampus  Right hippocampus  Left anterior hippocampus  Left posterior hippocampus  Right anterior hippocampus  Right posterior hippocampus  Left hippocampus  Right hippocampus  Left anterior hippocampus  Left posterior hippocampus  Right anterior hippocampus  Right posterior hippocampus  Left hippocampus  Right hippocampus  Left anterior hippocampus  Left posterior hippocampus  Right anterior hippocampus  Right posterior hippocampus  Left hippocampus  Right hippocampus  Left anterior hippocampus  Left posterior hippocampus  Right anterior hippocampus  Right posterior hippocampus  Left hippocampus  Right hippocampus  Left anterior hippocampus  Left posterior hippocampus  Right anterior hippocampus  Right posterior hippocampus  Left hippocampus  Right hippocampus  Left anterior hippocampus  Left posterior hippocampus  Right anterior hippocampus  Right posterior hippocampus | VM  WM  SP  R&EF  VisM | WMS - Logical memory immediate recall WMS - Logical memory immediate recall WMS - Logical memory immediate recall WMS - Logical memory immediate recall WMS - Logical memory immediate recall WMS - Logical memory immediate recall WMS - Logical memory delayed recall WMS - Logical memory delayed recall WMS - Logical memory delayed recall WMS - Logical memory delayed recall WMS - Logical memory delayed recall WMS - Logical memory delayed recall Digit span forward Digit span forward Digit span forward Digit span forward Digit span forward Digit span forward Digit span backward Digit span backward Digit span backward Digit span backward Digit span backward Digit span backward Trail-Making Test A Trail-Making Test A Trail-Making Test A Trail-Making Test A Trail-Making Test A Trail-Making Test A Trail-Making Test B Trail-Making Test B Trail-Making Test B Trail-Making Test B Trail-Making Test B Trail-Making Test B Bielefelder Famous Faces Test(BFFT) Bielefelder Famous Faces Test(BFFT) Bielefelder Famous Faces Test(BFFT) Bielefelder Famous Faces Test(BFFT) Bielefelder Famous Faces Test(BFFT) Bielefelder Famous Faces Test(BFFT) | volume | r=0.201(ns)  r=0.33  r=0.223(ns)  r=0.18(ns)  r=0.344  r=0.314  r=0.155(ns)  r=0.294  r=0.194(ns)  r=0.124(ns)  r=0.321  r=0.271(ns)  r=0.201(ns)  r=0.087(ns)  r=0.204(ns)  r=0.193(ns)  r=0.113(ns)  r=0.068(ns)  r=0.26(ns)  r=0.291  r=0.319  r=0.213(ns)  r=0.325  r=0.264(ns)  r=0.181(ns)  r=0.141(ns)  r=0.193(ns)  r=0.167(ns)  r=0.142(ns)  r=0.137(ns)  r=0.446*  r=0.426*  r=0.466*  r=0.419*  r=0.412*  r=0.427*  r=0.073(ns)  r=0.03(ns)  r=0.098(ns)  r=0.054(ns)  r=0.054(ns)  r=0.014(ns) | 48 |
| Hidese et al. (2017) | *Not included in meta-analyses* |  | *Results not compatible with software* |  |  |  |
| Hirao et al. (2008) | posterior dorsolat prefrontal cortex  left ventrolateral prefrontal cortex  anterior dorsolateral prefrontal  right insula  ventro-medial prefrontal cortex  anterior cingulate cortex | SC | ToM task - Reading the Mind in the Eyes test  ToM task - Reading the Mind in the Eyes test  ToM task - Reading the Mind in the Eyes test  ToM task - Reading the Mind in the Eyes test  ToM task - Reading the Mind in the Eyes test  ToM task - Reading the Mind in the Eyes test | Volume | r = 0.24 (ns)  r = 0.48  r = 0.19 (ns)  r = 0.26 (ns)  r = 0.13 (ns)  r = 0.14 (ns) | 20 |
| Hooker, Bruce, Lincoln, Fisher, and Vinogradov (2011) | left anterior sup temporal sulcus  right superior parietal gyrus  left anterior cingulate sulcus  left anterior cingulate cortex  right superior frontal gyrus  right superior temporal gyrus  right mid cingulate/suppl. motor  right sup frontal gyrus ant/ dors  left middle occipital gyrus  left sup frontal/sup orbital gyrus  right anterior cingulate cortex  right hippocampus  left anterior cingulate cortex  left supplementary motor area  left sup temporal/Herschl's gyrus  left rolandic operculum/insula  right mid occipital gyrus post  left precuneus  right rolandic operculum/insula  left middle frontal gyrus - anterior  right middle occipital gyrus - post  right posterior cingulate cortex  right superior frontal gyrus  right supplementary motor area  left middle/ant cingulate cortex  right rolandic operculum (SRC)  left angular gyrus  right insula/rolandic operculum  right superior frontal gyrus  right middle frontal gyrus - ant.  right anterior orbital gyrus  right middle cingulate gyrus  left precentral gyrus  right anterior cingulate gyrus  left supplementary motor area  left inf frontal gyrus-triangularis  right post insula/rolandic operc  left ant cingulate/cingulate sulcus  right anterior insula (SRC)  left post insula/rolandic operc  left sup frontal gyrus dor/ant  left orbital frontal gyrus - ant  right ant cingulate/supraorbital sul  left precentral gyrus  left middle frontal gyrus - ant  right anterior cingulate cortex  left middle frontal gyrus  right sup frontal gyrus/cing sulc  left precentral gyrus  right precuneus  right inferior frontal gyrus-triang  right supplementary motor area  right precuneus | SC | Faux Pas Score  Faux Pas Score  Faux Pas Score  Faux Pas Score  Faux Pas Score  Faux Pas Score  Faux Pas Score  Faux Pas Score  Faux Pas Score  Faux Pas Score  Faux Pas Score  IRI Perspective-Taking  IRI Perspective-Taking  IRI Perspective-Taking  IRI Perspective-Taking  IRI Perspective-Taking  IRI Perspective-Taking  IRI Perspective-Taking  IRI Perspective-Taking  IRI Perspective-Taking  IRI Perspective-Taking  IRI Perspective-Taking  IRI Perspective-Taking  IRI Perspective-Taking  IRI Perspective-Taking  IRI Perspective-Taking  IRI Perspective-Taking  IRI Perspective-Taking  QLS-Empathy  QLS-Empathy  QLS-Empathy  QLS-Empathy  QLS-Empathy  QLS-Empathy  QLS-Empathy  QLS-Empathy  QLS-Empathy  QLS-Empathy  QLS-Empathy  QLS-Empathy  QLS-Empathy  QLS-Empathy  QLS-Empathy  QLS-Empathy  QLS-Empathy  QLS-Empathy  QLS-Empathy  QLS-Empathy  QLS-Empathy  QLS-Empathy  QLS-Empathy  QLS-Empathy  QLS-Empathy | Volume | t = 4.80 (ns)  t = 4.66 (ns)  t = 4.43 (ns)  t = 3.97 (ns)  t = 5.15 (ns)  t = 5.04 (ns)  t = 4.26 (ns)  t = 4.31 (ns)  t = 4.40 (ns)  t = 4.35 (ns)  t = 3.75 (ns)  t = 7.09 (ns)  t = 5.81  t = 5.57 (ns)  t = 4.72 (ns)  t = 4.71 (ns)  t = 5.21 (ns)  t = 6.35 (ns)  t = 4.71 (ns)  t = 4.28 (ns)  t = 4.12 (ns)  t = 4.05 (ns)  t = 5.33 (ns)  t = 4.83 (ns)  t = 4.41 (ns)  t = 4.36 (ns)  t = 4.35 (ns)  t = 4.09 (ns)  t = 6.50 (ns)  t = 5.71 (ns)  t = 4.87 (ns)  t = 8.05 (ns)  t = 4.59 (ns)  t = 6.89  t = 5.18 (ns)  t = 4.90 (ns)  t = 4.77 (ns)  t = 5.15  t = 5.40 (ns)  t = 4.32 (ns)  t = 4.29 (ns)  t = 4.24 (ns)  t = 4.13 (ns)  t = 4.59 (ns)  t = 3.92 (ns)  t = 4.15 (ns)  t = 3.72 (ns)  t = 4.49 (ns)  t = 4.92 (ns)  t = 4.44 (ns)  t = 4.30 (ns)  t = 4.16 (ns)  t = 3.94 (ns) | 21 |
| Hoptman et al. (2005) | right OFC grey  right OFC white  right OFC grey  left OFC white  right OFC white | ATT  R&EF | Attention and Processing Speed  Attention and Processing Speed  TMT B Errors  TMT B Errors  WCST Perseverative errors | Volume | r = −0.48  r = -0.55  r = 0.45*  r = 0.51*  r = -0.40* | 49 |
| Hoseth et al. (2016) | hippocampal formation  CA1  CA2/3  CA4/DG  Presubiculum  Subiculum  hippocampal formation  CA1  CA2/3  CA4/DG  Presubiculum  Subiculum  hippocampal formation  CA1  CA2/3  CA4/DG  Presubiculum  Subiculum  hippocampal formation  CA1  CA2/3  CA4/DG  Presubiculum  Subiculum | VM | WMS-III - Logical Memory immediate recall  WMS-III - Logical Memory immediate recall  WMS-III - Logical Memory immediate recall  WMS-III - Logical Memory immediate recall  WMS-III - Logical Memory immediate recall  WMS-III - Logical Memory immediate recall  WMS-III - Logical Memory delayed recall  WMS-III - Logical Memory delayed recall  WMS-III - Logical Memory delayed recall  WMS-III - Logical Memory delayed recall  WMS-III - Logical Memory delayed recall  WMS-III - Logical Memory delayed recall  CVLT - Summed recall over learning list  CVLT - Summed recall over learning list  CVLT - Summed recall over learning list  CVLT - Summed recall over learning list  CVLT - Summed recall over learning list  CVLT - Summed recall over learning list  CVLT - Delayed free recall  CVLT - Delayed free recall  CVLT - Delayed free recall  CVLT - Delayed free recall  CVLT - Delayed free recall  CVLT - Delayed free recall | Volume | t = 0.48 (ns)  t = 0.42 (ns)  t = 0.77 (ns)  t = 0.92 (ns)  t = 0.06 (ns)  t = 0.73 (ns)  t = 1.49 (ns)  t = 0.57 (ns)  t = 0.90 (ns)  t = 1.13 (ns)  t = 0.78 (ns)  t = 1.00 (ns)  t = 2.78  t = 1.78 (ns)  t = 2.87  t = 2.72  t = 2.05  t = 3.04  t = 2.71  t = 2.23  t = 2.89  t = 2.72  t = 2.19  t = 2.91 | 46 |
| Kareken et al. (1995) | whole brain  whole brain  whole brain  whole brain  whole brain | R&EF  ATT  VM  VisM  R&EF | Neuropsychological tests - abstraction/mental flex  Neuropsychological tests - attention  Neuropsychological tests - verbal memory  Neuropsychological tests - visuo-spatial memory  Neuropsychological tests - visuo-spatial perception | Volume | r = 0.28  r = 0.12 (ns)  r = 0.21  r = 0.20 (ns)  r = 0.18 (ns) | 68 |
| Karnik-Henry et al. (2012) | left hippocampus | VM/VisM | Neuropsychological tests (episodic memory: family pictures, logical memory, CVLT) | Volume | r = 0.29 | 39 |
| Kelly et al. (2019) | *Review* |  | *Review* |  | *Review* |  |
| Killgore, Rosso, Gruber, and Yurgelun-Todd (2009) | left amygdala  left amygdala  right amygdala | VM  VisM | WMS - Prose recall immediate  WMS- Prose recall delayed  WMS - Figure reproduction delayed | Volume | r = -0.51  r = -0.49  r = -0.57 | 19 |
| Knochel, Reuter, et al. (2016) | left pars opercularis (IFG)  left pars opercularis (IFG) | SP  R&EF | Trail making test A  Trail making test B | Cortical Thickness | ρ = -0.52*  ρ = -0.84* | 32 |
| Knochel, Stablein, et al. (2016) | bilateral thalamus  left middle frontal gyrus  left superior frontal gyrus  left middle frontal gyrus  left middle frontal gyrus | VM  SP  R&EF | HVLT-R  HVLT-R  HVLT-R  Trail Making Test A  Trail Making Test B | Volume | r = −0.51  r = 0.64  r = 0.52  r = −0.52  r = -0.61 | 57 |
| Kochunov et al. (2020) | grey matter thickness  subcortical volume  grey matter thickness  subcortical volume  grey matter thickness  subcortical volume  grey matter thickness  subcortical volume  grey matter thickness  subcortical volume  grey matter thickness  subcortical volume | SP  WM  VM  VisM  R&EF  SC | MCCB speed of processing  MCCB speed of processing  MCCB working memory  MCCB working memory  MCCB verbal learning and memory  MCCB verbal learning and memory  MCCB visual learning and memory  MCCB visual learning and memory  MCCB reasoning and problem solving  MCCB reasoning and problem solving  MCCB social cognition  MCCB social cognition | Cort Thick  Volume  Cort Thick  Volume  Cort Thick  Volume  Cort Thick  Volume  Cort Thick  Volume  Cort Thick  Volume | r = -0.14 (ns)  r = -0.21  r = -0.19  r = -0.20  r = -0.14 (ns)  r = -0.28  r = -0.26  r = -0.08 (ns)  r = -0.17 (ns)  r = -0.11 (ns)  r = -0.05 (ns)  r = -0.13 (ns) | 28 |
| Koshiyama et al. (2018a) | left hippocampus  right hippocampus  left amygdala  right amygdala  left thalamus  right thalamus  left nucleus accumbens  right nucleus accumbens  left caudate  right caudate  left putamen  right putamen  left pallidum  right pallidum  left hippocampus  right hippocampus  left amygdala  right amygdala  left thalamus  right thalamus  left nucleus accumbens  right nucleus accumbens  left caudate  right caudate  left putamen  right putamen  left pallidum  right pallidum  left hippocampus  right hippocampus  left amygdala  right amygdala  left thalamus  right thalamus  left nucleus accumbens  right nucleus accumbens  left caudate  right caudate  left putamen  right putamen  left pallidum  right pallidum  left hippocampus  right hippocampus  left amygdala  right amygdala  left thalamus  right thalamus  left nucleus accumbens  right nucleus accumbens  left caudate  right caudate  left putamen  right putamen  left pallidum  right pallidum  left hippocampus  right hippocampus  left amygdala  right amygdala  left thalamus  right thalamus  left nucleus accumbens  right nucleus accumbens  left caudate  right caudate  left putamen  right putamen  left pallidum  right pallidum  left hippocampus  right hippocampus  left amygdala  right amygdala  left thalamus  right thalamus  left nucleus accumbens  right nucleus accumbens  left caudate  right caudate  left putamen  right putamen  left pallidum  right pallidum  left hippocampus  right hippocampus  left amygdala  right amygdala  left thalamus  right thalamus  left nucleus accumbens  right nucleus accumbens  left caudate  right caudate  left putamen  right putamen  left pallidum  right pallidum  left hippocampus  right hippocampus  left amygdala  right amygdala  left thalamus  right thalamus  left nucleus accumbens  right nucleus accumbens  left caudate  right caudate  left putamen  right putamen  left pallidum  right pallidum  left hippocampus  right hippocampus  left amygdala  right amygdala  left thalamus  right thalamus  left nucleus accumbens  right nucleus accumbens  left caudate  right caudate  left putamen  right putamen  left pallidum  right pallidum | WM  SP  WM  R&EF  SP | WAIS-III working memory  WAIS-III working memory  WAIS-III working memory  WAIS-III working memory  WAIS-III working memory  WAIS-III working memory  WAIS-III working memory  WAIS-III working memory  WAIS-III working memory  WAIS-III working memory  WAIS-III working memory  WAIS-III working memory  WAIS-III working memory  WAIS-III working memory  WAIS-III processing speed  WAIS-III processing speed  WAIS-III processing speed  WAIS-III processing speed  WAIS-III processing speed  WAIS-III processing speed  WAIS-III processing speed  WAIS-III processing speed  WAIS-III processing speed  WAIS-III processing speed  WAIS-III processing speed  WAIS-III processing speed  WAIS-III processing speed  WAIS-III processing speed  WAIS-III arithmetic  WAIS-III arithmetic  WAIS-III arithmetic  WAIS-III arithmetic  WAIS-III arithmetic  WAIS-III arithmetic  WAIS-III arithmetic  WAIS-III arithmetic  WAIS-III arithmetic  WAIS-III arithmetic  WAIS-III arithmetic  WAIS-III arithmetic  WAIS-III arithmetic  WAIS-III arithmetic  WAIS-III digit span  WAIS-III digit span  WAIS-III digit span  WAIS-III digit span  WAIS-III digit span  WAIS-III digit span  WAIS-III digit span  WAIS-III digit span  WAIS-III digit span  WAIS-III digit span  WAIS-III digit span  WAIS-III digit span  WAIS-III digit span  WAIS-III digit span  WAIS-III letter number sequencing  WAIS-III letter number sequencing  WAIS-III letter number sequencing  WAIS-III letter number sequencing  WAIS-III letter number sequencing  WAIS-III letter number sequencing  WAIS-III letter number sequencing  WAIS-III letter number sequencing  WAIS-III letter number sequencing  WAIS-III letter number sequencing  WAIS-III letter number sequencing  WAIS-III letter number sequencing  WAIS-III letter number sequencing  WAIS-III letter number sequencing  WAIS-III Block Design  WAIS-III Block Design  WAIS-III Block Design  WAIS-III Block Design  WAIS-III Block Design  WAIS-III Block Design  WAIS-III Block Design  WAIS-III Block Design  WAIS-III Block Design  WAIS-III Block Design  WAIS-III Block Design  WAIS-III Block Design  WAIS-III Block Design  WAIS-III Block Design  WAIS-III Matrix Reasoning  WAIS-III Matrix Reasoning  WAIS-III Matrix Reasoning  WAIS-III Matrix Reasoning  WAIS-III Matrix Reasoning  WAIS-III Matrix Reasoning  WAIS-III Matrix Reasoning  WAIS-III Matrix Reasoning  WAIS-III Matrix Reasoning  WAIS-III Matrix Reasoning  WAIS-III Matrix Reasoning  WAIS-III Matrix Reasoning  WAIS-III Matrix Reasoning  WAIS-III Matrix Reasoning  WAIS-III Digit symbol coding  WAIS-III Digit symbol coding  WAIS-III Digit symbol coding  WAIS-III Digit symbol coding  WAIS-III Digit symbol coding  WAIS-III Digit symbol coding  WAIS-III Digit symbol coding  WAIS-III Digit symbol coding  WAIS-III Digit symbol coding  WAIS-III Digit symbol coding  WAIS-III Digit symbol coding  WAIS-III Digit symbol coding  WAIS-III Digit symbol coding  WAIS-III Digit symbol coding  WAIS-III Symbol Search  WAIS-III Symbol Search  WAIS-III Symbol Search  WAIS-III Symbol Search  WAIS-III Symbol Search  WAIS-III Symbol Search  WAIS-III Symbol Search  WAIS-III Symbol Search  WAIS-III Symbol Search  WAIS-III Symbol Search  WAIS-III Symbol Search  WAIS-III Symbol Search  WAIS-III Symbol Search  WAIS-III Symbol Search | Volume | r = 0.219 (ns)  r = 0.207 (ns)  r = 0.207 (ns)  r = 0.093 (ns)  r = 0.144 (ns)  r = 0.247  r = 0.086 (ns)  r = 0.21 (ns)  r = 0.022 (ns)  r = -0.052 (ns)  r = 0.119 (ns)  r = 0.139 (ns)  r = -0.031 (ns)  r = 0.001 (ns)  r = 0.252  r = 0.252  r = 0.166 (ns)  r = 0.173 (ns)  r = 0.1 (ns)  r = 0.261  r = 0.16 (ns)  r = 0.244  r = 0.014 (ns)  r = -0.014 (ns)  r = 0.114 (ns)  r = 0.128 (ns)  r = -0.048 (ns)  r = 0.007 (ns)  r = 0.187 (ns)  r = 0.149 (ns)  r = 0.166 (ns)  r = 0.033 (ns)  r = 0.094 (ns)  r = 0.214 (ns)  r = 0.128 (ns)  r = 0.161 (ns)  r = 0.093 (ns)  r = 0.002 (ns)  r = 0.111 (ns)  r = 0.119 (ns)  r = -0.003 (ns)  r = -0.053 (ns)  r = 0.201 (ns)  r = 0.166 (ns)  r = 0.165 (ns)  r = 0.126 (ns)  r = 0.135 (ns)  r = 0.218 (ns)  r = 0.05 (ns)  r = 0.243 (ns)  r = 0.002 (ns)  r = -0.047 (ns)  r = 0.068 (ns)  r = 0.12 (ns)  r = -0.022 (ns)  r = 0.037 (ns)  r = 0.155 (ns)  r = 0.192 (ns)  r = 0.198 (ns)  r = 0.09 (ns)  r = 0.109 (ns)  r = 0.162 (ns)  r = 0.031 (ns)  r = 0.14 (ns)  r = -0.051 (ns)  r = -0.094 (ns)  r = 0.104 (ns)  r = 0.109 (ns)  r = -0.058 (ns)  r = -0.004 (ns)  r = 0.212 (ns)  r = 0.263  r = 0.137 (ns)  r = 0.171 (ns)  r = 0.073 (ns)  r = 0.132 (ns)  r = 0.086 (ns)  r = 0.172 (ns)  r = 0.061 (ns)  r = 0.009 (ns)  r = 0.095 (ns)  r = 0.091 (ns)  r = -0.101 (ns)  r = -0.125 (ns)  r = 0.228 (ns)  r = 0.22 (ns)  r = 0.209 (ns)  r = 0.116 (ns)  r = 0.084 (ns)  r = 0.214 (ns)  r = 0.117 (ns)  r = 0.163 (ns)  r = 0.08 (ns)  r = 0.068 (ns)  r = 0.105 (ns)  r = 0.16 (ns)  r = -0.083 (ns)  r = -0.036 (ns)  r = 0.211 (ns)  r = 0.221 (ns)  r = 0.177 (ns)  r = 0.185 (ns)  r = 0.051 (ns)  r = 0.219 (ns)  r = 0.179 (ns)  r = 0.266  r = 0.136 (ns)  r = 0.096 (ns)  r = 0.168 (ns)  r = 0.162 (ns)  r = -0.003 (ns)  r = 0.055 (ns)  r = 0.245  r = 0.243 (ns)  r = 0.122 (ns)  r = 0.131 (ns)  r = 0.123 (ns)  r = 0.256  r = 0.121 (ns)  r = 0.188 (ns)  r = -0.114 (ns)  r = -0.126 (ns)  r = 0.046 (ns)  r = 0.068 (ns)  r = -0.088 (ns)  r = -0.034 (ns) | 163 |
| Koshiyama et al. (2018b) | Left hippocampus  Right hippocampus  Left amygdala  Right amygdala  Left thalamus  Right thalamus  Left Accumbens  Right Accumbens  Left Caudate  Right Caudate  Left Putamen  Right Putamen  Left Pallidum  Right Pallidum  Left hippocampus  Right hippocampus  Left amygdala  Right amygdala  Left thalamus  Right thalamus  Left Accumbens  Right Accumbens  Left Caudate  Right Caudate  Left Putamen  Right Putamen  Left Pallidum  Right Pallidum  Left hippocampus  Right hippocampus  Left amygdala  Right amygdala  Left thalamus  Right thalamus  Left Accumbens  Right Accumbens  Left Caudate  Right Caudate  Left Putamen  Right Putamen  Left Pallidum  Right Pallidum  Left hippocampus  Right hippocampus  Left amygdala  Right amygdala  Left thalamus  Right thalamus  Left Accumbens  Right Accumbens  Left Caudate  Right Caudate  Left Putamen  Right Putamen  Left Pallidum  Right Pallidum | VM  VisM  VM/VisM  ATT | WMS - R - Verbal immediate recall  WMS - R - Verbal immediate recall  WMS - R - Verbal immediate recall  WMS - R - Verbal immediate recall  WMS - R - Verbal immediate recall  WMS - R - Verbal immediate recall  WMS - R - Verbal immediate recall  WMS - R - Verbal immediate recall  WMS - R - Verbal immediate recall  WMS - R - Verbal immediate recall  WMS - R - Verbal immediate recall  WMS - R - Verbal immediate recall  WMS - R - Verbal immediate recall  WMS - R - Verbal immediate recall  WMS - R - Visual immediate recall  WMS - R - Visual immediate recall  WMS - R - Visual immediate recall  WMS - R - Visual immediate recall  WMS - R - Visual immediate recall  WMS - R - Visual immediate recall  WMS - R - Visual immediate recall  WMS - R - Visual immediate recall  WMS - R - Visual immediate recall  WMS - R - Visual immediate recall  WMS - R - Visual immediate recall  WMS - R - Visual immediate recall  WMS - R - Visual immediate recall  WMS - R - Visual immediate recall  WMS - R - Delayed recall  WMS - R - Delayed recall  WMS - R - Delayed recall  WMS - R - Delayed recall  WMS - R - Delayed recall  WMS - R - Delayed recall  WMS - R - Delayed recall  WMS - R - Delayed recall  WMS - R - Delayed recall  WMS - R - Delayed recall  WMS - R - Delayed recall  WMS - R - Delayed recall  WMS - R - Delayed recall  WMS - R - Delayed recall  WMS - R - Attention/Concentration  WMS - R - Attention/Concentration  WMS - R - Attention/Concentration  WMS - R - Attention/Concentration  WMS - R - Attention/Concentration  WMS - R - Attention/Concentration  WMS - R - Attention/Concentration  WMS - R - Attention/Concentration  WMS - R - Attention/Concentration  WMS - R - Attention/Concentration  WMS - R - Attention/Concentration  WMS - R - Attention/Concentration  WMS - R - Attention/Concentration  WMS - R - Attention/Concentration | Volume | r = 0.339  r = 0.281  r = 0.118 (ns)  r = 0.142 (ns)  r = 0.145 (ns)  r = 0.177 (ns)  r = 0.244  r = 0.246  r = 0.118 (ns)  r = 0.095 (ns)  r = 0.127 (ns)  r = 0.089 (ns)  r = −0.060 (ns)  r = 0.02 (ns)  r = 0.361  r = 0.41  r = 0.196 (ns)  r = 0.079 (ns)  r = 0.236 (ns)  r = 0.27  r = 0.17 (ns)  r = 0.192 (ns)  r = 0.033 (ns)  r = 0.005 (ns)  r = 0.039 (ns)  r = 0.013 (ns)  r = −0.161 (ns)  r = −0.012 (ns)  r = 0.333  r = 0.31  r = 0.099 (ns)  r = 0.084 (ns)  r = 0.17 (ns)  r = 0.194 (ns)  r = 0.255  r = 0.27  r = 0.031 (ns)  r = −0.002 (ns)  r = 0.075 (ns)  r = 0.041 (ns)  r = −0.120 (ns)  r = 0.002 (ns)  r = 0.252  r = 0.229  r = 0.123 (ns)  r = 0.076 (ns)  r = 0.093 (ns)  r = 0.163 (ns)  r = 0.088 (ns)  r = 0.231  r = −0.013 (ns)  r = −0.034 (ns)  r = 0.082 (ns)  r = 0.035 (ns)  r = −0.033 (ns)  r = 0.08 (ns) | 174 |
| Krabbendam et al. (2000) | left parahippocampal gyrus | ATT | Stroop Color-Word Test (SCWT) interferences | Volume | r = –0.57 | 27 |
| Kumarasinghe et al. (2014) | left ventral temporal cortex  left ectorhinal  left temporopolar areas (BA38)  right ventral temporal cortex  right ectorhinal (BA36)  right temporopolar areas (BA38)  right ventro-temporal cortex  right dorsolateral prefrontal cortex | SP  VF  WM | RBANS - figure copy  RBANS - figure copy  RBANS - figure copy  RBANS - figure copy  RBANS - figure copy  RBANS - figure copy  RBANS - semantic fluency  RBANS - digit span | Volume | r = 0.567  r = 0.608  r = 0.5  r = 0.644  r = 0.397  r = 0.611  r = 0.59  r = 0.64 | 18 |
| Lee et al. (2007) | vermis grey matter  vermis white matter  left ant.cerebellum grey matter  left ant. cerebellum white matter  right ant. cerebellum grey matter  right ant.cerebellum white matter  left post. cerebellum grey matter  left post. cerebellum white matter  right post.cerebellum grey matter  right post.cerebellum white matter  vermis grey matter  vermis white matter  left ant.cerebellum grey matter  left ant. cerebellum white matter  right ant. cerebellum grey matter  right ant.cerebellum white matter  left post. cerebellum grey matter  left post.cerebellum white matter  right post.cerebellum grey matter  right post.cerebellum white matter  vermis grey matter  vermis white matter  left ant. cerebellum grey matter  left ant. cerebellum white matter  right ant. cerebellum grey matter  right ant.cerebellum white matter  left post. cerebellum grey matter  left post. cerebellum white matter  right post. cerebellum grey matter  right post.cerebellum white matter  vermis grey matter  vermis white matter  left ant. cerebellum grey matter  left ant. cerebellum white matter  right ant. cerebellum grey matter  right ant. cerebellum white matter  left post. cerebellum grey matter  left post. cerebellum white matter  right post. cerebellum grey matter  right post.cerebellum white matter  vermis grey matter  vermis white matter  left ant. cerebellum grey matter  left ant. cerebellum white matter  right ant. cerebellum grey matter  right ant. cerebellum white matter  left post. cerebellum grey matter  left post. cerebellum white matter  right post. cerebellum grey matter  right post.cerebellum white matter  vermis grey matter  vermis white matter  left ant. cerebellum grey matter  left ant. cerebellum white matter  right ant. cerebellum grey matter  right ant. cerebellum white matter  left post. cerebellum grey matter  left post. cerebellum white matter  right post. cerebellum grey matter  right post.cerebellum white matter  vermis grey matter  vermis white matter  left ant. cerebellum grey matter  left ant. cerebellum white matter  right ant. cerebellum grey matter  right ant. cerebellum white matter  left post. cerebellum grey matter  left post. cerebellum white matter  right post. cerebellum grey matter  right post.cerebellum white matter  vermis grey matter  vermis white matter  left ant. cerebellum grey matter  left ant. cerebellum white matter  right ant. cerebellum grey matter  right ant. cerebellum white matter  left post. cerebellum grey matter  left post. cerebellum white matter  right post. cerebellum grey matter  right post.cerebellum white matter  vermis grey matter  vermis white matter  left ant. cerebellum grey matter  left ant. cerebellum white matter  right ant. cerebellum grey matter  right ant. cerebellum white matter  left post. cerebellum grey matter  left post. cerebellum white matter  right post. cerebellum grey matter  right post.cerebellum white matter  vermis grey matter  vermis white matter  left ant. cerebellum grey matter  left ant. cerebellum white matter  right ant. cerebellum grey matter  right ant. cerebellum white matter  left post. cerebellum grey matter  left post. cerebellum white matter  right post. cerebellum grey matter  right post.cerebellum white matter | ATT  VM  VF  R&EF | Auditory CPT - total correct  Auditory CPT - total correct  Auditory CPT - total correct  Auditory CPT - total correct  Auditory CPT - total correct  Auditory CPT - total correct  Auditory CPT - total correct  Auditory CPT - total correct  Auditory CPT - total correct  Auditory CPT - total correct  Auditory CPT - false positives  Auditory CPT - false positives  Auditory CPT - false positives  Auditory CPT - false positives  Auditory CPT - false positives  Auditory CPT - false positives  Auditory CPT - false positives  Auditory CPT - false positives  Auditory CPT - false positives  Auditory CPT - false positives  HVLT-R - total correct recall  HVLT-R - total correct recall  HVLT-R - total correct recall  HVLT-R - total correct recall  HVLT-R - total correct recall  HVLT-R - total correct recall  HVLT-R - total correct recall  HVLT-R - total correct recall  HVLT-R - total correct recall  HVLT-R - total correct recall  HVLT-R - total delayed recall  HVLT-R - total delayed recall  HVLT-R - total delayed recall  HVLT-R - total delayed recall  HVLT-R - total delayed recall  HVLT-R - total delayed recall  HVLT-R - total delayed recall  HVLT-R - total delayed recall  HVLT-R - total delayed recall  HVLT-R - total delayed recall  Delis–Kaplan verbal fluency test - letter fluency  Delis–Kaplan verbal fluency test - letter fluency  Delis–Kaplan verbal fluency test - letter fluency  Delis–Kaplan verbal fluency test - letter fluency  Delis–Kaplan verbal fluency test - letter fluency  Delis–Kaplan verbal fluency test - letter fluency  Delis–Kaplan verbal fluency test - letter fluency  Delis–Kaplan verbal fluency test - letter fluency  Delis–Kaplan verbal fluency test - letter fluency  Delis–Kaplan verbal fluency test - letter fluency  Delis–Kaplan verbal fluency test - category fluency  Delis–Kaplan verbal fluency test - category fluency  Delis–Kaplan verbal fluency test - category fluency  Delis–Kaplan verbal fluency test - category fluency  Delis–Kaplan verbal fluency test - category fluency  Delis–Kaplan verbal fluency test - category fluency  Delis–Kaplan verbal fluency test - category fluency  Delis–Kaplan verbal fluency test - category fluency  Delis–Kaplan verbal fluency test - category fluency  Delis–Kaplan verbal fluency test - category fluency  Delis–Kaplan verbal fluency test - repetition error  Delis–Kaplan verbal fluency test - repetition error  Delis–Kaplan verbal fluency test - repetition error  Delis–Kaplan verbal fluency test - repetition error  Delis–Kaplan verbal fluency test - repetition error  Delis–Kaplan verbal fluency test - repetition error  Delis–Kaplan verbal fluency test - repetition error  Delis–Kaplan verbal fluency test - repetition error  Delis–Kaplan verbal fluency test - repetition error  Delis–Kaplan verbal fluency test - repetition error  Wisconsin card sorting test (WCST) - total correct  Wisconsin card sorting test (WCST) - total correct  Wisconsin card sorting test (WCST) - total correct  Wisconsin card sorting test (WCST) - total correct  Wisconsin card sorting test (WCST) - total correct  Wisconsin card sorting test (WCST) - total correct  Wisconsin card sorting test (WCST) - total correct  Wisconsin card sorting test (WCST) - total correct  Wisconsin card sorting test (WCST) - total correct  Wisconsin card sorting test (WCST) - total correct  WCST - total perseverative errors  WCST - total perseverative errors  WCST - total perseverative errors  WCST - total perseverative errors  WCST - total perseverative errors  WCST - total perseverative errors  WCST - total perseverative errors  WCST - total perseverative errors  WCST - total perseverative errors  WCST - total perseverative errors  WCST - total categories completed  WCST - total categories completed  WCST - total categories completed  WCST - total categories completed  WCST - total categories completed  WCST - total categories completed  WCST - total categories completed  WCST - total categories completed  WCST - total categories completed  WCST - total categories completed | Volume | r = 0.29 (ns)  r = 0.04 (ns)  r = 0.29 (ns)  r = 0.1 (ns)  r = 0.31 (ns)  r = -0.03 (ns)  r = 0.2 (ns)  r = 0.1 (ns)  r = 0.39  r = 0.23 (ns)  r = -0.09 (ns)  r = 0.28 (ns)  r = -0.15 (ns)  r = 0.05 (ns)  r = -0.02 (ns)  r = 0.03 (ns)  r = -0.18 (ns)  r = 0.27 (ns)  r = -0.17 (ns)  r = 0.17 (ns)  r = 0.12 (ns)  r = -0.26 (ns)  r = 0.04 (ns)  r = -0.31 (ns)  r = 0.18 (ns)  r = -0.41  r = 0.13 (ns)  r = -0.29 (ns)  r = 0.31 (ns)  r = -0.13 (ns)  r = 0.25 (ns)  r = -0.15 (ns)  r = 0.08 (ns)  r = -0.15 (ns)  r = 0.2 (ns)  r = -0.37  r = 0.21 (ns)  r = -0.11 (ns)  r = 0.33  r = -0.03 (ns)  r = 0.15 (ns)  r = 0.13 (ns)  r = 0.2 (ns)  r = 0.13 (ns)  r = 0.31 (ns)  r = 0.05 (ns)  r = 0.00 (ns)  r = 0.39  r = 0.14 (ns)  r = 0.38  r = 0.3 (ns)  r = -0.18 (ns)  r = 0.21 (ns)  r = 0.07 (ns)  r = 0.25 (ns)  r = 0.02 (ns)  r = 0.24 (ns)  r = 0.04 (ns)  r = 0.35  r = 0.12 (ns)  r = 0.15 (ns)  r = 0.51  r = 0.26 (ns)  r = 0.43  r = 0.21 (ns)  r = 0.10 (ns)  r = -0.07 (ns)  r = 0.51  r = -0.05 (ns)  r = 0.41  r = 0.21 (ns)  r = -0.12 (ns)  r = 0.08 (ns)  r = -0.06 (ns)  r = 0.24 (ns)  r = 0.2 (ns)  r = 0.22 (ns)  r = -0.19 (ns)  r = 0.13 (ns)  r = -0.03 (ns)  r = -0.15* (ns)  r = 0.11* (ns)  r = -0.09* (ns)  r = 0.04* (ns)  r = -0.23* (ns)  r = -0.15* (ns)  r = -0.11* (ns)  r = 0.06* (ns)  r = -0.02* (ns)  r = -0.11* (ns)  r = 0.26 (ns)  r = -0.07 (ns)  r = 0.13 (ns)  r = -0.05 (ns)  r = 0.28 (ns)  r = 0.27 (ns)  r = 0.19 (ns)  r = -0.13 (ns)  r = 0.2 (ns)  r = 0.01 (ns) | 40 |
| Levitt et al. (1999) | vermis white matter | VM | WMS - Logical memory - immediate | Volume | r = 0.54 | 15 |
| Levitt et al. (2013) | left precommissural caudate  right precommissural caudate  left precommissural caudate  left postcommissural putamen  left precommissural caudate  left postcommissural caudate  right postcommissural caudate  left postcommissural putamen | R&EF | Wisconsin Card Sorting Test - categories  Wisconsin Card Sorting Test - categories  WCST - perseverative responses  WCST - perseverative responses  Wisconsin Card Sorting Test - perseverative errors  Trail making test B  Trail making test B  Trail making test B | Volume | ρ = 0.55  ρ = 0.57  ρ = -0.44*  ρ = -0.39*  ρ = -0.43*  ρ = -0.44*  ρ = -0.55*  ρ = 0.41* | 26  27 |
| Maat, van Haren, Bartholomeusz, Kahn, and Cahn (2016) | inferior PFC  pars triangularis | SC | Degraded facial affect recognition task - anger  Degraded facial affect recognition task - anger | Volume | t = 2.19  t = 2.29 | 166 |
| Madre et al. (2016) | *Review* |  | *Review* |  | *Review* |  |
| Maher, Manschreck, Woods, Yurgelun-Todd, and Tsuang (1995) | whole brain  total frontal  total temporal  total temporal horns  total corpus striatum  total ventral pallidum  total lateral ventricles  whole brain  total frontal  total temporal  total temporal horns  total corpus striatum  total ventral pallidum  total lateral ventricles | VM | Verbal Recall List 1 + 2  Verbal Recall List 1 + 2  Verbal Recall List 1 + 2  Verbal Recall List 1 + 2  Verbal Recall List 1 + 2  Verbal Recall List 1 + 2  Verbal Recall List 1 + 2  Verbal Recall Lists 3 + 4  Verbal Recall Lists 3 + 4  Verbal Recall Lists 3 + 4  Verbal Recall Lists 3 + 4  Verbal Recall Lists 3 + 4  Verbal Recall Lists 3 + 4  Verbal Recall Lists 3 + 4 | Volume | r = -0.176  r = 0.031  r = -0.059  r = 0.089  r = -0.116  r = -0.173  r = -0.07  r = -0.042  r = 0.311  r = -0.149  r = -0.116  r = -0.397  r = -0.031  r = -0.163 | 18 |
| Mathew et al. (2014) | *Some missing results* |  |  |  |  |  |
| Matsui et al. (2008) | *Some results missing* |  |  |  |  |  |
| Massey et al. (2017) | left medial prefrontal  right medial prefrontal  left inferior frontal  right inferior frontal  left anterior-mid cingulate  right anterior-mid cingulate  left insula  right insula  left supplementary motor  right supplementary motor  left temporo-parietal junction  right temporo-parietal junction  left precuneus  right precuneus | SC | Emotional Perspective-Taking Task (EPT)  Emotional Perspective-Taking Task (EPT)  Emotional Perspective-Taking Task (EPT)  Emotional Perspective-Taking Task (EPT)  Emotional Perspective-Taking Task (EPT)  Emotional Perspective-Taking Task (EPT)  Emotional Perspective-Taking Task (EPT)  Emotional Perspective-Taking Task (EPT)  Emotional Perspective-Taking Task (EPT)  Emotional Perspective-Taking Task (EPT)  Emotional Perspective-Taking Task (EPT)  Emotional Perspective-Taking Task (EPT)  Emotional Perspective-Taking Task (EPT)  Emotional Perspective-Taking Task (EPT) | Cortical Thickness | r = 0.00 (ns)  r = -0.09 (ns)  r = 0.14 (ns)  r = 0.11 (ns)  r = -0.12 (ns)  r = -0.19 (ns)  r = 0.20 (ns)  r = -0.05 (ns)  r = 0.13 (ns)  r = 0.14 (ns)  r = 0.06 (ns)  r = 0.12 (ns)  r = 0.04 (ns)  r = 0.16 (ns) | 39 |
| McCarley, Shenton, O'Donnell, and Nestor (1993) | *Review* |  | *Review* |  | *Review* |  |
| McIntosh et al. (2009) | *Some results missing* |  |  |  |  |  |
| McKenna, Miles, Babb, Goff, and Lazar (2019) | frontal lobe  right lateral orbitofrontal  left lateral orbitofrontal  left medial orbital frontal  right pars opercularis  left pars opercularis  left pars triangularis  right precentral  left precentral  right rostral middle frontal  left rostral middle frontal  left superior frontal  parietal lobe  right inferior parietal  left inferior parietal  right precuneus  left precuneus  temporal lobe  left BANKSSTS  right fusiform  left fusiform  right inferior temporal  left inferior temporal  right middle temporal  left middle temporal  right superior temporal  right transverse temporal  left caudal anterior cingulate  left rostral anterior cingulate  frontal lobe  right lateral orbitofrontal  left lateral orbitofrontal  right medial orbital frontal  left medial orbital frontal  left pars opercularis  right precentral  left precentral  right rostral middle frontal  left rostral middle frontal  right superior frontal  left superior frontal  parietal lobe  right inferior parietal  left inferior parietal  temporal lobe  left BANKSSTS  left inferior temporal  right middle temporal  left middle temporal  right superior temporal  left superior temporal  right transverse temporal  right cuneus  right insula  left insula  left posterior cingulate  right rostral anterior cingulate  left rostral anterior cingulate  frontal lobe  left lateral orbital frontal  right superior frontal  left superior frontal  parietal lobe  left inferior parietal  right superior parietal  left superior parietal  left supramarginal  temporal lobe  left BANKSSTS  left entorhinal  right fusiform  left fusiform  left inferior temporal  right parahippocampal  occipital lobe  right lateral occipital  left lateral occipital  right lingual  left lingual  right insula  left insula  right posterior cingulate  occipital lobe  left lateral occipital  right lingual  left lingual  frontal lobe  left lateral orbitofrontal  left pars opercularis  left precentral  right rostral middle frontal  left rostral middle frontal  left inferior parietal  left precuneus  left supramarginal  temporal lobe  right BANKSSTS  left entorhinal  left fusiform  left inferior temporal  right parahippocampal  right transverse temporal  right lingual  right insula  temporal lobe  right BANKSSTS  left fusiform  right middle temporal  left middle temporal  right superior temporal  left superior temporal  right transverse temporal  frontal lobe  right caudal middle frontal  left lateral orbital frontal  left precentral  left rostral middle frontal  left superior frontal  right superior parietal  right BANKSSTS  left inferior temporal  right transverse temporal  occipital lobe  right lingual  frontal lobe  right medial orbital frontal  right rostral middle frontal  left rostral middle frontal  parietal lobe  right inferior parietal  temporal lobe  right BANKSSTS  left fusiform  left inferior temporal  right middle temporal  left middle temporal  right superior temporal  left superior temporal  right transverse temporal  right cuneus  left lateral occipital  left lingual  right insula  left precentral  right superior frontal  left superior frontal  left inferior parietal  left superior parietal  left supramarginal  right BANKSSTS  left BANKSSTS  left entorhinal  left fusiform  right lateral occipital  left lateral occipital  right lingual  left lingual  right insula  right posterior cingulate  right superior parietal  left supramarginal  left fusiform  occipital lobe  left lateral occipital  right lingual  left lingual  right pars opercularis  left precentral  left BANKSSTS  right precentral  right superior temporal  left superior temporal  right transverse temporal  left rostral anterior cingulate | R&EF | Wisconsin Card Sorting Test - perseverative errors  Wisconsin Card Sorting Test - perseverative errors  Wisconsin Card Sorting Test - perseverative errors  Wisconsin Card Sorting Test - perseverative errors  Wisconsin Card Sorting Test - perseverative errors  Wisconsin Card Sorting Test - perseverative errors  Wisconsin Card Sorting Test - perseverative errors  Wisconsin Card Sorting Test - perseverative errors  Wisconsin Card Sorting Test - perseverative errors  Wisconsin Card Sorting Test - perseverative errors  Wisconsin Card Sorting Test - perseverative errors  Wisconsin Card Sorting Test - perseverative errors  Wisconsin Card Sorting Test - perseverative errors  Wisconsin Card Sorting Test - perseverative errors  Wisconsin Card Sorting Test - perseverative errors  Wisconsin Card Sorting Test - perseverative errors  Wisconsin Card Sorting Test - perseverative errors  Wisconsin Card Sorting Test - perseverative errors  Wisconsin Card Sorting Test - perseverative errors  Wisconsin Card Sorting Test - perseverative errors  Wisconsin Card Sorting Test - perseverative errors  Wisconsin Card Sorting Test - perseverative errors  Wisconsin Card Sorting Test - perseverative errors  Wisconsin Card Sorting Test - perseverative errors  Wisconsin Card Sorting Test - perseverative errors Wisconsin Card Sorting Test - perseverative errors  Wisconsin Card Sorting Test - perseverative errors  Wisconsin Card Sorting Test - perseverative errors  Wisconsin Card Sorting Test - perseverative errors  Wisconsin Card Sorting Test - perseverative errors  Wisconsin Card Sorting Test - perseverative errors  Wisconsin Card Sorting Test - perseverative errors  Wisconsin Card Sorting Test - perseverative errors  Wisconsin Card Sorting Test - perseverative errors  Wisconsin Card Sorting Test - perseverative errors  Wisconsin Card Sorting Test - perseverative errors  Wisconsin Card Sorting Test - perseverative errors  Wisconsin Card Sorting Test - perseverative errors  Wisconsin Card Sorting Test - perseverative errors  Wisconsin Card Sorting Test - perseverative errors  Wisconsin Card Sorting Test - perseverative errors  Wisconsin Card Sorting Test - perseverative errors  Wisconsin Card Sorting Test - perseverative errors  Wisconsin Card Sorting Test - perseverative errors  Wisconsin Card Sorting Test - perseverative errors  Wisconsin Card Sorting Test - perseverative errors  Wisconsin Card Sorting Test - perseverative errors  Wisconsin Card Sorting Test - perseverative errors  Wisconsin Card Sorting Test - perseverative errors  Wisconsin Card Sorting Test - perseverative errors  Wisconsin Card Sorting Test - perseverative errors  Wisconsin Card Sorting Test - perseverative errors  Wisconsin Card Sorting Test - perseverative errors  Wisconsin Card Sorting Test - perseverative errors  Wisconsin Card Sorting Test - perseverative errors  Wisconsin Card Sorting Test - perseverative errors  Wisconsin Card Sorting Test - perseverative errors  Wisconsin Card Sorting Test - perseverative errors  Wisconsin Card Sorting Test - non perseverative errors  Wisconsin Card Sorting Test - non perseverative errors  Wisconsin Card Sorting Test - non perseverative errors  Wisconsin Card Sorting Test - non perseverative errors  Wisconsin Card Sorting Test - non perseverative errors  Wisconsin Card Sorting Test - non perseverative errors  Wisconsin Card Sorting Test - non perseverative errors  Wisconsin Card Sorting Test - non perseverative errors  Wisconsin Card Sorting Test - non perseverative errors  Wisconsin Card Sorting Test - non perseverative errors Wisconsin Card Sorting Test - non perseverative errors  Wisconsin Card Sorting Test - non perseverative errors  Wisconsin Card Sorting Test - non perseverative errors  Wisconsin Card Sorting Test - non perseverative errors  Wisconsin Card Sorting Test - non perseverative errors  Wisconsin Card Sorting Test - non perseverative errors  Wisconsin Card Sorting Test - non perseverative errors  Wisconsin Card Sorting Test - non perseverative errors  Wisconsin Card Sorting Test - non perseverative errors  Wisconsin Card Sorting Test - non perseverative errors  Wisconsin Card Sorting Test - non perseverative errors  Wisconsin Card Sorting Test - non perseverative errors  Wisconsin Card Sorting Test - non perseverative errors  Wisconsin Card Sorting Test - non perseverative errors  Wisconsin Card Sorting Test - non perseverative errors  Wisconsin Card Sorting Test - non perseverative errors  Wisconsin Card Sorting Test - non perseverative errors  Wisconsin Card Sorting Test - non perseverative errors  Wisconsin Card Sorting Test - categories completed  Wisconsin Card Sorting Test - categories completed  Wisconsin Card Sorting Test - categories completed  Wisconsin Card Sorting Test - categories completed  Wisconsin Card Sorting Test - categories completed  Wisconsin Card Sorting Test - categories completed  Wisconsin Card Sorting Test - categories completed  Wisconsin Card Sorting Test - categories completed  Wisconsin Card Sorting Test - categories completed  Wisconsin Card Sorting Test - categories completed  Wisconsin Card Sorting Test - categories completed  Wisconsin Card Sorting Test - categories completed  Wisconsin Card Sorting Test - categories completed  Wisconsin Card Sorting Test - categories completed  Wisconsin Card Sorting Test - categories completed  Wisconsin Card Sorting Test - categories completed  Wisconsin Card Sorting Test - categories completed  Wisconsin Card Sorting Test - categories completed  Wisconsin Card Sorting Test - categories completed  Wisconsin Card Sorting Test - categories completed  Wisconsin Card Sorting Test - categories completed  Wisconsin Card Sorting Test - categories completed  Wisconsin Card Sorting Test - categories completed  Wisconsin Card Sorting Test - categories completed  Wisconsin Card Sorting Test - categories completed  Wisconsin Card Sorting Test - categories completed  Wisconsin Card Sorting Test - concept level responses  Wisconsin Card Sorting Test - concept level responses  Wisconsin Card Sorting Test - concept level responses  Wisconsin Card Sorting Test - concept level responses  Wisconsin Card Sorting Test - concept level responses  Wisconsin Card Sorting Test - concept level responses  Wisconsin Card Sorting Test - concept level responses  Wisconsin Card Sorting Test - concept level responses  Wisconsin Card Sorting Test - concept level responses  Wisconsin Card Sorting Test - concept level responses  Wisconsin Card Sorting Test - concept level responses  Wisconsin Card Sorting Test - concept level responses  Wisconsin Card Sorting Test - concept level responses  Wisconsin Card Sorting Test - concept level responses  Wisconsin Card Sorting Test - concept level responses  Wisconsin Card Sorting Test - concept level responses  Wisconsin Card Sorting Test - concept level responses  Wisconsin Card Sorting Test - concept level responses  Wisconsin Card Sorting Test - concept level responses  Wisconsin Card Sorting Test - concept level responses  Wisconsin Card Sorting Test - concept level responses  Wisconsin Card Sorting Test - concept level responses  Wisconsin Card Sorting Test - concept level responses  Wisconsin Card Sorting Test - concept level responses  Wisconsin Card Sorting Test - concept level responses  Wisconsin Card Sorting Test - concept level responses  Wisconsin Card Sorting Test - concept level responses  Wisconsin Card Sorting Test - concept level responses  Wisconsin Card Sorting Test - concept level responses  Wisconsin Card Sorting Test - concept level responses  Wisconsin Card Sorting Test - concept level responses  Wisconsin Card Sorting Test - trials to first category  Wisconsin Card Sorting Test - trials to first category  Wisconsin Card Sorting Test - trials to first category  Wisconsin Card Sorting Test - trials to first category  Wisconsin Card Sorting Test - trials to first category  Wisconsin Card Sorting Test - trials to first category  Wisconsin Card Sorting Test - trials to first category  Wisconsin Card Sorting Test - trials to first category  Wisconsin Card Sorting Test - trials to first category  Wisconsin Card Sorting Test - trials to first category  Wisconsin Card Sorting Test - trials to first category  Wisconsin Card Sorting Test - trials to first category  Wisconsin Card Sorting Test - trials to first category  Wisconsin Card Sorting Test - trials to first category  Wisconsin Card Sorting Test - trials to first category  Wisconsin Card Sorting Test - trials to first category  Wisconsin Card Sorting Test - trials to first category  Wisconsin Card Sorting Test - trials to first category  Wisconsin Card Sorting Test - trials to first category  Wisconsin Card Sorting Test - trials to first category  Wisconsin Card Sorting Test - trials to first category  Wisconsin Card Sorting Test - trials to first category  Wisconsin Card Sorting Test - trials to first category  Trail making test B  Trail making test B  Trail making test B  Trail making test B  Trail making test B  Trail making test B  Trail making test B  Trail making test B | Cortical thickness  Surface Area  Cortical thickness  Surface Area  Cortical Thickness  Surface Area  Cortical Thickness  Surface Area  Cortical Thickness  Surface Area  Cortical Thickness  Surface Area | ρ = -0.64*  ρ = -0.62*  ρ = -0.57*  ρ = -0.61*  ρ = -0.5*  ρ = -0.74*  ρ = -0.53*  ρ = -0.63*  ρ = -0.86*  ρ = -0.61*  ρ = -0.63*  ρ = -0.56*  ρ = -0.51*  ρ = -0.61*  ρ = -0.51*  ρ = -0.5*  ρ = -0.52*  ρ = -0.73*  ρ = -0.59*  ρ = -0.51*  ρ = -0.65*  ρ = -0.66*  ρ = -0.74*  ρ = -0.52*  ρ = -0.65*  ρ = -0.7*  ρ = -0.51*  ρ = -0.64*  ρ = -0.64*  ρ = -0.69*  ρ = -0.54*  ρ = -0.52*  ρ = -0.67*  ρ = -0.5*  ρ = -0.53*  ρ = -0.54*  ρ = -0.7*  ρ = -0.59*  ρ = -0.59*  ρ = -0.51*  ρ = -0.56*  ρ = -0.5*  ρ = -0.71*  ρ = -0.52*  ρ = -0.64*  ρ = -0.63*  ρ = -0.54*  ρ = -0.68*  ρ = -0.62*  ρ = -0.74*  ρ = -0.57*  r= -0.55*  ρ = -0.61*  ρ = -0.52*  ρ = -0.54*  ρ = -0.64*  ρ = -0.59*  ρ = -0.71*  r= -0.5*  ρ = -0.51*  ρ = -0.64*  ρ = -0.63*  ρ = -0.54*  ρ = -0.61*  r= -0.51*  r= -0.58*  ρ = -0.68*  ρ = -0.54*  ρ = -0.52*  ρ = -0.75*  ρ = -0.58*  ρ = -0.71*  ρ = -0.57*  ρ = -0.5*  ρ = -0.67*  ρ = -0.61*  ρ = -0.7*  ρ = -0.82*  ρ = -0.65*  ρ = -0.69*  ρ = -0.65*  ρ = -0.68*  ρ = -0.54*  ρ = -0.5*  r= -0.6 *  ρ = -0.58*  ρ =0.51  ρ =0.63  ρ =0.54  ρ =0.71  r= 0.5  ρ =0.59  r=0.54  r= 0.53  ρ =0.56  ρ =0.6  ρ =0.65  ρ =0.61  ρ =0.68  ρ =0.65  r= 0.52  ρ =0.65  ρ =0.62  ρ =0.57  r= 0.54  r= 0.66  ρ =0.54  ρ =0.52  r=0.53  r=0.5  r= 0.5  ρ =0.56  r= 0.5  r= 0.51  ρ =0.61  ρ =0.66  ρ =0.5  ρ =0.5  r= 0.54  ρ =0.52  ρ =0.57  ρ =0.58  ρ =0.74  r =0.53  r=0.56  r=0.53  r=0.53  r=0.52  r=0.56  r=0.56  r= 0.62  r= 0.51  ρ =0.53  r=0.54  r=0.66  r=0.56  r=0.61  r=0.52  r=0.62  r=0.59  r= 0.52  ρ =0.57  r= 0.54  r= -0.54*  ρ = -0.56*  ρ = -0.56*  ρ = -0.62*  ρ = -0.52*  ρ = -0.84*  r= -0.56*  ρ = -0.55*  ρ = -0.7*  ρ = -0.61*  ρ = -0.64*  ρ = -0.71*  ρ = -0.69*  ρ = -0.58*  r= -0.56*  ρ = -0.56*  ρ = -0.51*  r =-0.71*  r =-0.53*  ρ =-0.57*  ρ =-0.60*  ρ =-0.65*  ρ =-0.63*  r= -0.52*  r= -0.6*  r= -0.5*  r= -0.54*  r= -0.51*  ρ =-0.56*  r= -0.54*  ρ =-0.56* | 18 |
| Molina et al. (2009) | left DLPFC  right DLPFC  left DLPFC  right DLPFC  left DLPFC  right DLPFC  left DLPFC  right DLPFC  left DLPFC  right DLPFC  left DLPFC  right DLPFC  left DLPFC  right DLPFC  left DLPFC  right DLPFC  left DLPFC  right DLPFC  left DLPFC  right DLPFC  left DLPFC  right DLPFC  left DLPFC  right DLPFC | SP  WM  ATT  VM  VisM  R&EF  SP  WM  ATT  VM  VisM  R&EF | Neuropsychological tests (TMT A+WAIS-digit symbol)  Neuropsychological tests (TMT A+WAIS-digit symbol)  WAIS arithmetic  WAIS arithmetic  Toulouse-Pieron  Toulouse-Pieron  TAVEC short-term memory  TAVEC short-term memory  Rey accuracy and richness  Rey accuracy and richness  WCST (categories + perseverative errors)  WCST (categories + perseverative errors)  Neuropsychological tests (TMT A + WAIS-digit symbol)  Neuropsychological tests (TMT A + WAIS-digit symbol)  WAIS arithmetic  WAIS arithmetic  Toulouse-Pieron  Toulouse-Pieron  TAVEC short-term memory  TAVEC short-term memory  Rey accuracy and richness  Rey accuracy and richness  WCST (categories + perseverative errors)  WCST (categories + perseverative errors) | Volume | r= 0.179 (ns)  r= 0.157 (ns)  r= 0.402 (ns)  r= 0.474  r= 0.105 (ns)  r= − 0.229 (ns)  r= 0.326 (ns)  r= 0.459  r= 0.128 (ns)  r= − 0.091(ns)  r= 0.288 (ns)  r= 0.095 (ns)  ρ= 0.100 (ns)  ρ= 0.367 (ns)  ρ= 0.192 (ns)  ρ= 0.743  ρ= 0.383 (ns)  ρ= 0.283 (ns)  ρ= 0.034 (ns)  ρ= 0.604 (ns)  ρ= -0.192 (ns)  ρ= 0.494 (ns)  ρ= -0.317 (ns)  ρ= 0.133 (ns) | 22 |
| Namiki et al. (2007) | left amygdala right amygdala  left amygdala | SC | Pictures of Facial Affect series - Happy  Pictures of Facial Affect series - Happy  Pictures of Facial Affect series - Sadness | Volume | r= 0.59 r= 0.49 r= 0.45 | 20 |
| Nestor et al. (1993) | left parahippocampal gyrus  right parahippocampal gyrus  left posterior sup temporal gyrus  right posterior sup temporal gyrus  left posterior sup temporal gyrus | R&EF  VM | Wisconsin Card Sorting Test  Wisconsin Card Sorting Test  Wisconsin Card Sorting Test  Wisconsin Card Sorting Test  WMS paired association immediate | Volume | r=0.69  r=0.62  r=0.71  r=0.67  r=0.58 | 14    15 |
| Nestor, Onitsuka, et al. (2007) | right anterior fusiform gyrus  right posterior fusiform gyrus  left anterior fusiform gyrus  left posterior fusiform gyrus  right anterior sup temporal gyrus  right posterior sup temporal gyrus  left anterior sup temporal gyrus  left posterior sup temporal gyrus  right anterior fusiform gyrus  right posterior fusiform gyrus  left anterior fusiform gyrus  left posterior fusiform gyrus  right anterior sup temporal gyrus  right posterior sup temporal gyrus  left anterior sup temporal gyrus  left posterior sup temporal gyrus | R&EF  VisM | WCST: Non-perseverative errors  WCST: Non-perseverative errors  WCST: Non-perseverative errors  WCST: Non-perseverative errors  WCST: Non-perseverative errors  WCST: Non-perseverative errors  WCST: Non-perseverative errors  WCST: Non-perseverative errors  Wechsler Memory Scale-III: Facial memory  Wechsler Memory Scale-III: Facial memory  Wechsler Memory Scale-III: Facial memory  Wechsler Memory Scale-III: Facial memory  Wechsler Memory Scale-III: Facial memory  Wechsler Memory Scale-III: Facial memory  Wechsler Memory Scale-III: Facial memory  Wechsler Memory Scale-III: Facial memory | Volume | ρ= 0.248* (ns)  ρ= -0.148* (ns)  ρ= 0.421* (ns)  ρ= -0.201* (ns)  ρ= -0.25* (ns)  ρ= -0.492*  ρ= -0.066* (ns)  ρ= -0.432* (ns)  ρ= 0.543  ρ= 0.364  ρ= 0.579 (ns)  ρ= 0.354  ρ= -0.315 (ns)  ρ= 0.343 (ns)  ρ= -0.127 (ns)  ρ= -0.272 (ns) | 22 |
| Nestor, Kubicki, et al. (2007) | left hippocampus  right hippocampus  left hippocampus  right hippocampus  left hippocampus  right hippocampus  left hippocampus  right hippocampus  left hippocampus  right hippocampus | R&EF  VisM  VM | Wisconsin Card Sorting Test - categories  Wisconsin Card Sorting Test - categories  Wisconsin Card Sorting Test - perseverative errors  Wisconsin Card Sorting Test - perseverative errors  Wisconsin Card Sorting Test - non perseverative errors  Wisconsin Card Sorting Test - non perseverative errors  Doors and people test - visual  Doors and people test - visual  Doors and people test - verbal  Doors and people test - verbal | Volume | r= 0.066 (ns)  r= 0.107 (ns)  r= 0.073* (ns)  r= 0.026* (ns)  r= 0.001* (ns)  r= −0.248*  r= 0.215 (ns)  r= 0.47 (ns)  r= 0.602  r= 0.103 (ns) | 21  14 |
| Nestor et al. (2010) | left orbitofrontal cortex  right orbitofrontal cortex  left orbitofrontal cortex  right orbitofrontal cortex  left orbitofrontal cortex  right orbitofrontal cortex | WM  SP  R&EF | WAIS–III Working Memory  WAIS–III Working Memory  WAIS–III Processing Speed  WAIS–III Processing Speed  Iowa gambling task Block 1  Iowa gambling task Block 1 | Volume | r= -0.159 (ns)  r= -0.255 (ns)  r= -0.226 (ns)  r= -0.236 (ns)  r= 0.204 (ns)  r= -0.185 (ns) | 23  24 |
| Nestor et al. (2013) | left OFC  right OFC  left OFC  right OFC  left OFC  right OFC  left OFC  right OFC  left OFC  right OFC  left OFC  right OFC | WM  SP  VM  VisM  VM  VisM | WAIS-III - working memory  WAIS-III - working memory  WAIS-III - processing speed  WAIS-III - processing speed  WMS-III - auditory immediate  WMS-III - auditory immediate  WMS-III - visual immediate  WMS-III - visual immediate  WMS-III - auditory delayed  WMS-III - auditory delayed  WMS-III - visual delayed  WMS-III - visual delayed | Volume | r= −0.038 (ns)  r= −0.002 (ns)  r= −0.053 (ns)  r= −0.110 (ns)  r= −0.015 (ns)  r= 0.018 (ns)  r= −0.026 (ns)  r= 0.031 (ns)  r= −0.451  r= −0.542  r= −0.037 (ns)  r= 0.044 (ns) | 23 |
| Nestor et al. (2020) | left inferior frontal gyrus  right inferior frontal gyrus | VisM | WMS-III visual delayed  WMS-III visual delayed | Volume | r= 0.423  r= 0.401 | 27 |
| Ohtani et al. (2014) | left IFG relative volume | R&EF | Verbal fluency categories switching | Volume | r= 0.44 | 27 |
| Onitsuka et al. (2003) | left anterior Fusiform gyrus  right anterior Fusiform gyrus  left posterior Fusiform gyrus  right posterior Fusiform gyrus  left anterior Fusiform gyrus  right anterior Fusiform gyrus  left posterior Fusiform gyrus  right posterior Fusiform gyrus | VisM | WMS-III Immediate facial memory  WMS-III Immediate facial memory  WMS-III Immediate facial memory  WMS-III Immediate facial memory  WMS-III delayed facial memory  WMS-III delayed facial memory  WMS-III delayed facial memory  WMS-III delayed facial memory | Volume | ρ= -0.004 (ns)  ρ= 0.038 (ns)  ρ= -0.169 (ns)  ρ= -0.373 (ns)  ρ= 0.640  ρ= 0.658  ρ= 0.372 (ns)  ρ= 0.351 (ns) | 14 |
| Premkumar, Fannon, Kuipers, Cooke, et al. (2008) | dorsomedial prefrontal  dorsomedial prefrontal | WM  R&EF | Letter–Number Test  Stroop Test | Volume | r= 0.30  r= 0.38 | 64  62 |
| Premkumar, Fannon, Kuipers, Simmons, et al. (2008) | left thalamus  left superior temporal gyrus  right superior frontal gyrus  right inferior temporal gyrus  right dorsolateral prefrontal cortex  left superior temporal gyrus  right posterior cingulate gyrus | R&EF | Iowa Gambling Task - Attention to reward  Iowa Gambling Task - Attention to reward  Iowa Gambling Task Memory - past outcomes  Iowa Gambling Task Memory - past outcomes  Iowa Gambling Task - Impulsivity  Iowa Gambling Task - Impulsivity  Iowa Gambling Task - Impulsivity | Volume | r= 0.51  r= 0.36  r= −0.39  r= −0.40  r= 0.44  r= 0.39  r= 0.25 | 75 |
| Premkumar, Kumari, Corr, Fannon, and Sharma (2008) | temporal  prefrontal  prefrontal  hippocampus  hippocampus | R&EF  VisM  VF | WCST - perseverative errors  WMS Visual Reproductions immediate recall  WMS Visual Reproductions-delayed recall  WMS Visual Reproductions-delayed recall  Verbal fluency-letters (phonemic verbal fluency) | Volume | r= −0.26* (ns)  r= 0.29  r= 0.34  r= 0.07 (ns)  r= −0.003 (ns) | 42  54  52 |
| Pujol et al. (2014) | hippocampus  hippocampus | VM | RAVLT-total  RAVLT-Delayed | Volume | t= 1.52(ns)  t= 0.579(ns) | 51 |
| Rametti et al. (2007) | left anterior hippocampus  left hippocampus | VM | RAVLT - delayed recall  RAVLT - forgetting | Volume | r= 0.39  r= 0.54 | 28 |
| Ridler et al. (2006) | right inferior frontal gyrus  right posterolateral cerebellum  bilateral superior prefrontal cortex  bilateral medial cerebellum | R&EF | Adult executive function: AIM PC score  Adult executive function: AIM PC score  Adult executive function: AIM PC score  Adult executive function: AIM PC score | Volume | r= 0 (ns)  r= 0.03(ns)  r= 0.2 (ns)  r= 0.32 | 49 |
| Rodrigue et al. (2018) | *Canonical correlation analyses* |  | *not included in meta-analyses* |  |  |  |
| Rusch et al. (2008) | left amygdala  right amygdala  left hippocampus  right hippocampus  left DLPFC  right DLPFC  right amygdala  right amygdala  right amygdala | R&EF | Wisconsin Card Sorting Test - perseverative errors  Wisconsin Card Sorting Test - perseverative errors  Wisconsin Card Sorting Test - perseverative errors  Wisconsin Card Sorting Test - perseverative errors  Wisconsin Card Sorting Test - perseverative errors  Wisconsin Card Sorting Test - perseverative errors  Wisconsin Card Sorting Tests - categories completed  Wisconsin Card Sorting Tests - conceptual level responses  Wisconsin Card Sorting Tests - learning to learn index | Volume | r= −0.18* (ns)  r= −0.44*  r= −0.06* (ns)  r= 0.34* (ns)  r= −0.07* (ns)  r= 0.19* (ns)  r= 0.32  r= 0.31  r= 0.48 | 29 |
| Sachdev, Brodaty, Cheang, and Cathcart (2000) | right hippocampus  left hippocampus  right amygdala  left amygdala  right hippocampus  left hippocampus  right amygdala  left amygdala | VisM  VM | Visual memory (WMS: figural + visual reproduction 1 + 2)  Visual memory (WMS: figural + visual reproduction 1 + 2)  Visual memory (WMS: figural + visual reproduction 1 + 2) Visual memory (WMS: figural + visual reproduction 1 + 2)  Verbal memory (logical 1/2 + paired associates easy/hard)  Verbal memory (logical 1/2 + paired associates easy/hard)  Verbal memory (logical 1/2 + paired associates easy/hard)  Verbal memory (logical 1/2 + paired associates easy/hard) | Volume | r= 0.46  r= 0.26 (ns)  r= 0.6  r= 0.5  r= 0.56  r= 0.28 (ns)  r= 0.7  r= 0.61 | 20 |
| Sanfilipo et al. (2002) | left prefrontal  right prefrontal  left hippocampus  right hippocampus  left parahippocampus  right parahippocampus  left superior temporal gyrus  right superior temporal gyrus  left temporal  right temporal  left prefrontal white  right prefrontal white  left superior temporal white  right superior temporal white  left temporal white  right temporal white  left prefrontal  right prefrontal  left hippocampus  right hippocampus  left parahippocampus  right parahippocampus  left superior temporal gyrus  right superior temporal gyrus  left temporal  right temporal  left prefrontal white  right prefrontal white  left superior temporal white  right superior temporal white  left temporal white  right temporal white  left prefrontal  right prefrontal  left hippocampus  right hippocampus  left parahippocampus  right parahippocampus  left superior temporal gyrus  right superior temporal gyrus  left temporal  right temporal  left prefrontal white  right prefrontal white  left superior temporal white  right superior temporal white  left temporal white  right temporal white  left prefrontal  right prefrontal  left hippocampus  right hippocampus  left parahippocampus  right parahippocampus  left superior temporal gyrus  right superior temporal gyrus  left temporal  right temporal  left prefrontal white  right prefrontal white  left superior temporal white  right superior temporal white  left temporal white  right temporal white  left prefrontal  right prefrontal  left hippocampus  right hippocampus  left parahippocampus  right parahippocampus  left superior temporal gyrus  right superior temporal gyrus  left temporal  right temporal  left prefrontal white  right prefrontal white  left superior temporal white  right superior temporal white  left temporal white  right temporal white | R&EF  VM  VF  VisM  SP | Factor 2: cognitive flexibility  Factor 2: cognitive flexibility  Factor 2: cognitive flexibility  Factor 2: cognitive flexibility  Factor 2: cognitive flexibility  Factor 2: cognitive flexibility  Factor 2: cognitive flexibility  Factor 2: cognitive flexibility  Factor 2: cognitive flexibility  Factor 2: cognitive flexibility  Factor 2: cognitive flexibility  Factor 2: cognitive flexibility  Factor 2: cognitive flexibility  Factor 2: cognitive flexibility  Factor 2: cognitive flexibility  Factor 2: cognitive flexibility  Factor 3: word memory  Factor 3: word memory  Factor 3: word memory  Factor 3: word memory  Factor 3: word memory  Factor 3: word memory  Factor 3: word memory  Factor 3: word memory  Factor 3: word memory  Factor 3: word memory  Factor 3: word memory  Factor 3: word memory  Factor 3: word memory  Factor 3: word memory  Factor 3: word memory  Factor 3: word memory  Factor 4: verbal fluency  Factor 4: verbal fluency  Factor 4: verbal fluency  Factor 4: verbal fluency  Factor 4: verbal fluency  Factor 4: verbal fluency  Factor 4: verbal fluency  Factor 4: verbal fluency  Factor 4: verbal fluency  Factor 4: verbal fluency  Factor 4: verbal fluency  Factor 4: verbal fluency  Factor 4: verbal fluency  Factor 4: verbal fluency  Factor 4: verbal fluency  Factor 4: verbal fluency  Factor 5: visual memory  Factor 5: visual memory  Factor 5: visual memory  Factor 5: visual memory  Factor 5: visual memory  Factor 5: visual memory  Factor 5: visual memory  Factor 5: visual memory  Factor 5: visual memory  Factor 5: visual memory  Factor 5: visual memory  Factor 5: visual memory  Factor 5: visual memory  Factor 5: visual memory  Factor 5: visual memory  Factor 5: visual memory  WAIS-R Digit Symbol  WAIS-R Digit Symbol  WAIS-R Digit Symbol  WAIS-R Digit Symbol  WAIS-R Digit Symbol  WAIS-R Digit Symbol  WAIS-R Digit Symbol  WAIS-R Digit Symbol  WAIS-R Digit Symbol  WAIS-R Digit Symbol  WAIS-R Digit Symbol  WAIS-R Digit Symbol  WAIS-R Digit Symbol  WAIS-R Digit Symbol  WAIS-R Digit Symbol  WAIS-R Digit Symbol | Volume | r= -0.2 (ns)  r= -0.19 (ns)  r= 0.09 (ns)  r= 0.05 (ns)  r= 0.08 (ns)  r= 0.11 (ns)  r= -0.02 (ns)  r= -0.08 (ns)  r= 0.04 (ns)  r= -0.01 (ns)  r= 0.31  r= 0.27  r= 0.23 (ns)  r= 0.19 (ns)  r= 0.24 (ns)  r= 0.21 (ns)  r= 0.15 (ns)  r= 0.21 (ns)  r= 0.26  r= 0.4  r= 0.14 (ns)  r= 0.22 (ns)  r= 0.03 (ns)  r= 0.05 (ns)  r= 0.06 (ns)  r= 0.08 (ns)  r= 0.08 (ns)  r= -0.03 (ns)  r= 0.08 (ns)  r= -0.06 (ns)  r= 0.03 (ns)  r= -0.11 (ns)  r= -0.24 (ns)  r= -0.14 (ns)  r= -0.14 (ns)  r= -0.18 (ns)  r= -0.24 (ns)  r= -0.07 (ns)  r= -0.09 (ns)  r= -0.19 (ns)  r= -0.15 (ns)  r= -0.1 (ns)  r= 0.1 (ns)  r= 0.05 (ns)  r= -0.07 (ns)  r= -0.03 (ns)  r= -0.1 (ns)  r= -0.03 (ns)  r= 0.08 (ns)  r= 0.08 (ns)  r= 0.1 (ns)  r= 0.06 (ns)  r= 0.1 (ns)  r= -0.06 (ns)  r= -0.06 (ns)  r= 0.05 (ns)  r= -0.1 (ns)  r= -0.1 (ns)  r= -0.15 (ns)  r= -0.06 (ns)  r= -0.2 (ns)  r= 0.02 (ns)  r= -0.14 (ns)  r= -0.03 (ns)  r= 0.03 (ns)  r= 0.07 (ns)  r= 0.03 (ns)  r= -0.08 (ns)  r= -0.09 (ns)  r= -0.08 (ns)  r= -0.08 (ns)  r= -0.14 (ns)  r= -0.13 (ns)  r= -0.16 (ns)  r= 0.18 (ns)  r= 0.2 (ns)  r= 0.16 (ns)  r= 0.11 (ns)  r= 0.14 (ns)  r= 0.04 (ns) | 61 |
| Schobel et al. (2009) | Left anterior hippocampus  Left posterior hippocampus  Left orbitofrontal  Left dorsolateral prefrontal  Left anterior hippocampus  Left posterior hippocampus  Left orbitofrontal  Left dorsolateral prefrontal  Left anterior hippocampus  Left posterior hippocampus  Left orbitofrontal  Left dorsolateral prefrontal  Left anterior hippocampus  Left posterior hippocampus  Left orbitofrontal  Left dorsolateral prefrontal  Left anterior hippocampus  Left posterior hippocampus  Left orbitofrontal  Left dorsolateral prefrontal  Left anterior hippocampus  Left posterior hippocampus  Left orbitofrontal  Left dorsolateral prefrontal | WM  SP  R&EF | WAIS Working Memory Index  WAIS Working Memory Index  WAIS Working Memory Index  WAIS Working Memory Index  WAIS Processing Speed Index  WAIS Processing Speed Index  WAIS Processing Speed Index  WAIS Processing Speed Index  Wisconsin Card Sorting Test - Errors  Wisconsin Card Sorting Test - Errors  Wisconsin Card Sorting Test - Errors  Wisconsin Card Sorting Test - Errors  Wisconsin Card Sorting Test - Categories  Wisconsin Card Sorting Test - Categories  Wisconsin Card Sorting Test - Categories  Wisconsin Card Sorting Test - Categories  Trail-making Test B - seconds  Trail-making Test B - seconds  Trail-making Test B - seconds  Trail-making Test B - seconds  Trail-making Test B - errors  Trail-making Test B - errors  Trail-making Test B - errors  Trail-making Test B - errors | Volume | r= 0.33 (ns)  r= -0.02 (ns)  r= 0.44  r= -0.07 (ns)  r= 0.28 (ns)  r= 0.04 (ns)  r= 0.25 (ns)  r= -0.06 (ns)  r= -0.26* (ns)  r= -0.07* (ns)  r= -0.36*  r= -0.1* (ns)  r= 0.34(ns)  r= 0.15(ns)  r= 0.41  r= 0.08(ns)  r= -0.42*  r= -0.28* (ns)  r= -0.44*  r= -0.04* (ns)  r= -0.32* (ns)  r= 0.04* (ns)  r= -0.51*  r= -0.18* (ns) | 35 |
| Schretlen et al. (2010) | right cerebellar declive  left inferior occipital gyrus  left cingulate gyrus (BA 31)  left claustrum  right parahippocampal gyrus  right cingulate gyrus (BA31)  left cerebellar declive  right cerebellum  right lingual gyrus (BA18)  right anterior cingulate (BA32)  left fusiform gyrus (BA19) | R&EF | Trail making test B  Trail making test B  Trail making test B  Trail making test B  Trail making test B  Trail making test B  Trail making test B  Trail making test B  Trail making test B  Trail making test B  Trail making test B | Volume | t= 4.65  t= 4.28  t= 4.24  t= 4.06  t= 4.02  t= 4  t= 3.98  t= 3.79  t= 3.78  t= 3.77  t= 3.7 | 25 |
| Segarra et al. (2008) | bilateral hemispheres and vermis  bilateral hemispheres and vermis  right cerebellum  left cerebellum | WM  R&EF | WAIS-III letter number sequencing  WAIS-III arithmetic  Trail making test B-A  Trail making test B-A | Volume | r= 0.6  r= 0.72  r= -0.75  r= -0.7 | 28 |
| Seidman et al. (1994) | left dorsolateral  right dorsolateral  left orbitofrontal  right orbitofrontal  frontal  temporal  whole brain  left dorsolateral  right dorsolateral  left orbitofrontal  right orbitofrontal  frontal  temporal  whole brain  left dorsolateral  right dorsolateral  left orbitofrontal  right orbitofrontal  frontal  temporal  whole brain  left dorsolateral  right dorsolateral  left orbitofrontal  right orbitofrontal  frontal  temporal  whole brain  left dorsolateral  right dorsolateral  left orbitofrontal  right orbitofrontal  frontal  temporal  whole brain  left dorsolateral  right dorsolateral  left orbitofrontal  right orbitofrontal  frontal  temporal  whole brain  left dorsolateral  right dorsolateral  left orbitofrontal  right orbitofrontal  frontal  temporal  whole brain  left dorsolateral  right dorsolateral  left orbitofrontal  right orbitofrontal  frontal  temporal  whole brain  left dorsolateral  right dorsolateral  left orbitofrontal  right orbitofrontal  frontal  temporal  whole brain | R&EF  ATT  VM  VisM | Wisconsin Card Sorting Test - categories  Wisconsin Card Sorting Test - categories  Wisconsin Card Sorting Test - categories  Wisconsin Card Sorting Test - categories  Wisconsin Card Sorting Test - categories  Wisconsin Card Sorting Test - categories  Wisconsin Card Sorting Test - categories  Wisconsin Card Sorting Test - preservative responses  Wisconsin Card Sorting Test - preservative responses  Wisconsin Card Sorting Test - preservative responses  Wisconsin Card Sorting Test - preservative responses  Wisconsin Card Sorting Test - preservative responses  Wisconsin Card Sorting Test - preservative responses  Wisconsin Card Sorting Test - preservative responses  Auditory Continuous Performance Test (CPT)  Auditory Continuous Performance Test (CPT)  Auditory Continuous Performance Test (CPT)  Auditory Continuous Performance Test (CPT)  Auditory Continuous Performance Test (CPT)  Auditory Continuous Performance Test (CPT)  Auditory Continuous Performance Test (CPT)  WMS- Logical Memory immediate recall  WMS- Logical Memory immediate recall  WMS- Logical Memory immediate recall  WMS- Logical Memory immediate recall  WMS- Logical Memory immediate recall  WMS- Logical Memory immediate recall  WMS- Logical Memory immediate recall  WMS - Logical Memory delayed recall  WMS - Logical Memory delayed recall  WMS - Logical Memory delayed recall  WMS - Logical Memory delayed recall  WMS - Logical Memory delayed recall  WMS - Logical Memory delayed recall  WMS - Logical Memory delayed recall  WMS - Logical Memory % retained  WMS - Logical Memory % retained  WMS - Logical Memory % retained  WMS - Logical Memory % retained  WMS - Logical Memory % retained  WMS - Logical Memory % retained  WMS - Logical Memory % retained  WMS - Visual reproduction immediate recall  WMS - Visual reproduction immediate recall  WMS - Visual reproduction immediate recall  WMS - Visual reproduction immediate recall  WMS - Visual reproduction immediate recall  WMS - Visual reproduction immediate recall  WMS - Visual reproduction immediate recall  WMS - Visual reproduction delayed recall  WMS - Visual reproduction delayed recall  WMS - Visual reproduction delayed recall  WMS - Visual reproduction delayed recall  WMS - Visual reproduction delayed recall  WMS - Visual reproduction delayed recall  WMS - Visual reproduction delayed recall  WMS - Visual reproduction %retained  WMS - Visual reproduction %retained  WMS - Visual reproduction %retained  WMS - Visual reproduction %retained  WMS - Visual reproduction %retained  WMS - Visual reproduction %retained  WMS - Visual reproduction %retained | Volume | r= 0.65  r= 0.39 (ns)  r= 0.29 (ns)  r= 0.37 (ns)  r= 0.17 (ns)  r= -0.13 (ns)  r= 0.4 (ns)  r= -0.55*  r= -0.41* (ns)  r= -0.11* (ns)  r= -0.17* (ns)  r= -0.36* (ns)  r= 0.04* (ns)  r= -0.38* (ns)  r= -0.2 (ns)  r= -0.53  r= 0.28 (ns)  r= 0.26 (ns)  r= -0.49 (ns)  r= -0.1 (ns)  r= 0.26 (ns)  r= 0.52  r= 0.08 (ns)  r= -0.01 (ns)  r= 0.07 (ns)  r= 0.4 (ns)  r= 0.1 (ns)  r= 0.34 (ns)  r= 0.63  r= 0.18 (ns)  r= 0.02 (ns)  r= 0.04 (ns)  r= 0.37 (ns)  r= 0.06 (ns)  r= 0.33 (ns)  r= 0.42 (ns)  r= 0.11 (ns)  r= 0.1 (ns)  r= 0.15 (ns)  r= 0.28 (ns)  r= -0.13 (ns)  r= 0.22 (ns)  r= 0.50  r= 0.33 (ns)  r= 0.12 (ns)  r= 0.27 (ns)  r= 0.42 (ns)  r= -0.05 (ns)  r= 0.34 (ns)  r= 0.32 (ns)  r= 0.25 (ns)  r= -0.09 (ns)  r= 0.03 (ns)  r= 0.42 (ns)  r= -0.03 (ns)  r= 0.21 (ns)  r= 0.25 (ns)  r= 0.06 (ns)  r= -0.09 (ns)  r= 0.05 (ns)  r= 0.11 (ns)  r= 0.01 (ns)  r= 0.24 (ns) | 19 |
| Spalletta et al. (2008) | left pars opercularis white matter | WM | ‘2-back' verbal working memory task | Density | r= 0.878 | 21 |
| Stratta et al. (1997) | right caudate  left caudate  right putamen  left putamen  right accumbens  left accumbens  right caudate  left caudate  right putamen  left putamen  right accumbens  left accumbens  right caudate  left caudate  right putamen  left putamen  right accumbens  left accumbens | R&EF | WCST - categories  WCST - categories  WCST - categories  WCST - categories  WCST - categories  WCST - categories  WCST - perseverative errors  WCST - perseverative errors  WCST - perseverative errors  WCST - perseverative errors  WCST - perseverative errors  WCST - perseverative errors  WCST - unique responses  WCST - unique responses  WCST - unique responses  WCST - unique responses  WCST - unique responses  WCST - unique responses | Volume | r= 0.09 (ns)  r= 0.22 (ns)  r= 0.25 (ns)  r= 0.31 (ns)  r= 0.27 (ns)  r= 0.30 (ns)  r= -0.24 (ns)  r= -0.15 (ns)  r= 0.26 (ns)  r= 0.28 (ns)  r= 0.09 (ns)  r= 0.00 (ns)  r= -0.02 (ns)  r= -0.10 (ns)  r= -0.35 (ns)  r= -0.48  r= -0.30 (ns)  r= -0.42 | 35  28 |
| Suazo, Diez, Montes, and Molina (2014) | *Not included in meta-analyses* |  | *Incompatible with used software* |  |  |  |
| Szendi et al. (2006) | left middle frontal gyrus  total straight gyrus  third ventricle  third ventricle | WM  VisM  R&EF | Digit Span Backward  Visual Patterns Test  WCST - completed categories  WCST - conceptual level responses | Volume | r= 0.46  r= -0.43  r= -0.52  r= -0.47 | 13  12 |
| Takahashi et al. (2018) | pituitary | WM | BACS - working memory | Volume | ρ= -0.31 | 63 |
| Takahashi et al. (2020) | left ventral lateral thalamus  right ventral lateral thalamus  left ventral lateral thalamus  right ventral lateral thalamus  left ventral lateral thalamus  right ventral lateral thalamus  left ventral lateral thalamus  right ventral lateral thalamus  left ventral lateral thalamus  right ventral lateral thalamus | VM  WM  VF  ATT  R&EF | BACS verbal memory  BACS verbal memory  BACS working memory  BACS working memory  BACS verbal fluency  BACS verbal fluency  BACS attention and processing speed  BACS attention and processing speed  BACS executive functions  BACS executive functions | Volume | ρ= 0.165 (ns)  ρ= 0.293 (ns)  ρ= 0.076 (ns)  ρ= 0.273 (ns)  ρ= 0.305 (ns)  ρ= 0.253 (ns)  ρ= 0.15 (ns)  ρ= 0.316 (ns)  ρ= 0.24 (ns)  ρ= 0.26 (ns) | 60 |
| Thoma et al. (2009) | right anterior hippocampus  left anterior hippocampus  right posterior hippocampus  left posterior hippocampus  white matter  grey matter  right anterior hippocampus  left anterior hippocampus  right posterior hippocampus  left posterior hippocampus  white matter  grey matter  right anterior hippocampus  left anterior hippocampus  right posterior hippocampus  left posterior hippocampus  white matter  grey matter  right anterior hippocampus  left anterior hippocampus  right posterior hippocampus  left posterior hippocampus  white matter  grey matter  right anterior hippocampus  left anterior hippocampus  right posterior hippocampus  left posterior hippocampus  white matter  grey matter  right anterior hippocampus  left anterior hippocampus  right posterior hippocampus  left posterior hippocampus  white matter  grey matter  right anterior hippocampus  left anterior hippocampus  right posterior hippocampus  left posterior hippocampus  white matter  grey matter  right anterior hippocampus  left anterior hippocampus  right posterior hippocampus  left posterior hippocampus  white matter  grey matter  right anterior hippocampus  left anterior hippocampus  right posterior hippocampus  left posterior hippocampus  white matter  grey matter  right anterior hippocampus  left anterior hippocampus  right posterior hippocampus  left posterior hippocampus  white matter  grey matter  right anterior hippocampus  left anterior hippocampus  right posterior hippocampus  left posterior hippocampus  white matter  grey matter | ATT  WM  SP  R&EF  VM  VisM | CPT d score  CPT d score  CPT d score  CPT d score  CPT d score  CPT d score  CPT overall deficit  CPT overall deficit  CPT overall deficit  CPT overall deficit  CPT overall deficit  CPT overall deficit  Auditory consonant trigrams  Auditory consonant trigrams  Auditory consonant trigrams  Auditory consonant trigrams  Auditory consonant trigrams  Auditory consonant trigrams  Digit span forward  Digit span forward  Digit span forward  Digit span forward  Digit span forward  Digit span forward  Digit span backward  Digit span backward  Digit span backward  Digit span backward  Digit span backward  Digit span backward  Trail making test A  Trail making test A  Trail making test A  Trail making test A  Trail making test A  Trail making test A  Trail making test B  Trail making test B  Trail making test B  Trail making test B  Trail making test B  Trail making test B  WMS - Logical memory I  WMS - Logical memory I  WMS - Logical memory I  WMS - Logical memory I  WMS - Logical memory I  WMS - Logical memory I  WMS - Logical memory II  WMS - Logical memory II  WMS - Logical memory II  WMS - Logical memory II  WMS - Logical memory II  WMS - Logical memory II  WMS - Visual reproduction I  WMS - Visual reproduction I  WMS - Visual reproduction I  WMS - Visual reproduction I  WMS - Visual reproduction I  WMS - Visual reproduction I  WMS - Visual reproduction II  WMS - Visual reproduction II  WMS - Visual reproduction II  WMS - Visual reproduction II  WMS - Visual reproduction II  WMS - Visual reproduction II | Volume | r= −0.387 (ns)  r= −0.322 (ns)  r= −0.177 (ns)  r= 0.096 (ns)  r= 0.037 (ns)  r= −0.238 (ns)  r= 0.27 (ns)  r= −0.001 (ns)  r= −0.132  r= −0.424 (ns)  r= −0.336 (ns)  r= −0.231 (ns)  r= −0.477  r= 0.153 (ns)  r= 0.296 (ns)  r= 0.197 (ns)  r= −0.003 (ns)  r= −0.017 (ns)  r= −0.278 (ns)  r= 0.071 (ns)  r= −0.008 (ns)  r= 0.208 (ns)  r= 0.239 (ns)  r= 0.202 (ns)  r= −0.350  r= −0.061 (ns)  r= 0.035 (ns)  r= 0.19 (ns)  r= 0.329 (ns)  r= 0.213 (ns)  r= 0.404  r= 0.18 (ns)  r= −0.256 (ns)  r= −0.602  r= −0.134 (ns)  r= −0.466  r= 0.35  r= 0.136 (ns)  r= −0.153 (ns)  r= −0.436  r= −0.104 (ns)  r= −0.369  r= −0.471  r= −0.284 (ns)  r= 0.189 (ns)  r= 0.25 (ns)  r= 0.321 (ns)  r= 0.238 (ns)  r= −0.298 (ns)  r= −0.179 (ns)  r= 0.108 (ns)  r= 0.14 (ns)  r= 0.328 (ns)  r= 0.167 (ns)  r= −0.529  r= −0.246 (ns)  r= 0.365  r= 0.65  r= 0.278 (ns)  r= 0.233 (ns)  r= −0.450  r= −0.271 (ns)  r= 0.427  r= 0.509  r= 0.301 (ns)  r= 0.244 (ns) | 24 |
| Toulopoulou et al. (2004) | Left hippocampus  Lateral ventricles | VM  R&EF | WMS - Logical memory delayed recall  Tower of London | Volume | r= -0.22  r= 0.26 | 56 |
| Tully, Lincoln, Liyanage-Don, and Hooker (2014) | superior frontal gyrus | VF | Category fluency animal naming task | Cortical Thickness | r= 0.15 (ns) | 26 |
| van Erp et al. (2008) | Left Hippocampus - DZ  Left Hippocampus - DZ  Left Hippocampus - MZ  Left Hippocampus - DZ | VM | CVLT - free recall  CVLT - cued recall  CVLT - correct recognition  CVLT - correct recognition | Volume | r= 0.74  r= 0.57 (ns)  r= 0.65 (ns)  r= 0.71 | 55 |
| Vargas et al. (2018) | left CA1  right CA1  left CA2/3  right CA2/3  left CA4/DG  right CA4/DG  left subiculum  right subiculum  left presubiculum  right presubiculum  left CA1  right CA1  left CA2/3  right CA2/3  left CA4/DG  right CA4/DG  left subiculum  right subiculum  left presubiculum  right presubiculum  left CA1  right CA1  left CA2/3  right CA2/3  left CA4/DG  right CA4/DG  left subiculum  right subiculum  left presubiculum  right presubiculum | VisM  VM  WM | MCCB visual learning  MCCB visual learning  MCCB visual learning  MCCB visual learning  MCCB visual learning  MCCB visual learning  MCCB visual learning  MCCB visual learning  MCCB visual learning  MCCB visual learning  MCCB verbal learning  MCCB verbal learning  MCCB verbal learning  MCCB verbal learning  MCCB verbal learning  MCCB verbal learning  MCCB verbal learning  MCCB verbal learning  MCCB verbal learning  MCCB verbal learning  MCCB working memory  MCCB working memory  MCCB working memory  MCCB working memory  MCCB working memory  MCCB working memory  MCCB working memory  MCCB working memory  MCCB working memory  MCCB working memory | Volume | r= 0.06 (ns)  r= 0.14 (ns)  r= 0.04 (ns)  r= 0.25  r= 0.01 (ns)  r= 0.24  r= 0.09 (ns)  r= 0.17 (ns)  r= 0.06 (ns)  r= 0.07 (ns)  r= 0.1 (ns)  r= 0.14 (ns)  r= 0.18 (ns)  r= 0.23  r= 0.1 (ns)  r= 0.23  r= -0.02 (ns)  r= 0.16 (ns)  r= 0.1 (ns)  r= 0.1 (ns)  r= 0.16 (ns)  r= 0.24  r= 0.19  r= 0.30  r= 0.17 (ns)  r= 0.28  r= 0.17 (ns)  r= 0.30  r= 0.33  r= 0.30 | 91 |
| Vita et al. (1995) | left superior temporal gyrus  right superior temporal gyrus | VF | Verbal fluency test - semantic category  Verbal fluency test - semantic category | Volume | ρ= 0.48  ρ= 0.48 | 19 |
| Wheeler et al. (2014) | *Some results missing* |  |  |  |  |  |
| Wojtalik, Eack, and Keshavan (2013) | Left parahippocampal gyrus  Right posterior cingulate  Left parahippocampal gyrus  Right posterior cingulate  Left parahippocampal gyrus  Left parahippocampal gyrus  Left parahippocampal gyrus  Right posterior cingulate  Left parahippocampal gyrus  Right posterior cingulate | SC | MSCEI - facilitating emotions  MSCEI - facilitating emotions  MSCEI - facilitating emotions  MSCEI - facilitating emotions  MSCEI - understanding emotions  MSCEI - understanding emotions  MSCEI - managing emotions  MSCEI - managing emotions  MSCEI - managing emotions  MSCEI - managing emotions | Density  Volume  Density  Volume  Density  Volume | z= 4.22  z= 3.66  r= 0.41  r= 0.24 (ns)  z= 4.35  r= 0.34  z= 3.61  z= 3.51  r= 0.31  r= 0.34 | 51 |
| Wolfers et al. (2018) | grey matter  grey matter  white matter  white matter  grey matter  grey matter  white matter  white matter  grey matter  grey matter  white matter  white matter  grey matter  grey matter  white matter  white matter | VM  SP  WM  R&EF | Neuropsychological tests - verbal learning and memory  Neuropsychological tests - verbal learning and memory  Neuropsychological tests - verbal learning and memory  Neuropsychological tests - verbal learning and memory  Neuropsychological tests - speed of processing  Neuropsychological tests - speed of processing  Neuropsychological tests - speed of processing  Neuropsychological tests - speed of processing  Neuropsychological tests - working memory  Neuropsychological tests - working memory  Neuropsychological tests - working memory  Neuropsychological tests - working memory  Neuropsychological test - executive function  Neuropsychological test - executive function  Neuropsychological test - executive function  Neuropsychological test - executive function | %GM neg dev  %GM pos dev  %WM neg dev  %WM pos dev  %GM neg dev  %GM pos dev  %WM neg dev  %WM pos dev  %GM neg dev  %GM pos dev  %WM neg dev  %WM pos dev  %GM neg dev  %GM pos dev  %WM neg dev  %WM pos dev | p= 0.917 (ns)  p= 0.649 (ns)  p= 0.325 (ns)  p= 0.19 (ns)  p= 0.113 (ns)  p= 0.332 (ns)  p= 0.07 (ns)  p= 0.625 (ns)  p= 0.542 (ns)  p= 0.079 (ns)  p= 0.864 (ns)  p= 0.14 (ns)  p= 0.063 (ns)  p= 0.466 (ns)  p= 0.309 (ns)  p= 0.582 (ns) | 218 |
| Xie et al. (2019) | right inferior frontal gyrus  right inferior frontal gyrus | R&EF  VisM | Cognitive flexibility composite score  Visuospatial memory composite score | Cortical Thickness | p= 0.0018  p= 0.00006 | 74 |
| Yamada et al. (2007) | medial prefrontal cortex | SC | Perception of Affect Task - subtest 4 | Concentration | r= 0.464 | 20 |
| Yan et al. (2019) | left middle frontal cortex  left calcarine  right insula  left precuneus  left insula  left middle frontal cortex  left insula  left middle occipital gyrus  left rectus | SP  VF  WM  VM | Trail making test A  Trail making test A  Trail making test A  Category fluency test - animal naming  WMS-R digit span  WMS - Logical Memory  WMS - Logical Memory  WMS - Logical Memory  WMS - Logical Memory | Cortical Thickness  Sulcal depth  Thickness  Sulcal depth | r= −0.597*  r= −0.489*  r= −0.338* (ns)  r= 0.293 (ns)  r= 0.307 (ns)  r= 0.34 (ns)  r= 0.287 (ns)  r= −0.293 (ns)  r= −0.280 (ns) | 32 |
| Yang et al. (2019) | right lateral PFC | WM | Letter Number Span (LNS) - correct | Volume | r= 0.56 | 37 |
| Zierhut et al. (2013) | right superior temporal gyrus | WM | PICS - 2 back task - hits | Volume | r= 0.509 | 26 |
| Zuffante et al. (2001) | left area 46  right area 46  left area 46  right area 46  left area 46  right area 46  left area 46  right area 46 | WM | Spatial delayed response task (SDRT) - error distance  Spatial delayed response task (SDRT) - error distance  Spatial delayed response task (SDRT) - response time  Spatial delayed response task (SDRT) - response time  Abbreviated version of the SOP - verbal  Abbreviated version of the SOP - verbal  Abbreviated version of the SOP - nonverbal  Abbreviated version of the SOP - nonverbal | Volume | ρ = -0.13 (ns)  ρ = 0.2 (ns)  ρ = 0.003 (ns)  ρ = -0.11 (ns)  ρ = -0.27 (ns)  ρ = -0.3 (ns)  ρ = -0.09 (ns)  ρ = -0.2 (ns) | 23 |

*The results reported here have been reversed to the opposite sign since the scale used in this test is inverted compared to the others.

** Categories and preservative responses, Wisconsin Card-Sorting Test

*** Logical Memory Passages (immediate and 30-min delay); WMS Learning trails 1 through 5, California Verbal Learning Test

**** Design Reproduction immediate and 30-min delay; WMS

***** Benton Line Orientation Test; Block Design WAIS-R; Rosen Drawing Test

****** Logical Memory Passages (immediate and 30-min delay); WMS Learning trails 1 through 5; California Verbal Learning Test

### **Table S3.** Scoring of articles included in the meta-analysis based on five criteria to determine overall quality of studies.

| **Studies** | | | **1** | **2** | | | **3** | | | | **4** | | | | | **5** | | | **Quality** | | | | | | | | | |
| --- | --- | --- | --- | --- | --- | --- | --- | --- | --- | --- | --- | --- | --- | --- | --- | --- | --- | --- | --- | --- | --- | --- | --- | --- | --- | --- | --- | --- |
| Abbs et al. (2011) | | |  |  | | | |  | | | |  | | | |  | | | | | Low | | | | | | | |
| Antonova et al. (2005) | | |  |  | | | |  | | | |  | | | |  | | | | | Low | | | | | | | |
| Baare et al. (1999) | | |  |  | | | |  | | | |  | | | |  | | | | | Low | | | | | | | |
| Banaj et al. (2018) | | |  |  | | | |  | | | |  | | | |  | | | | | High | | | | | | | |
| Bonilha et al. (2008) | | |  |  | | | |  | | | |  | | | |  | | | | | High | | | | | | | |
| Bornstein et al. (1992) | | |  |  | | | |  | | | |  | | | |  | | | | | Low | | | | | | | |
| Caldiroli et al. (2018) | | |  |  | | | |  | | | |  | | | |  | | | | | High | | | | | | | |
| DeLisi et al. (1991) | | |  |  | | | |  | | | |  | | | |  | | | | | Low | | | | | | | |
| Dickey et al. (2007) | | |  |  | | | |  | | | |  | | | |  | | | | | Low | | | | | | | |
| Edgar et al. (2012) | | |  |  | | | |  | | | |  | | | |  | | | | | Low | | | | | | | |
| Ehrlich et al. (2012) | | |  |  | | | |  | | | |  | | | |  | | | | | Low | | | | | | | |
| Exner et al. (2004) | | |  |  | | | |  | | | |  | | | |  | | | | | Low | | | | | | | |
| Exner et al. (2008) | | |  |  | | | |  | | | |  | | | |  | | | | | Low | | | | | | | |
| Francis et al. (2016) | | |  |  | | | |  | | | |  | | | |  | | | | | High | | | | | | | |
| Frascarelli et al. (2015) | | |  |  | | | |  | | | |  | | | |  | | | | | High | | | | | | | |
| Fujiwara et al. (2007) | | |  |  | | | |  | | | |  | | | |  | | | | | Low | | | | | | | |
| Garlinghouse et al. (2010) | | |  |  | | | |  | | | |  | | | |  | | | | | Low | | | | | | | |
| (Goghari et al., 2014) | | |  |  | | | |  | | | |  | | | |  | | | | | High | | | | | | | |
| Goldberg et al. (1994) | | |  |  | | | |  | | | |  | | | |  | | | | | High | | | | | | | |
| Goldstein et al. (2011) | | |  |  | | | |  | | | |  | | | |  | | | | | Low | | | | | | | |
| Gur et al. (1999) | | |  |  | | | |  | | | |  | | | |  | | | | | High | | | | | | | |
| Gur, Cowell, et al. (2000) | | |  |  | | | |  | | | |  | | | |  | | | | | High | | | | | | | |
| Gur, Turetsky, et al. (2000) | | |  |  | | | |  | | | |  | | | |  | | | | | High | | | | | | | |
| Hartberg, Sundet, Rimol, Haukvik, Lange, Nesvag, Dale, et al. (2011) | | |  |  | | | |  | | | |  | | | |  | | | | | High | | | | | | | |
| Hartberg, Sundet, Rimol, Haukvik, Lange, Nesvag, Melle, et al. (2011) | | |  |  | | | |  | | | |  | | | |  | | | | | High | | | | | | | |
| Herold et al. (2015) | | |  |  | | | |  | | | |  | | | |  | | | | | Low | | | | | | | |
|  | | **Studies** | | | | | **1** | | | | **2** | | | | | **3** | | | | | **4** | | | | **5** | | | **Quality** |
|  | | Hirao et al. (2008) | | | | |  | | | |  | | | |  | | | | |  | | | |  | | | High | |
|  | | Hooker et al. (2011) | | | | |  | | | |  | | | |  | | | | |  | | | |  | | | High | |
|  | | Hoptman et al. (2005) | | | | |  | | | |  | | | |  | | | | |  | | | |  | | | Low | |
|  | | Hoseth et al. (2016) | | | | |  | | | |  | | | |  | | | | |  | | | |  | | | High | |
|  | | Kareken et al. (1995) | | | | |  | | | |  | | | |  | | | | |  | | | |  | | | Low | |
|  | | Karnik-Henry et al. (2012) | | | | |  | | | |  | | | |  | | | | |  | | | |  | | | Low | |
|  | | Killgore et al. (2009) | | | | |  | | | |  | | | |  | | | | |  | | | |  | | | Low | |
|  | | Knochel, Reuter, et al. (2016) | | | | |  | | | |  | | | |  | | | | |  | | | |  | | | High | |
|  | | Knochel, Stablein, et al. (2016) | | | | |  | | | |  | | | |  | | | | |  | | | |  | | | High | |
|  | | Kochunov et al. (2020) | | | | |  | | | |  | | | |  | | | | |  | | | |  | | | High | |
|  | | Koshiyama et al. (2018a) | | | | |  | | | |  | | | |  | | | | |  | | | |  | | | High | |
|  | | Koshiyama et al. (2018b) | | | | |  | | | |  | | | |  | | | | |  | | | |  | | | High | |
|  | | Krabbendam et al. (2000) | | | | |  | | | |  | | | |  | | | | |  | | | |  | | | Low | |
|  | | Kumarasinghe et al. (2014) | | | | |  | | | |  | | | |  | | | | |  | | | |  | | | Low | |
|  | | Lee et al. (2007) | | | | |  | | | |  | | | |  | | | | |  | | | |  | | | High | |
|  | | Levitt et al. (1999) | | | | |  | | | |  | | | |  | | | | |  | | | |  | | | Low | |
|  | | Levitt et al. (2013) | | | | |  | | | |  | | | |  | | | | |  | | | |  | | | Low | |
|  | | Maat et al. (2016) | | | | |  | | | |  | | | |  | | | | |  | | | |  | | | High | |
|  | | Maher et al. (1995) | | | | |  | | | |  | | | |  | | | | |  | | | |  | | | High | |
|  | | Massey et al. (2017) | | | | |  | | | |  | | | |  | | | | |  | | | |  | | | Low | |
|  | | McKenna et al. (2019) | | | | |  | | | |  | | | |  | | | | |  | | | |  | | | High | |
|  | | Molina et al. (2009) | | | | |  | | | |  | | | |  | | | | |  | | | |  | | | High | |
|  | | Namiki et al. (2007) | | | | |  | | | |  | | | |  | | | | |  | | | |  | | | High | |
|  | | Nestor et al. (1993) | | | | |  | | | |  | | | |  | | | | |  | | | |  | | | Low | |
|  | | Nestor, Onitsuka, et al. (2007) | | | | |  | | | |  | | | |  | | | | |  | | | |  | | | High | |
|  | | Nestor, Kubicki, et al. (2007) | | | | |  | | | |  | | | |  | | | | |  | | | |  | | | Low | |
|  | | Nestor et al. (2010) | | | | |  | | | |  | | | |  | | | | |  | | | |  | | | High | |
|  | | Nestor et al. (2013) | | | | |  | | | |  | | | |  | | | | |  | | | |  | | | High | |
| **Studies** | | | **1** | | **2** | | | | **3** | | | | **4** | | | | **5** | | | | | **Quality** | | | | | | |
| Nestor et al. (2020) | | |  | |  | | | |  | | | |  | | | |  | | | | | Low | | | | | | |
| Ohtani et al. (2014) | | |  | |  | | | |  | | | |  | | | |  | | | | | Low | | | | | | |
| Onitsuka et al. (2003) | | |  | |  | | | |  | | | |  | | | |  | | | | | High | | | | | | |
| Premkumar, Fannon, Kuipers, Cooke, et al. (2008) | | |  | |  | | | |  | | | |  | | | |  | | | | | Low | | | | | | |
| Premkumar, Fannon, Kuipers, Simmons, et al. (2008) | | |  | |  | | | |  | | | |  | | | |  | | | | | High | | | | | | |
| Premkumar, Kumari, et al. (2008) | | |  | |  | | | |  | | | |  | | | |  | | | | | Low | | | | | | |
| Pujol et al. (2014) | | |  | |  | | | |  | | | |  | | | |  | | | | | Low | | | | | | |
| Rametti et al. (2007) | | |  | |  | | | |  | | | |  | | | |  | | | | | High | | | | | | |
| Ridler et al. (2006) | | |  | |  | | | |  | | | |  | | | |  | | | | | Low | | | | | | |
| Rusch et al. (2008) | | |  | |  | | | |  | | | |  | | | |  | | | | | Low | | | | | | |
| Sachdev et al. (2000) | | |  | |  | | | |  | | | |  | | | |  | | | | | High | | | | | | |
| Sanfilipo et al. (2002) | | |  | |  | | | |  | | | |  | | | |  | | | | | High | | | | | | |
| Schobel et al. (2009) | | |  | |  | | | |  | | | |  | | | |  | | | | | High | | | | | | |
| Schretlen et al. (2010) | | |  | |  | | | |  | | | |  | | | |  | | | | | Low | | | | | | |
| Segarra et al. (2008) | | |  | |  | | | |  | | | |  | | | |  | | | | | High | | | | | | |
| Seidman et al. (1994) | | |  | |  | | | |  | | | |  | | | |  | | | | | High | | | | | | |
| Spalletta et al. (2008) | | |  | |  | | | |  | | | |  | | | |  | | | | | High | | | | | | |
|  | **Studies** | | | | | **1** | | | | **2** | | | | **3** | | | | **4** | | | | | **5** | | | **Quality** | | |
|  | Stratta et al. (1997) | | | | |  | | | |  | | | |  | | | |  | | | | |  | | | High | | |
|  | Szendi et al. (2006) | | | | |  | | | |  | | | |  | | | |  | | | | |  | | | Low | | |
|  | Takahashi et al. (2018) | | | | |  | | | |  | | | |  | | | |  | | | | |  | | | Low | | |
|  | Takahashi et al. (2020) | | | | |  | | | |  | | | |  | | | |  | | | | |  | | | High | | |
|  | Thoma et al. (2009) | | | | |  | | | |  | | | |  | | | |  | | | | |  | | | High | | |
|  | Toulopoulou et al. (2004) | | | | |  | | | |  | | | |  | | | |  | | | | |  | | | High | | |
|  | Tully et al. (2014) | | | | |  | | | |  | | | |  | | | |  | | | | |  | | | Low | | |
|  | van Erp et al. (2008) | | | | |  | | | |  | | | |  | | | |  | | | | |  | | | Low | | |
|  | Vargas et al. (2018) | | | | |  | | | |  | | | |  | | | |  | | | | |  | | | High | | |
|  | Vita et al. (1995) | | | | |  | | | |  | | | |  | | | |  | | | | |  | | | Low | | |
|  | Wojtalik et al. (2013) | | | | |  | | | |  | | | |  | | | |  | | | | |  | | | High | | |
|  | Wolfers et al. (2018) | | | | |  | | | |  | | | |  | | | |  | | | | |  | | | Low | | |
|  | Xie et al. (2019) | | | | |  | | | |  | | | |  | | | |  | | | | |  | | | Low | | |
|  | Yamada et al. (2007) | | | | |  | | | |  | | | |  | | | |  | | | | |  | | | Low | | |
|  | Yan et al. (2019) | | | | |  | | | |  | | | |  | | | |  | | | | |  | | | High | | |
|  | Yang et al. (2019) | | | | |  | | | |  | | | |  | | | |  | | | | |  | | | High | | |
|  | Zierhut et al. (2013) | | | | |  | | | |  | | | |  | | | |  | | | | |  | | | High | | |
|  | Zuffante et al. (2001) | | | | |  | | | |  | | | |  | | | |  | | | | |  | | | High | | |

**Criteria:** 1: Segmentation method reporting, 2: Correction for multiple comparison, 3: Covariates (age and sex), 4: Nonsignificant results reported, 5: Scanner strength and reporting. **Colors**: green is for a complete score, red for a null score and orange for half a score.

### **Figure S2.** Forest plot of the correlation between speed of processing and all brain structures reported in the included studies

**Fisher’s z [95% CI]**

**Study**

**Brain region**

-1	0	1

|  | Data |
| --- | --- |
| 0.234189 | 110 |
| 0.140926 | 109 |
| 0.13074 | 108 |
| -0.100335 | 107 |
| -0.100335 | 106 |
| -0.120581 | 105 |
| -0.100335 | 104 |
| 0.0801713 | 103 |
| -0.0801713 | 102 |
| 0.276864 | 101 |
| 0.287682 | 100 |
| 0.183016 | 99 |
| 0.141946 | 98 |
| 0.195451 | 97 |
| 0.168579 | 96 |
| 0.142966 | 95 |
| 0.137867 | 94 |
| 0.57634 | 93 |
| 0.57634 | 92 |
| -0.140926 | 91 |
| -0.213171 | 90 |
| 0.257547 | 89 |
| 0.257547 | 88 |
| 0.16755 | 87 |
| 0.174758 | 86 |
| 0.100335 | 85 |
| 0.267181 | 84 |
| 0.161387 | 83 |
| 0.249023 | 82 |
| 0.0140009 | 81 |
| -0.0140009 | 80 |
| 0.114498 | 79 |
| 0.128706 | 78 |
| -0.0480369 | 77 |
| 0.00700011 | 76 |
| 0.214218 | 75 |
| 0.224707 | 74 |
| 0.178884 | 73 |
| 0.187155 | 72 |
| 0.0510443 | 71 |
| 0.222605 | 70 |
| 0.180949 | 69 |
| 0.272554 | 68 |
| 0.136848 | 67 |
| 0.0962966 | 66 |
| 0.169608 | 65 |
| 0.16344 | 64 |
| -0.00300001 | 63 |
| 0.0550556 | 62 |
| 0.250087 | 61 |
| 0.24796 | 60 |
| 0.122611 | 59 |
| 0.131757 | 58 |
| 0.123626 | 57 |
| 0.261823 | 56 |
| 0.121596 | 55 |
| 0.190263 | 54 |
| -0.114498 | 53 |
| -0.126673 | 52 |
| 0.0460325 | 51 |
| 0.0681051 | 50 |
| -0.0882282 | 49 |
| -0.0340131 | 48 |
| 0.420083 | 47 |
| 0.710516 | 46 |
| 0.64309 | 45 |
| 0.764978 | 44 |
| 0.705742 | 43 |
| 0.549306 | 42 |
| 0.180949 | 41 |
| -0.158309 | 40 |
| 0.100335 | 39 |
| 0.384952 | 38 |
| -0.22997 | 37 |
| -0.240534 | 36 |
| -0.0530497 | 35 |
| -0.110447 | 34 |
| 0.030009 | 33 |
| 0.0701147 | 32 |
| 0.030009 | 31 |
| -0.0801713 | 30 |
| -0.0902442 | 29 |
| -0.0801713 | 28 |
| -0.0801713 | 27 |
| -0.140926 | 26 |
| -0.13074 | 25 |
| -0.161387 | 24 |
| 0.181983 | 23 |
| 0.202733 | 22 |
| 0.161387 | 21 |
| 0.110447 | 20 |
| 0.140926 | 19 |
| 0.0400214 | 18 |
| 0.287682 | 17 |
| 0.0400214 | 16 |
| 0.255413 | 15 |
| -0.0600722 | 14 |
| 0.270403 | 13 |
| 0.120581 | 12 |
| 0.403571 | 11 |
| 0.132775 | 10 |
| 0.242653 | 9 |
| 0.345074 | 8 |
| -0.108065 | 7 |
| 0.066107 | 6 |
| -0.123584 | 5 |
| -0.0332986 | 4 |
| 0.534745 | 3 |
| 0.351833 | 2 |
| 0.688473 | 1 |
| 0.115375 | -2 |

|  | studies |
| --- | --- |
| -3.5 | 110 |
| -3.5 | 109 |
| -3.5 | 108 |
| -3.5 | 107 |
| -3.5 | 106 |
| -3.5 | 105 |
| -3.5 | 104 |
| -3.5 | 103 |
| -3.5 | 102 |
| -3.5 | 101 |
| -3.5 | 100 |
| -3.5 | 99 |
| -3.5 | 98 |
| -3.5 | 97 |
| -3.5 | 96 |
| -3.5 | 95 |
| -3.5 | 94 |
| -3.5 | 93 |
| -3.5 | 92 |
| -3.5 | 91 |
| -3.5 | 90 |
| -3.5 | 89 |
| -3.5 | 88 |
| -3.5 | 87 |
| -3.5 | 86 |
| -3.5 | 85 |
| -3.5 | 84 |
| -3.5 | 83 |
| -3.5 | 82 |
| -3.5 | 81 |
| -3.5 | 80 |
| -3.5 | 79 |
| -3.5 | 78 |
| -3.5 | 77 |
| -3.5 | 76 |
| -3.5 | 75 |
| -3.5 | 74 |
| -3.5 | 73 |
| -3.5 | 72 |
| -3.5 | 71 |
| -3.5 | 70 |
| -3.5 | 69 |
| -3.5 | 68 |
| -3.5 | 67 |
| -3.5 | 66 |
| -3.5 | 65 |
| -3.5 | 64 |
| -3.5 | 63 |
| -3.5 | 62 |
| -3.5 | 61 |
| -3.5 | 60 |
| -3.5 | 59 |
| -3.5 | 58 |
| -3.5 | 57 |
| -3.5 | 56 |
| -3.5 | 55 |
| -3.5 | 54 |
| -3.5 | 53 |
| -3.5 | 52 |
| -3.5 | 51 |
| -3.5 | 50 |
| -3.5 | 49 |
| -3.5 | 48 |
| -3.5 | 47 |
| -3.5 | 46 |
| -3.5 | 45 |
| -3.5 | 44 |
| -3.5 | 43 |
| -3.5 | 42 |
| -3.5 | 41 |
| -3.5 | 40 |
| -3.5 | 39 |
| -3.5 | 38 |
| -3.5 | 37 |
| -3.5 | 36 |
| -3.5 | 35 |
| -3.5 | 34 |
| -3.5 | 33 |
| -3.5 | 32 |
| -3.5 | 31 |
| -3.5 | 30 |
| -3.5 | 29 |
| -3.5 | 28 |
| -3.5 | 27 |
| -3.5 | 26 |
| -3.5 | 25 |
| -3.5 | 24 |
| -3.5 | 23 |
| -3.5 | 22 |
| -3.5 | 21 |
| -3.5 | 20 |
| -3.5 | 19 |
| -3.5 | 18 |
| -3.5 | 17 |
| -3.5 | 16 |
| -3.5 | 15 |
| -3.5 | 14 |
| -3.5 | 13 |
| -3.5 | 12 |
| -3.5 | 11 |
| -3.5 | 10 |
| -3.5 | 9 |
| -3.5 | 8 |
| -3.5 | 7 |
| -3.5 | 6 |
| -3.5 | 5 |
| -3.5 | 4 |
| -3.5 | 3 |
| -3.5 | 2 |
| -3.5 | 1 |
| -3.5 | -2 |

|  | regions |
| --- | --- |
| -2 | 110 |
| -2 | 109 |
| -2 | 108 |
| -2 | 107 |
| -2 | 106 |
| -2 | 105 |
| -2 | 104 |
| -2 | 103 |
| -2 | 102 |
| -2 | 101 |
| -2 | 100 |
| -2 | 99 |
| -2 | 98 |
| -2 | 97 |
| -2 | 96 |
| -2 | 95 |
| -2 | 94 |
| -2 | 93 |
| -2 | 92 |
| -2 | 91 |
| -2 | 90 |
| -2 | 89 |
| -2 | 88 |
| -2 | 87 |
| -2 | 86 |
| -2 | 85 |
| -2 | 84 |
| -2 | 83 |
| -2 | 82 |
| -2 | 81 |
| -2 | 80 |
| -2 | 79 |
| -2 | 78 |
| -2 | 77 |
| -2 | 76 |
| -2 | 75 |
| -2 | 74 |
| -2 | 73 |
| -2 | 72 |
| -2 | 71 |
| -2 | 70 |
| -2 | 69 |
| -2 | 68 |
| -2 | 67 |
| -2 | 66 |
| -2 | 65 |
| -2 | 64 |
| -2 | 63 |
| -2 | 62 |
| -2 | 61 |
| -2 | 60 |
| -2 | 59 |
| -2 | 58 |
| -2 | 57 |
| -2 | 56 |
| -2 | 55 |
| -2 | 54 |
| -2 | 53 |
| -2 | 52 |
| -2 | 51 |
| -2 | 50 |
| -2 | 49 |
| -2 | 48 |
| -2 | 47 |
| -2 | 46 |
| -2 | 45 |
| -2 | 44 |
| -2 | 43 |
| -2 | 42 |
| -2 | 41 |
| -2 | 40 |
| -2 | 39 |
| -2 | 38 |
| -2 | 37 |
| -2 | 36 |
| -2 | 35 |
| -2 | 34 |
| -2 | 33 |
| -2 | 32 |
| -2 | 31 |
| -2 | 30 |
| -2 | 29 |
| -2 | 28 |
| -2 | 27 |
| -2 | 26 |
| -2 | 25 |
| -2 | 24 |
| -2 | 23 |
| -2 | 22 |
| -2 | 21 |
| -2 | 20 |
| -2 | 19 |
| -2 | 18 |
| -2 | 17 |
| -2 | 16 |
| -2 | 15 |
| -2 | 14 |
| -2 | 13 |
| -2 | 12 |
| -2 | 11 |
| -2 | 10 |
| -2 | 9 |
| -2 | 8 |
| -2 | 7 |
| -2 | 6 |
| -2 | 5 |
| -2 | 4 |
| -2 | 3 |
| -2 | 2 |
| -2 | 1 |
| -2 | -2 |

|  | values |
| --- | --- |
| 2 | 110 |
| 2 | 109 |
| 2 | 108 |
| 2 | 107 |
| 2 | 106 |
| 2 | 105 |
| 2 | 104 |
| 2 | 103 |
| 2 | 102 |
| 2 | 101 |
| 2 | 100 |
| 2 | 99 |
| 2 | 98 |
| 2 | 97 |
| 2 | 96 |
| 2 | 95 |
| 2 | 94 |
| 2 | 93 |
| 2 | 92 |
| 2 | 91 |
| 2 | 90 |
| 2 | 89 |
| 2 | 88 |
| 2 | 87 |
| 2 | 86 |
| 2 | 85 |
| 2 | 84 |
| 2 | 83 |
| 2 | 82 |
| 2 | 81 |
| 2 | 80 |
| 2 | 79 |
| 2 | 78 |
| 2 | 77 |
| 2 | 76 |
| 2 | 75 |
| 2 | 74 |
| 2 | 73 |
| 2 | 72 |
| 2 | 71 |
| 2 | 70 |
| 2 | 69 |
| 2 | 68 |
| 2 | 67 |
| 2 | 66 |
| 2 | 65 |
| 2 | 64 |
| 2 | 63 |
| 2 | 62 |
| 2 | 61 |
| 2 | 60 |
| 2 | 59 |
| 2 | 58 |
| 2 | 57 |
| 2 | 56 |
| 2 | 55 |
| 2 | 54 |
| 2 | 53 |
| 2 | 52 |
| 2 | 51 |
| 2 | 50 |
| 2 | 49 |
| 2 | 48 |
| 2 | 47 |
| 2 | 46 |
| 2 | 45 |
| 2 | 44 |
| 2 | 43 |
| 2 | 42 |
| 2 | 41 |
| 2 | 40 |
| 2 | 39 |
| 2 | 38 |
| 2 | 37 |
| 2 | 36 |
| 2 | 35 |
| 2 | 34 |
| 2 | 33 |
| 2 | 32 |
| 2 | 31 |
| 2 | 30 |
| 2 | 29 |
| 2 | 28 |
| 2 | 27 |
| 2 | 26 |
| 2 | 25 |
| 2 | 24 |
| 2 | 23 |
| 2 | 22 |
| 2 | 21 |
| 2 | 20 |
| 2 | 19 |
| 2 | 18 |
| 2 | 17 |
| 2 | 16 |
| 2 | 15 |
| 2 | 14 |
| 2 | 13 |
| 2 | 12 |
| 2 | 11 |
| 2 | 10 |
| 2 | 9 |
| 2 | 8 |
| 2 | 7 |
| 2 | 6 |
| 2 | 5 |
| 2 | 4 |
| 2 | 3 |
| 2 | 2 |
| 2 | 1 |
| 2 | -2 |

### **Figure S3.** Forest plot of the correlation between attention and vigilance and all brain structures reported in included studies

-1	0	1

**Study**

**Brain region**

**Fisher’s z [95% CI]**

|  | Data |
| --- | --- |
| 0.52689 | 84 |
| 0.468517 | 83 |
| 0.497311 | 82 |
| 0.604156 | 81 |
| 0.181983 | 80 |
| 0.522984 | 79 |
| 0.320545 | 78 |
| 0.255413 | 77 |
| 0.266108 | 76 |
| 0.0500417 | 75 |
| 0.30952 | 74 |
| 0.342828 | 73 |
| 0.57634 | 72 |
| 0.181983 | 71 |
| 0.354093 | 70 |
| 0.376886 | 69 |
| 0.00900024 | 68 |
| 0.181983 | 67 |
| -0.161387 | 66 |
| -0.15114 | 65 |
| -0.171667 | 64 |
| -0.287682 | 63 |
| 0.354093 | 62 |
| -0.13074 | 61 |
| -0.618381 | 60 |
| -0.522984 | 59 |
| 0.120581 | 58 |
| 0.123626 | 57 |
| 0.0761468 | 56 |
| -0.0130007 | 55 |
| -0.0340131 | 54 |
| 0.257547 | 53 |
| 0.233134 | 52 |
| 0.0882282 | 51 |
| 0.235246 | 50 |
| -0.033012 | 49 |
| 0.0801713 | 48 |
| 0.0821845 | 47 |
| 0.0350143 | 46 |
| 0.0932695 | 45 |
| 0.164467 | 44 |
| -0.0902442 | 43 |
| -0.15114 | 42 |
| -0.0200027 | 41 |
| 0.287682 | 40 |
| 0.0500417 | 39 |
| 0.030009 | 38 |
| -0.181983 | 37 |
| -0.171667 | 36 |
| 0.276864 | 35 |
| 0.171667 | 34 |
| 0.298566 | 33 |
| 0.298566 | 32 |
| 0.320545 | 31 |
| 0.0400214 | 30 |
| 0.100335 | 29 |
| -0.030009 | 28 |
| 0.202733 | 27 |
| 0.4118 | 26 |
| 0.100335 | 25 |
| 0.234189 | 24 |
| 0.105388 | 23 |
| -0.233134 | 22 |
| 0.403571 | 21 |
| 0.29094 | 20 |
| -0.202733 | 19 |
| -0.590145 | 18 |
| -0.53606 | 17 |
| 0.287682 | 16 |
| 0.266108 | 15 |
| -0.100335 | 14 |
| 0.266108 | 13 |
| -0.242653 | 12 |
| -0.235246 | 11 |
| -0.333877 | 10 |
| -0.408267 | 9 |
| -0.178884 | 8 |
| 0.0962966 | 7 |
| -0.001 | 6 |
| 0.276864 | 5 |
| -0.132775 | 4 |
| -0.452559 | 3 |
| 0.0370169 | 2 |
| -0.349577 | 1 |
| 0.0962056 | -2 |

|  | Labels |
| --- | --- |
| -3 | 84 |
| -3 | 83 |
| -3 | 82 |
| -3 | 81 |
| -3 | 80 |
| -3 | 79 |
| -3 | 78 |
| -3 | 77 |
| -3 | 76 |
| -3 | 75 |
| -3 | 74 |
| -3 | 73 |
| -3 | 72 |
| -3 | 71 |
| -3 | 70 |
| -3 | 69 |
| -3 | 68 |
| -3 | 67 |
| -3 | 66 |
| -3 | 65 |
| -3 | 64 |
| -3 | 63 |
| -3 | 62 |
| -3 | 61 |
| -3 | 60 |
| -3 | 59 |
| -3 | 58 |
| -3 | 57 |
| -3 | 56 |
| -3 | 55 |
| -3 | 54 |
| -3 | 53 |
| -3 | 52 |
| -3 | 51 |
| -3 | 50 |
| -3 | 49 |
| -3 | 48 |
| -3 | 47 |
| -3 | 46 |
| -3 | 45 |
| -3 | 44 |
| -3 | 43 |
| -3 | 42 |
| -3 | 41 |
| -3 | 40 |
| -3 | 39 |
| -3 | 38 |
| -3 | 37 |
| -3 | 36 |
| -3 | 35 |
| -3 | 34 |
| -3 | 33 |
| -3 | 32 |
| -3 | 31 |
| -3 | 30 |
| -3 | 29 |
| -3 | 28 |
| -3 | 27 |
| -3 | 26 |
| -3 | 25 |
| -3 | 24 |
| -3 | 23 |
| -3 | 22 |
| -3 | 21 |
| -3 | 20 |
| -3 | 19 |
| -3 | 18 |
| -3 | 17 |
| -3 | 16 |
| -3 | 15 |
| -3 | 14 |
| -3 | 13 |
| -3 | 12 |
| -3 | 11 |
| -3 | 10 |
| -3 | 9 |
| -3 | 8 |
| -3 | 7 |
| -3 | 6 |
| -3 | 5 |
| -3 | 4 |
| -3 | 3 |
| -3 | 2 |
| -3 | 1 |
| -3 | -2 |

|  | Region |
| --- | --- |
| -2 | 84 |
| -2 | 83 |
| -2 | 82 |
| -2 | 81 |
| -2 | 80 |
| -2 | 79 |
| -2 | 78 |
| -2 | 77 |
| -2 | 76 |
| -2 | 75 |
| -2 | 74 |
| -2 | 73 |
| -2 | 72 |
| -2 | 71 |
| -2 | 70 |
| -2 | 69 |
| -2 | 68 |
| -2 | 67 |
| -2 | 66 |
| -2 | 65 |
| -2 | 64 |
| -2 | 63 |
| -2 | 62 |
| -2 | 61 |
| -2 | 60 |
| -2 | 59 |
| -2 | 58 |
| -2 | 57 |
| -2 | 56 |
| -2 | 55 |
| -2 | 54 |
| -2 | 53 |
| -2 | 52 |
| -2 | 51 |
| -2 | 50 |
| -2 | 49 |
| -2 | 48 |
| -2 | 47 |
| -2 | 46 |
| -2 | 45 |
| -2 | 44 |
| -2 | 43 |
| -2 | 42 |
| -2 | 41 |
| -2 | 40 |
| -2 | 39 |
| -2 | 38 |
| -2 | 37 |
| -2 | 36 |
| -2 | 35 |
| -2 | 34 |
| -2 | 33 |
| -2 | 32 |
| -2 | 31 |
| -2 | 30 |
| -2 | 29 |
| -2 | 28 |
| -2 | 27 |
| -2 | 26 |
| -2 | 25 |
| -2 | 24 |
| -2 | 23 |
| -2 | 22 |
| -2 | 21 |
| -2 | 20 |
| -2 | 19 |
| -2 | 18 |
| -2 | 17 |
| -2 | 16 |
| -2 | 15 |
| -2 | 14 |
| -2 | 13 |
| -2 | 12 |
| -2 | 11 |
| -2 | 10 |
| -2 | 9 |
| -2 | 8 |
| -2 | 7 |
| -2 | 6 |
| -2 | 5 |
| -2 | 4 |
| -2 | 3 |
| -2 | 2 |
| -2 | 1 |
| -2 | -2 |

|  | values |
| --- | --- |
| 1.5 | 84 |
| 1.5 | 83 |
| 1.5 | 82 |
| 1.5 | 81 |
| 1.5 | 80 |
| 1.5 | 79 |
| 1.5 | 78 |
| 1.5 | 77 |
| 1.5 | 76 |
| 1.5 | 75 |
| 1.5 | 74 |
| 1.5 | 73 |
| 1.5 | 72 |
| 1.5 | 71 |
| 1.5 | 70 |
| 1.5 | 69 |
| 1.5 | 68 |
| 1.5 | 67 |
| 1.5 | 66 |
| 1.5 | 65 |
| 1.5 | 64 |
| 1.5 | 63 |
| 1.5 | 62 |
| 1.5 | 61 |
| 1.5 | 60 |
| 1.5 | 59 |
| 1.5 | 58 |
| 1.5 | 57 |
| 1.5 | 56 |
| 1.5 | 55 |
| 1.5 | 54 |
| 1.5 | 53 |
| 1.5 | 52 |
| 1.5 | 51 |
| 1.5 | 50 |
| 1.5 | 49 |
| 1.5 | 48 |
| 1.5 | 47 |
| 1.5 | 46 |
| 1.5 | 45 |
| 1.5 | 44 |
| 1.5 | 43 |
| 1.5 | 42 |
| 1.5 | 41 |
| 1.5 | 40 |
| 1.5 | 39 |
| 1.5 | 38 |
| 1.5 | 37 |
| 1.5 | 36 |
| 1.5 | 35 |
| 1.5 | 34 |
| 1.5 | 33 |
| 1.5 | 32 |
| 1.5 | 31 |
| 1.5 | 30 |
| 1.5 | 29 |
| 1.5 | 28 |
| 1.5 | 27 |
| 1.5 | 26 |
| 1.5 | 25 |
| 1.5 | 24 |
| 1.5 | 23 |
| 1.5 | 22 |
| 1.5 | 21 |
| 1.5 | 20 |
| 1.5 | 19 |
| 1.5 | 18 |
| 1.5 | 17 |
| 1.5 | 16 |
| 1.5 | 15 |
| 1.5 | 14 |
| 1.5 | 13 |
| 1.5 | 12 |
| 1.5 | 11 |
| 1.5 | 10 |
| 1.5 | 9 |
| 1.5 | 8 |
| 1.5 | 7 |
| 1.5 | 6 |
| 1.5 | 5 |
| 1.5 | 4 |
| 1.5 | 3 |
| 1.5 | 2 |
| 1.5 | 1 |
| 1.5 | -2 |

### **Figure S4.** Forest plot of the correlation between working memory and all brain structures reported in the included studies

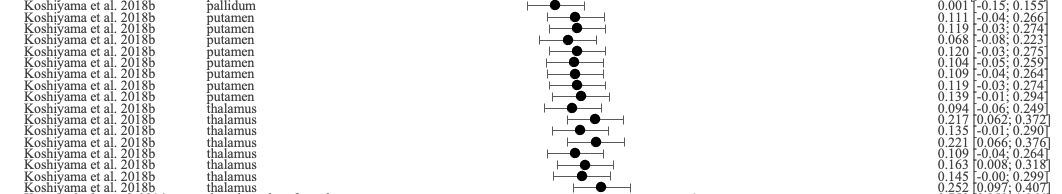

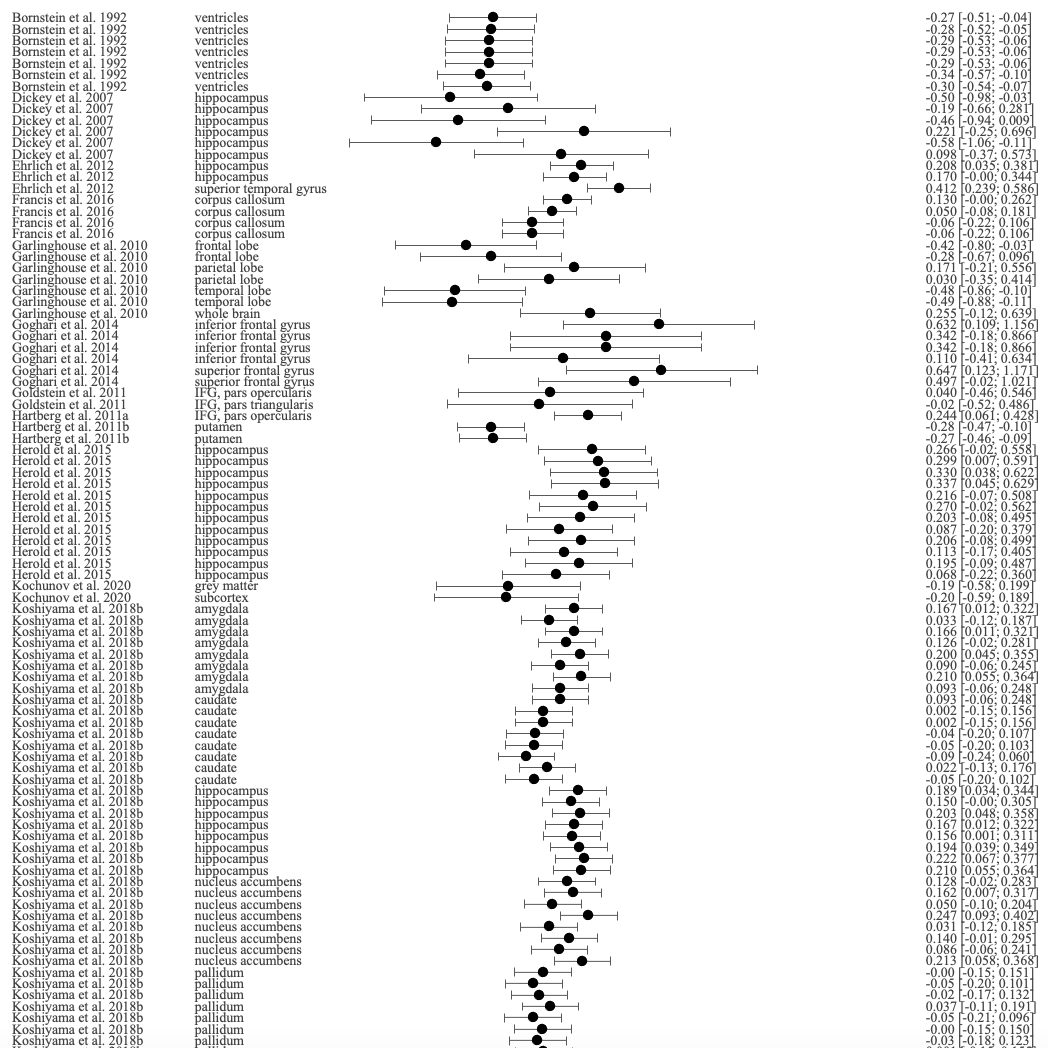

**Fisher’s z [95% CI]**

**Brain region**

**Study**

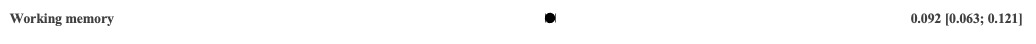

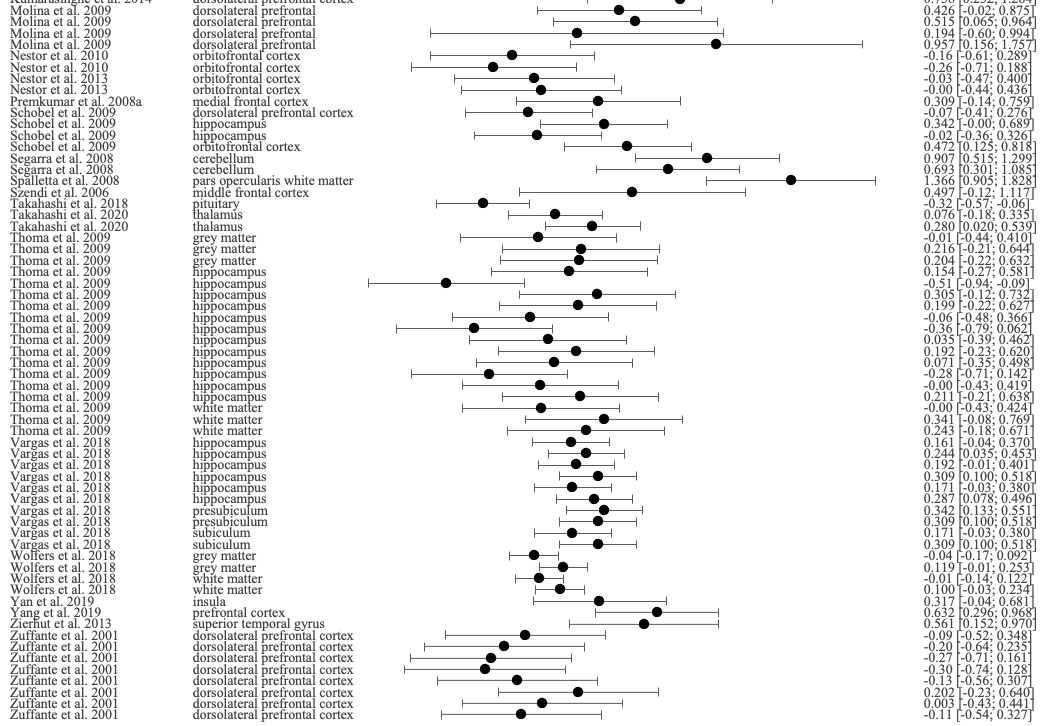

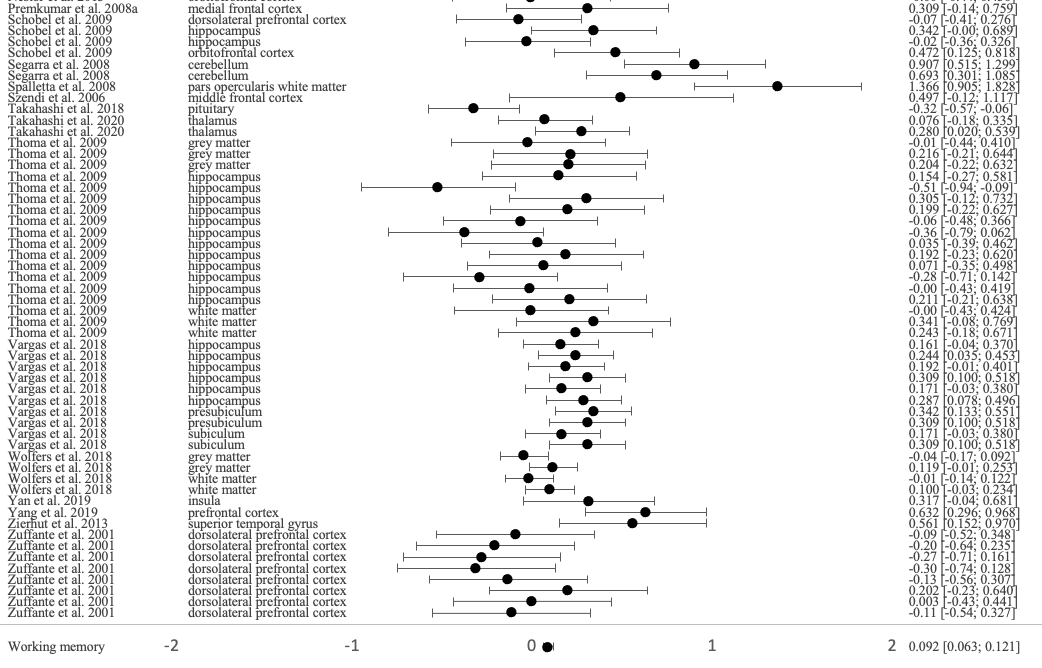

-1	0	1

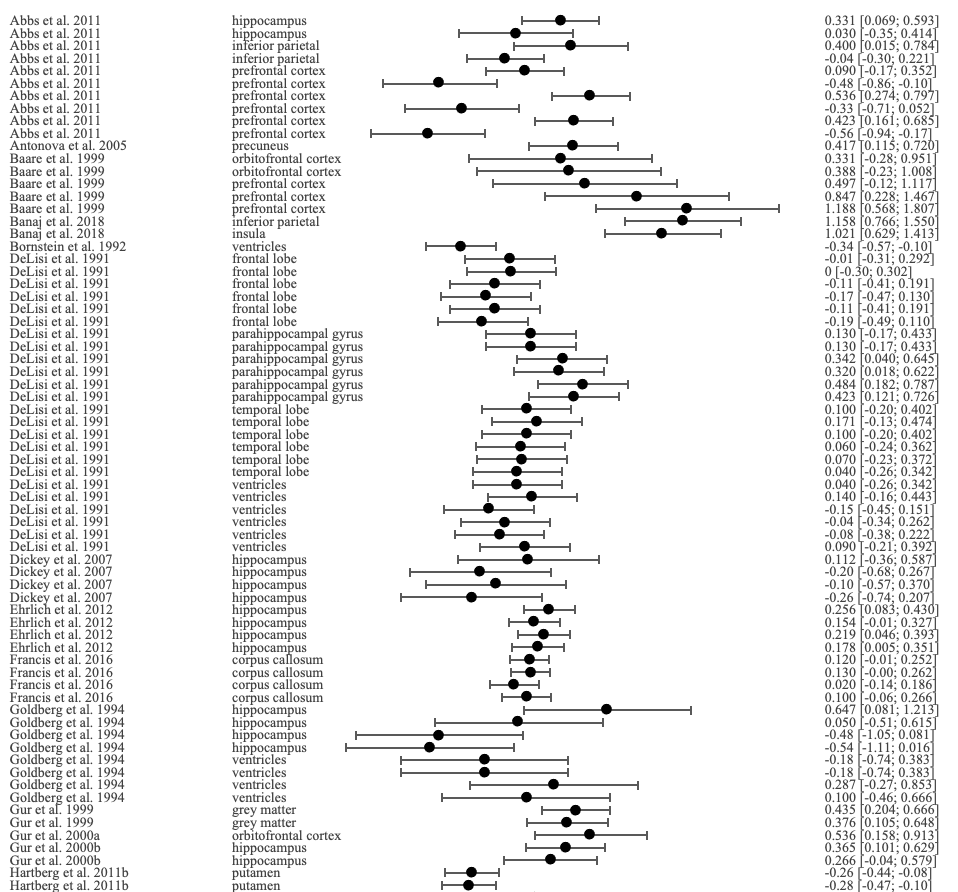

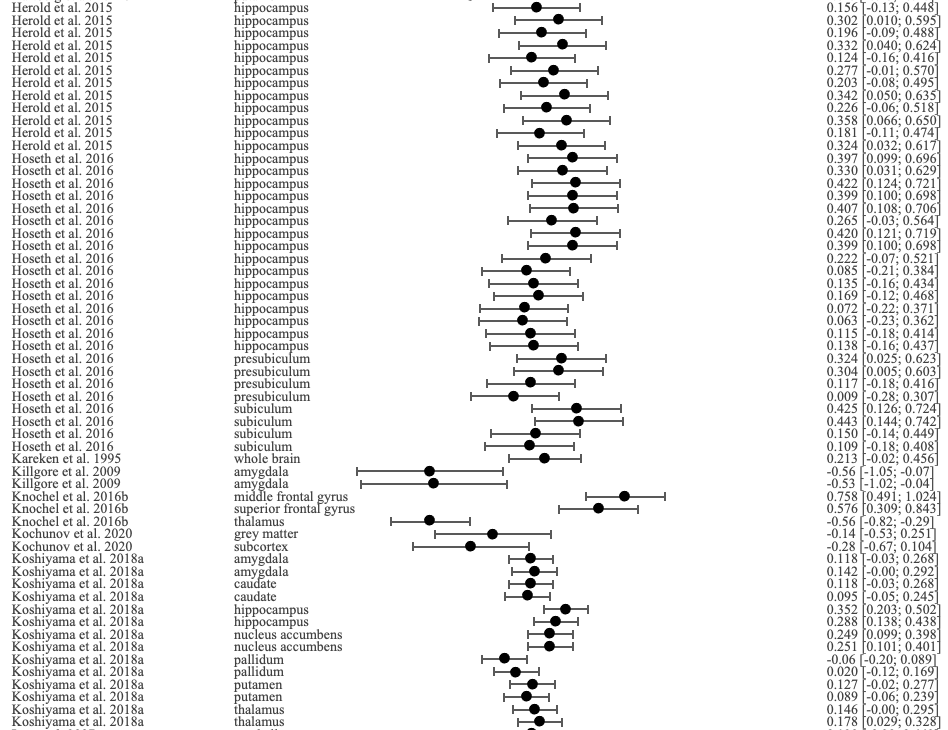

### **Figure S5.** Forest plot of the correlation between verbal learning and memory and all brain structures reported in the included studies

**Study**

**Brain region**

**Fisher’s z [95% CI]**

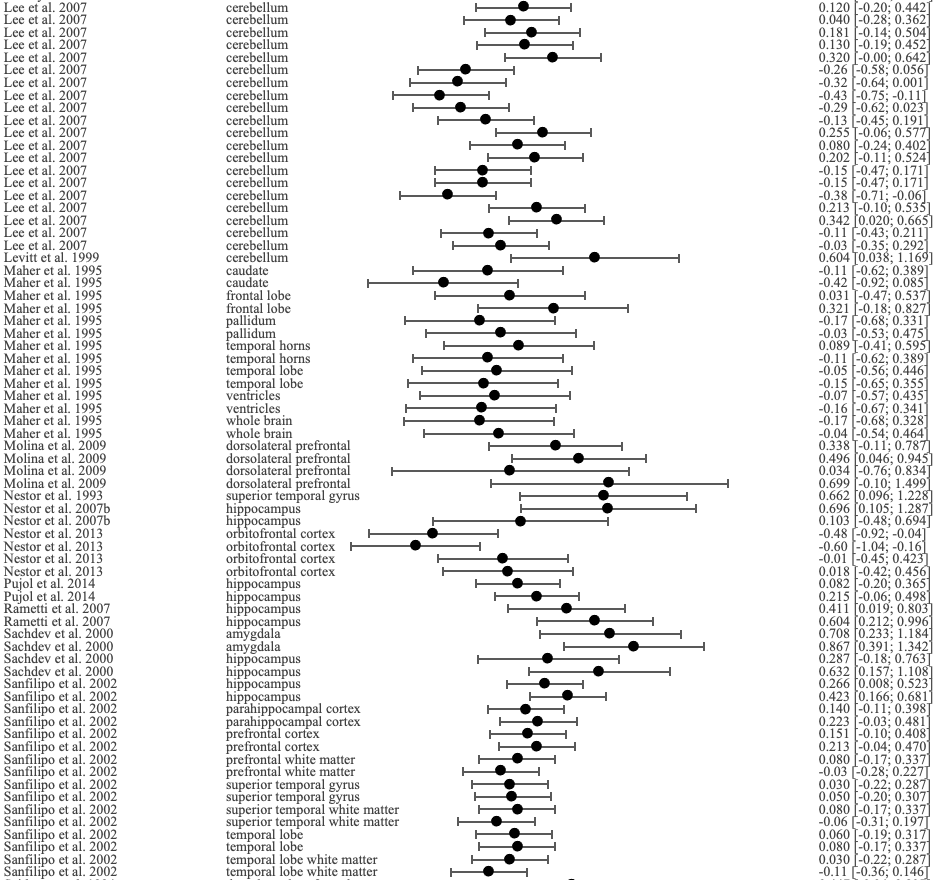

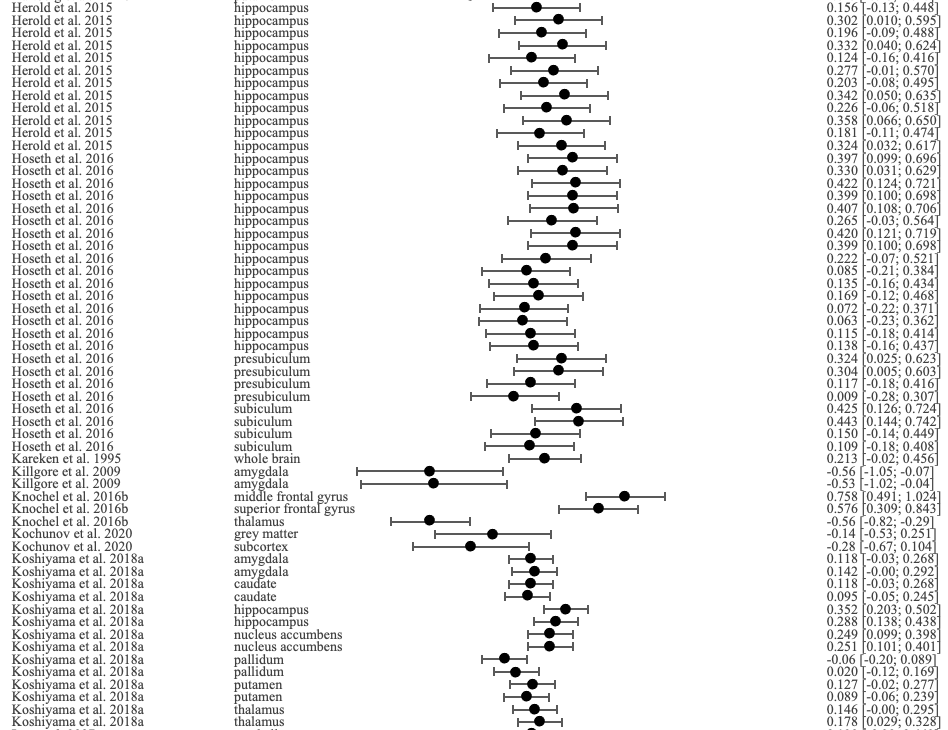

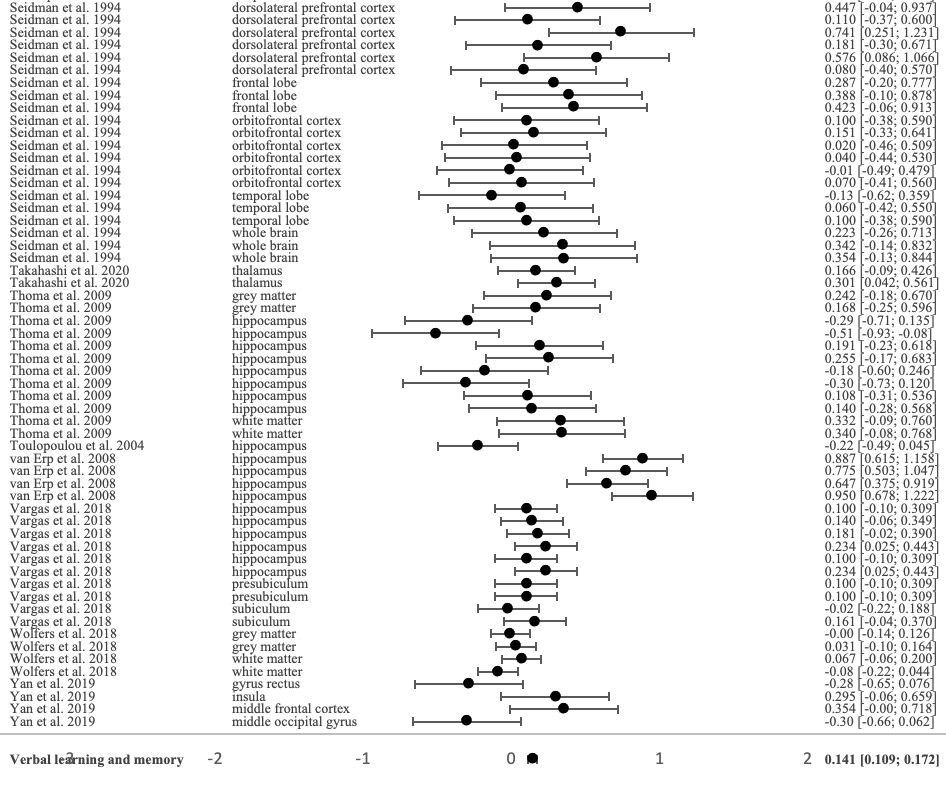

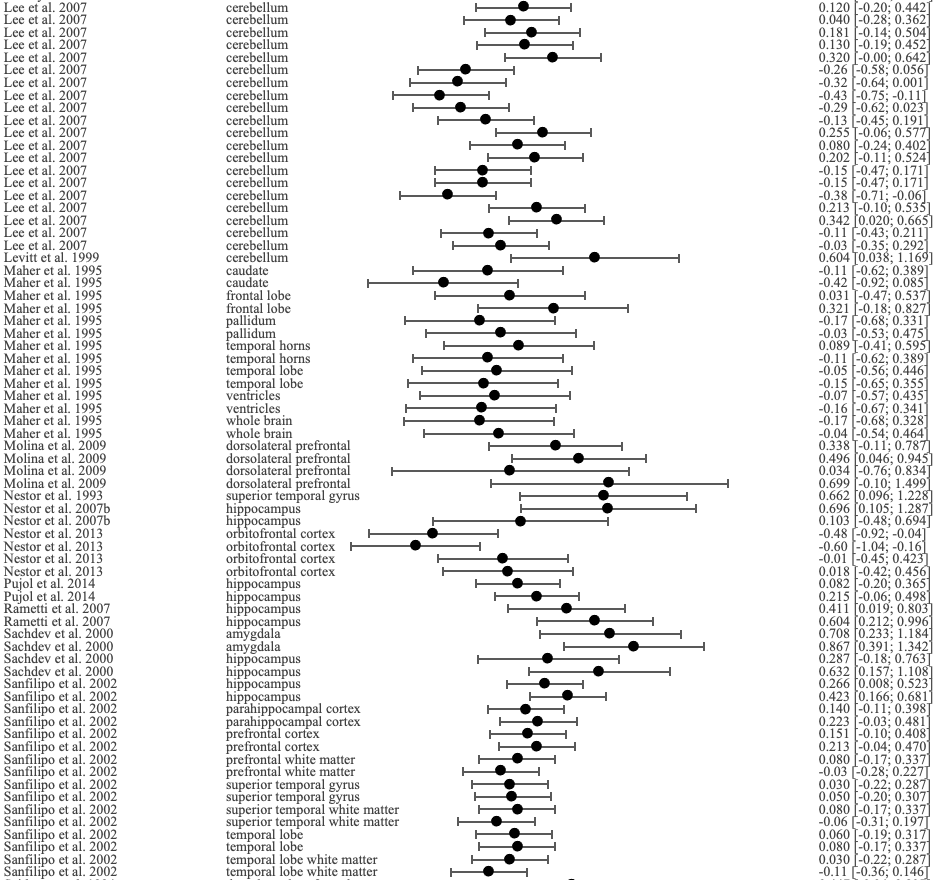

-1	0	1

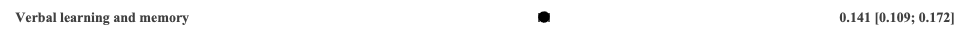

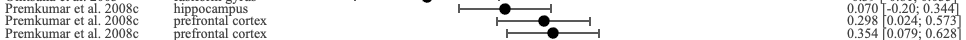

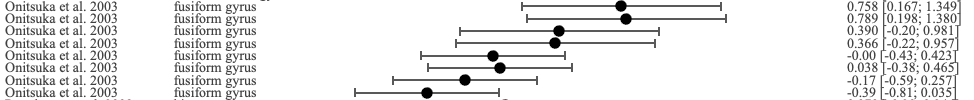

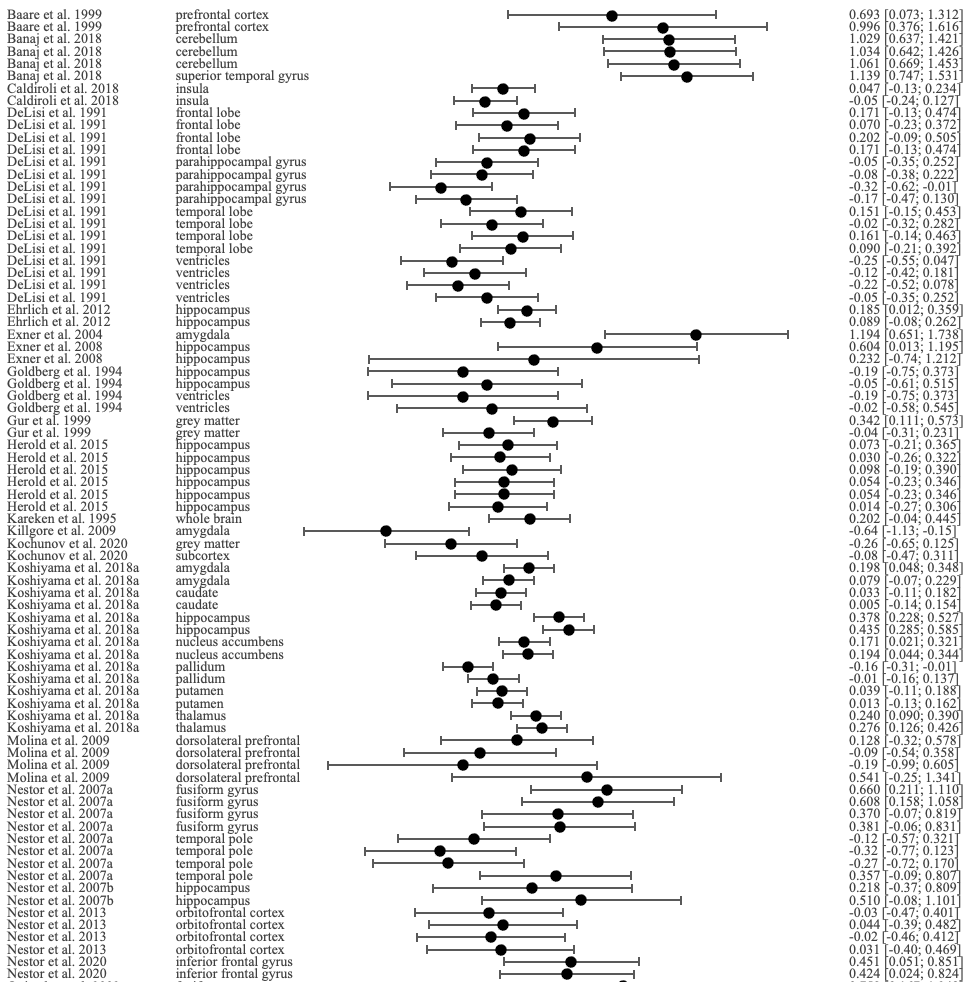

### **Figure S6.** Forest plot of the correlation between visual learning and memory and all brain structures reported in the included studies

**Fisher’s z [95% CI]**

**Brain region**

**Study**

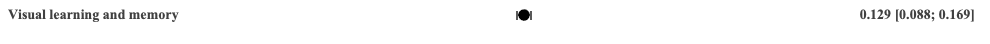

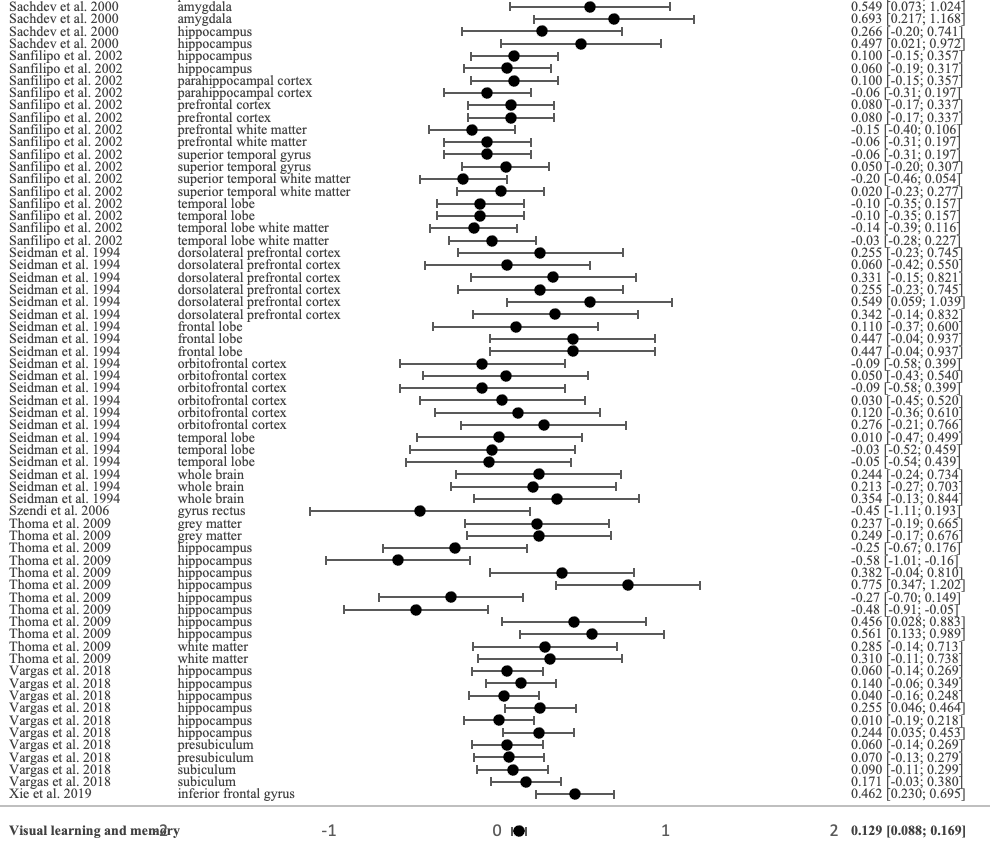

-1	0	1

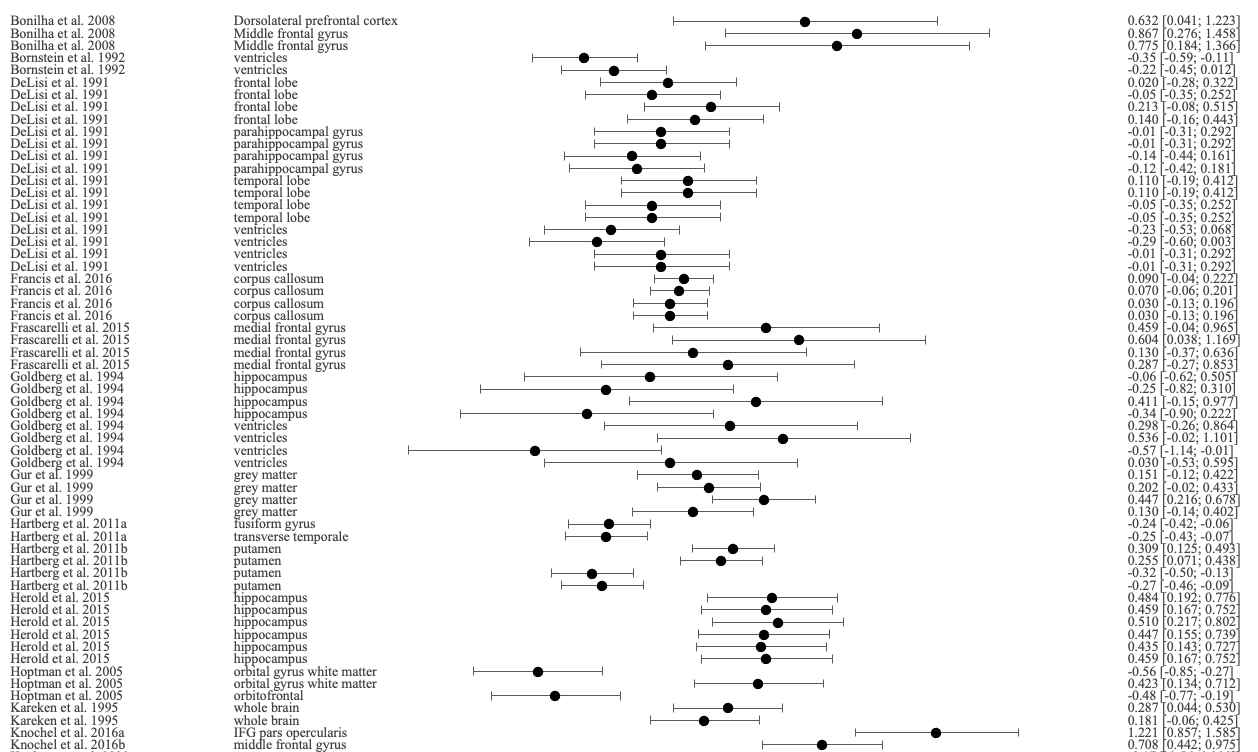

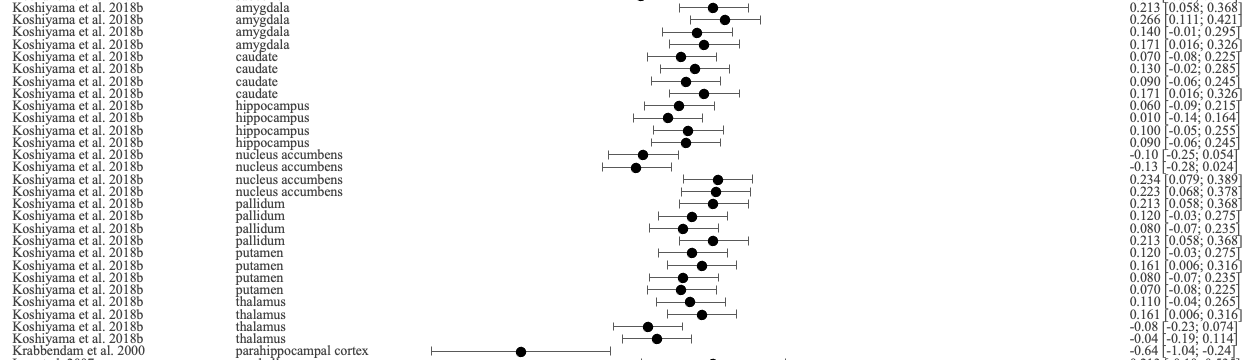

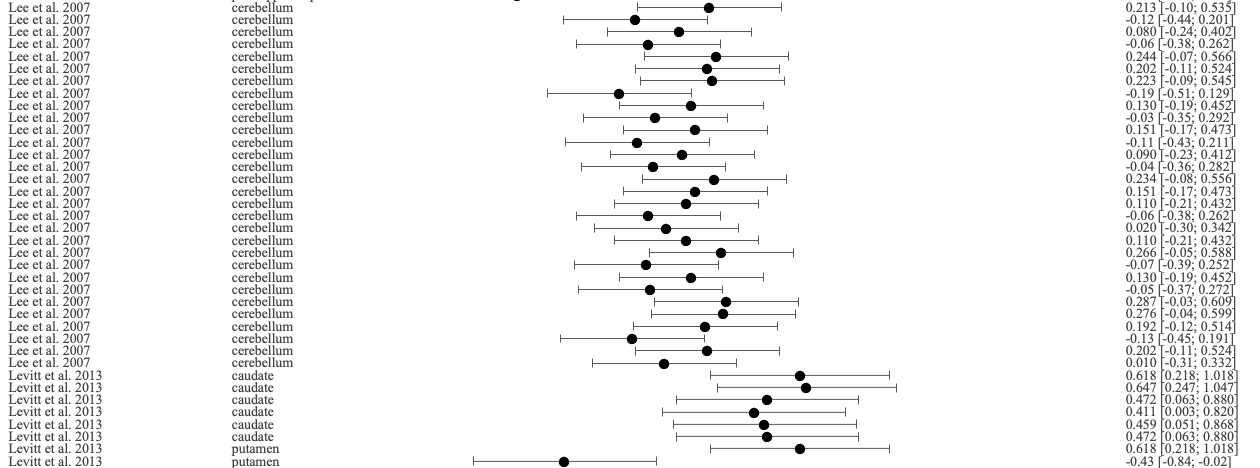

### **Figure S7.** Forest plot of the correlation between reasoning and executive functions and all brain structures reported in the included studies

**Study**

**Brain region**

**Fisher’s z [95% CI]**

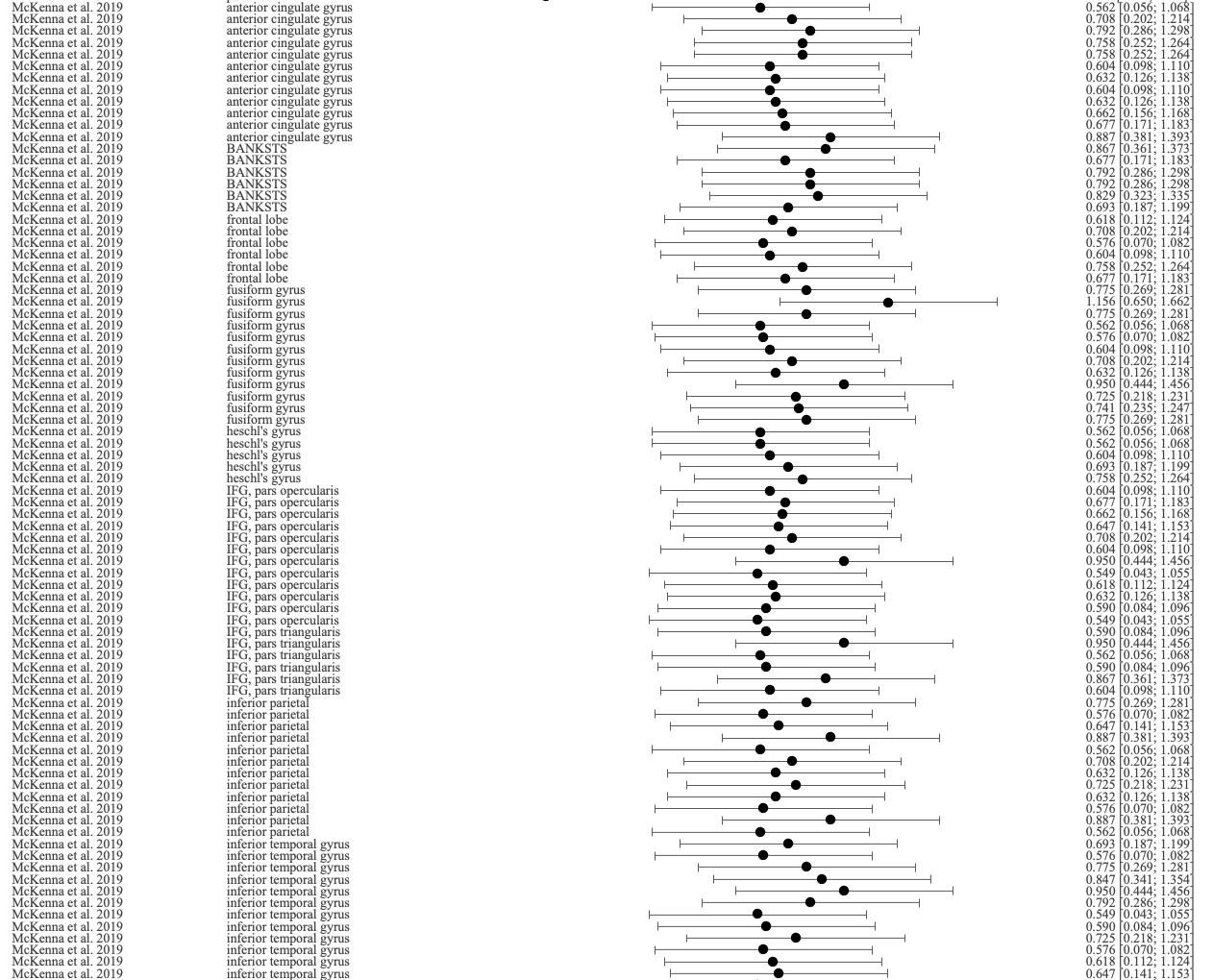

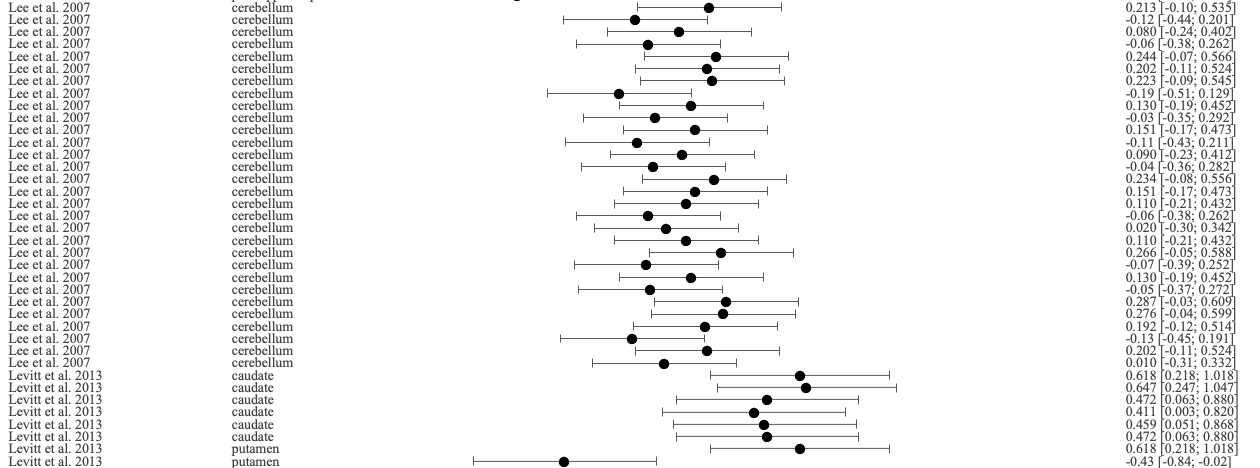

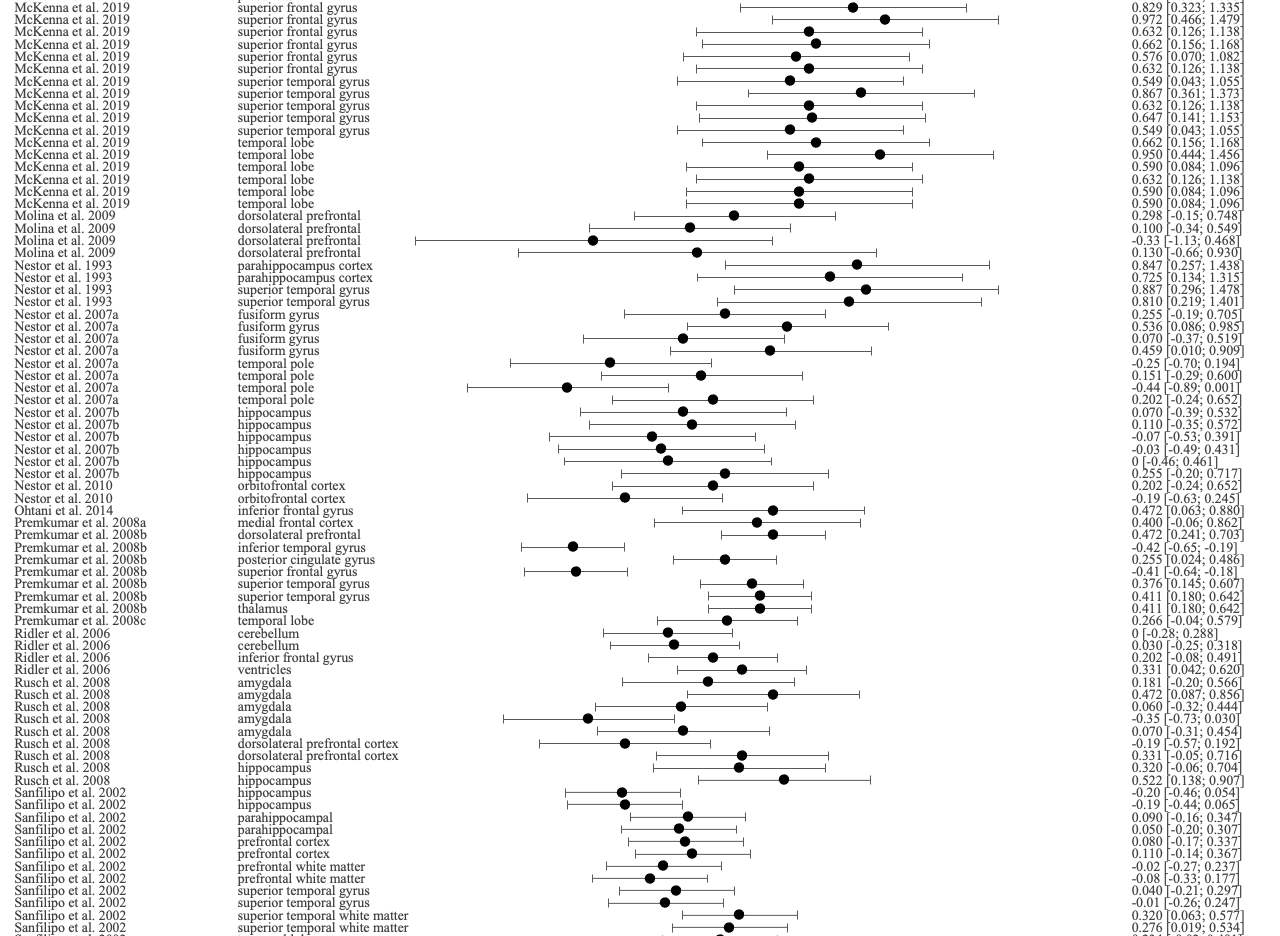

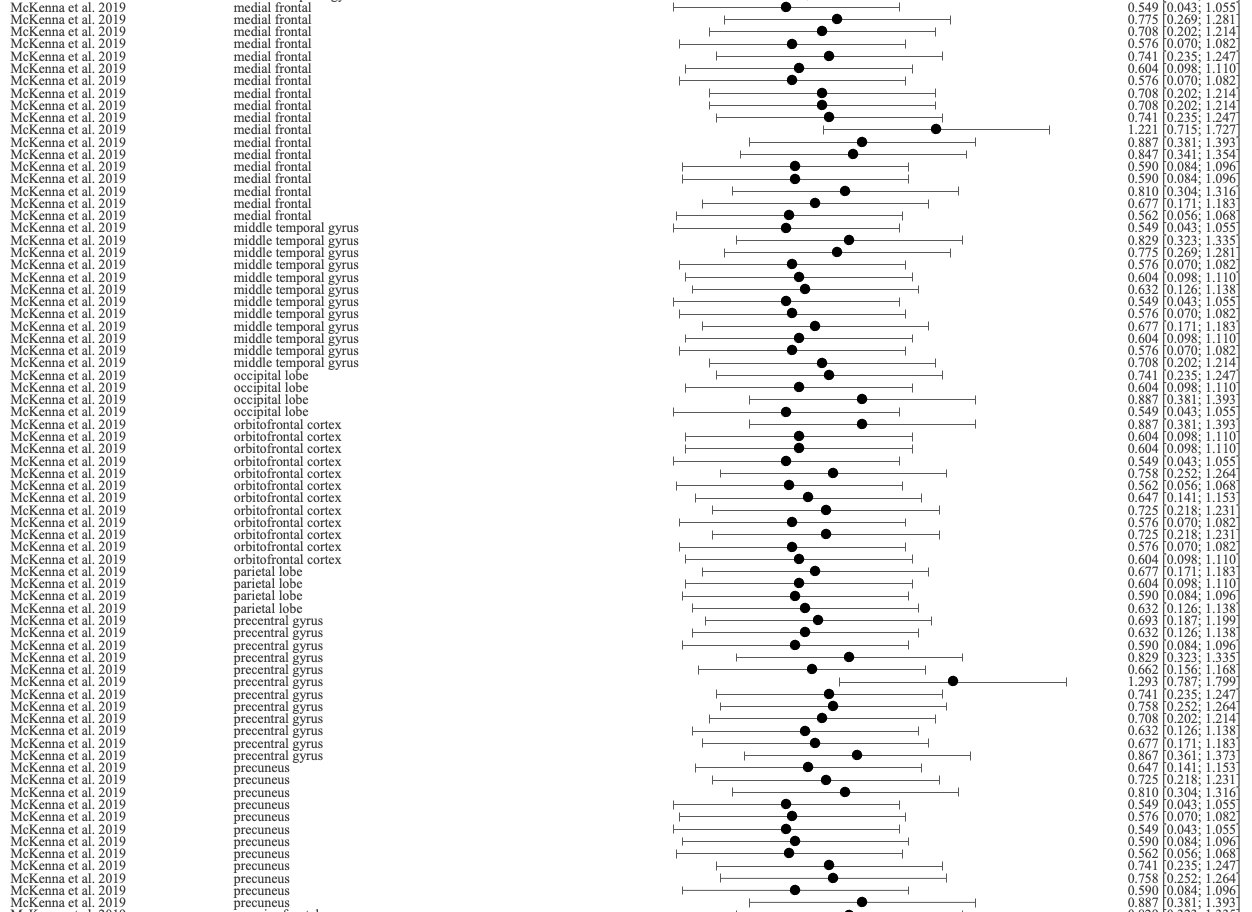

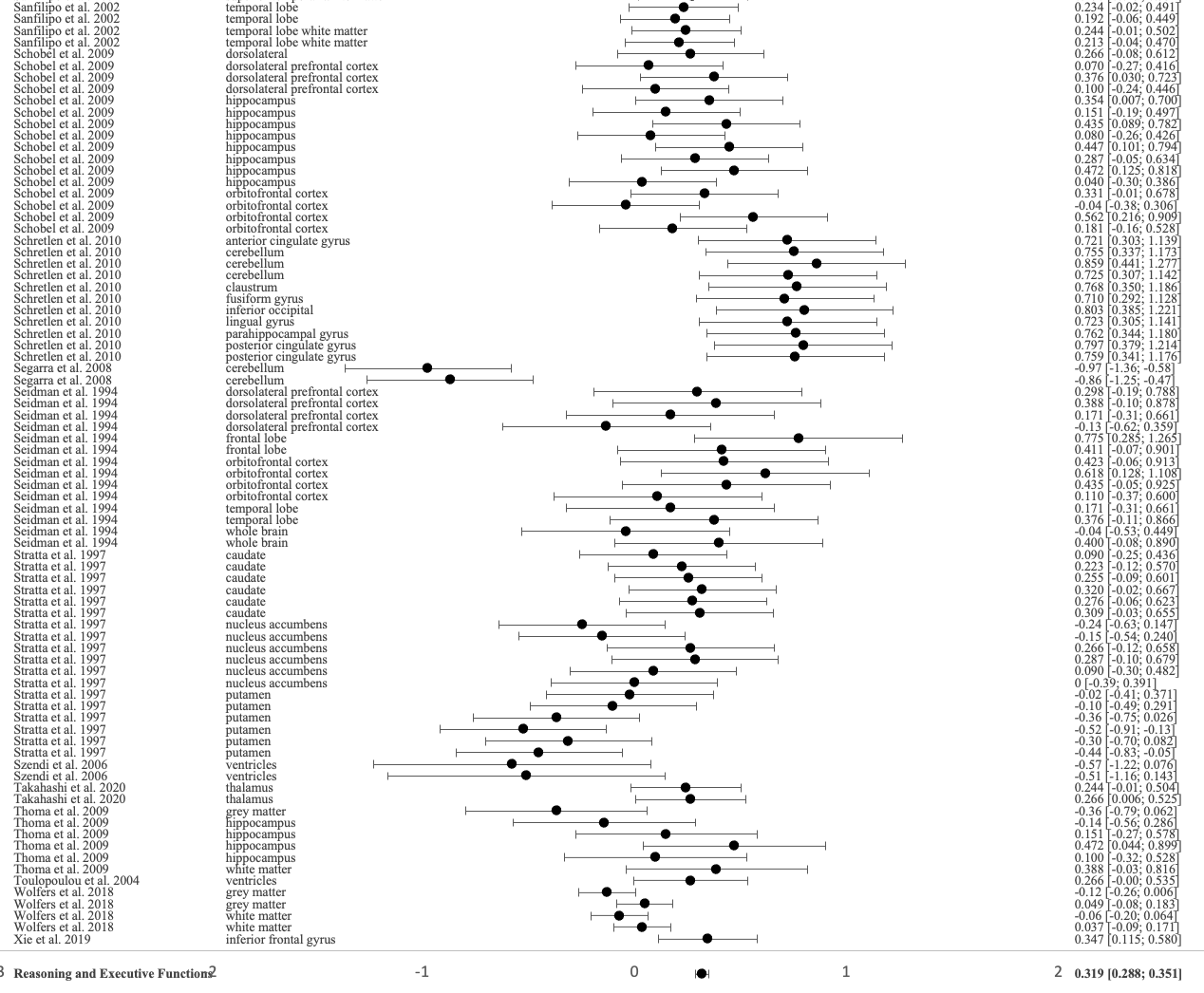

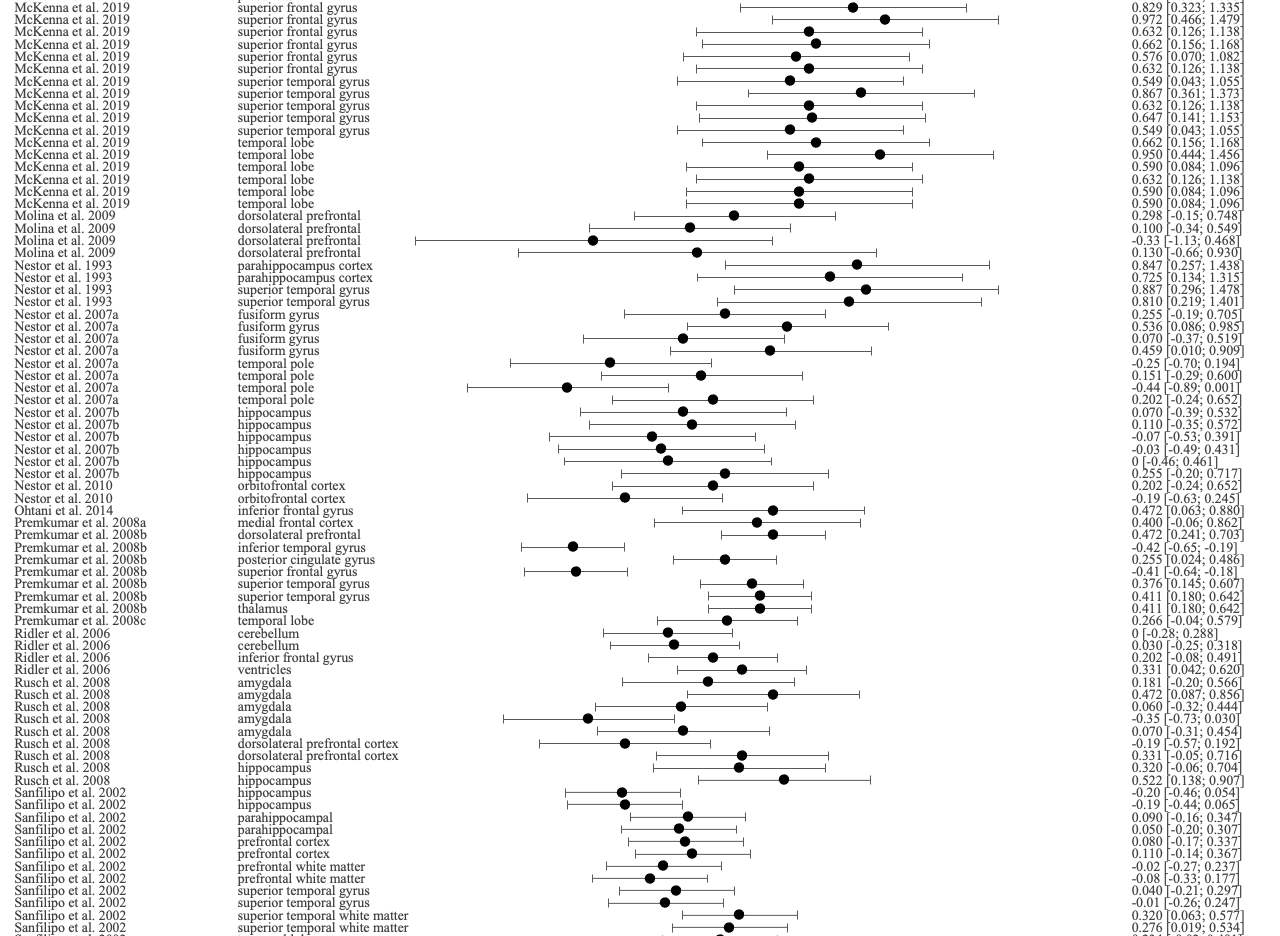

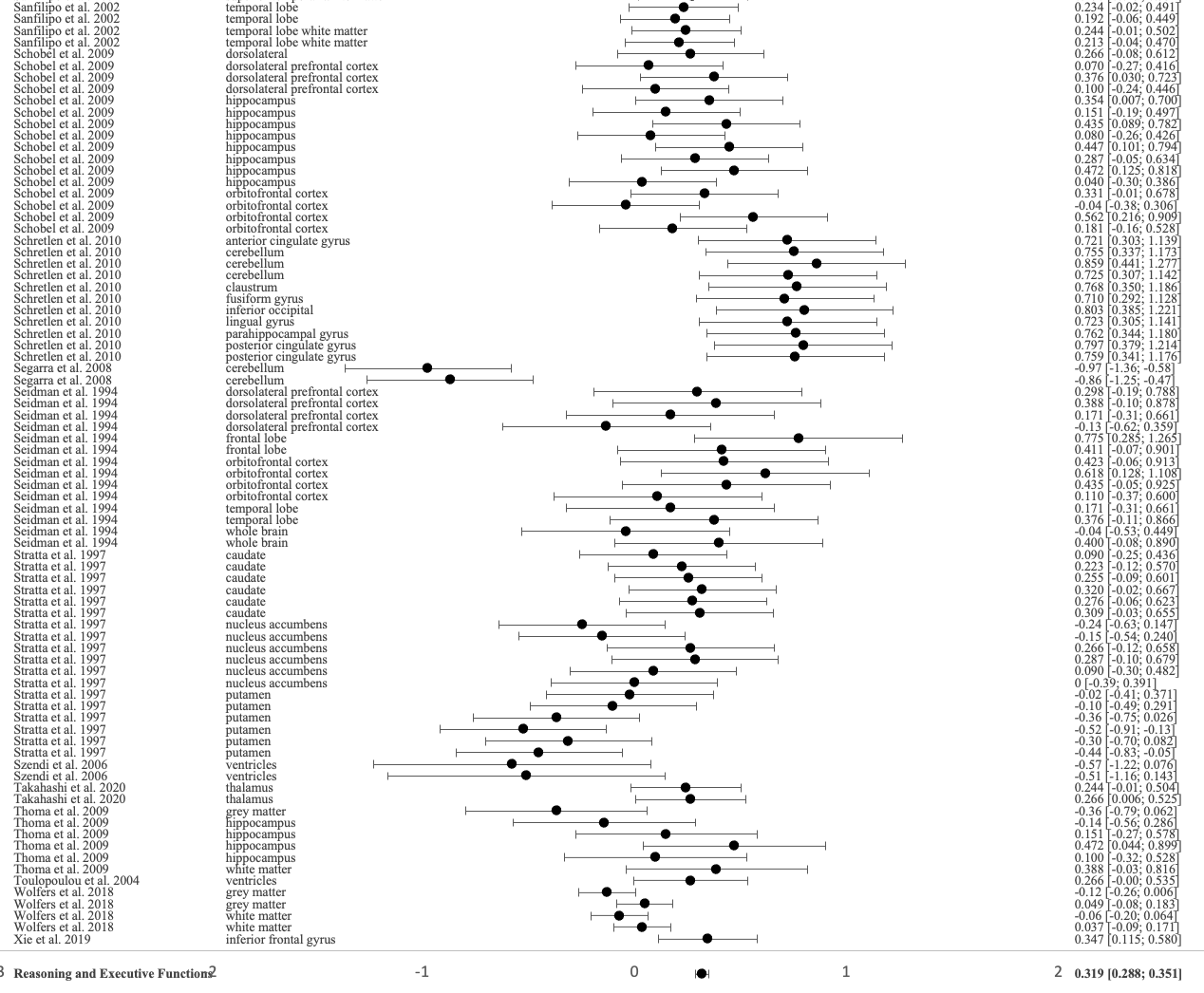

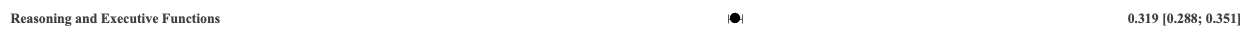

-1	0	1

### **Figure S8.** Forest plot of the correlation between social cognition and all brain structures reported in the included studies

-1	0	1

**Study**

**Brain region**

**Fisher’s z [95% CI]**

|  | data |
| --- | --- |
| 0.00474577 | 118 |
| -0.079645 | 117 |
| -0.018033 | 116 |
| 0.175639 | 115 |
| 0.0142369 | 114 |
| -0.0265733 | 113 |
| 0.0862663 | 112 |
| -0.0682863 | 111 |
| 0.0578663 | 110 |
| 0.0455438 | 109 |
| -0.0957187 | 108 |
| 0.0237267 | 107 |
| 0.0132878 | 106 |
| -0.00379662 | 105 |
| 0.142837 | 104 |
| -0.0360602 | 103 |
| 0.0654452 | 102 |
| -0.016135 | 101 |
| 0.0777526 | 100 |
| -0.0947739 | 99 |
| 0.712113 | 98 |
| 0.561743 | 97 |
| 0.422459 | 96 |
| 0.463583 | 95 |
| 0.441636 | 94 |
| 0.522984 | 93 |
| 0.479696 | 92 |
| 0.266108 | 91 |
| 0.522984 | 90 |
| 0.192337 | 89 |
| 0.244774 | 88 |
| 0.13074 | 87 |
| 0.140926 | 86 |
| 0.879929 | 85 |
| 0.892861 | 84 |
| 0.779072 | 83 |
| 0.816859 | 82 |
| 1.09836 | 81 |
| 1.00407 | 80 |
| 1.2387 | 79 |
| 0.847077 | 78 |
| 0.84375 | 77 |
| 1.26298 | 76 |
| 0.966486 | 75 |
| 0.871787 | 74 |
| 0.937199 | 73 |
| 0.937199 | 72 |
| 0.881552 | 71 |
| 0.837073 | 70 |
| 0.875049 | 69 |
| 0.946516 | 68 |
| 1.04061 | 67 |
| 0.868516 | 66 |
| 0.808354 | 65 |
| 1.08451 | 64 |
| 0.773846 | 63 |
| 0.889639 | 62 |
| 1.37294 | 61 |
| 0.888025 | 60 |
| 1.01293 | 59 |
| 0.842083 | 58 |
| 0.861954 | 57 |
| 0.961904 | 56 |
| 0.830366 | 55 |
| 0.918371 | 54 |
| 0.918371 | 53 |
| 0.969532 | 52 |
| 1.17055 | 51 |
| 0.811761 | 50 |
| 0.894469 | 49 |
| 0.879929 | 48 |
| 1.00407 | 47 |
| 0.873419 | 46 |
| 1.03048 | 45 |
| 1.18987 | 44 |
| 0.870152 | 43 |
| 0.902482 | 42 |
| 0.929385 | 41 |
| 0.987664 | 40 |
| 0.938756 | 39 |
| 1.06487 | 38 |
| 0.95577 | 37 |
| 1.00851 | 36 |
| 0.848737 | 35 |
| 0.951151 | 34 |
| 0.865239 | 33 |
| -0.0500417 | 32 |
| -0.13074 | 31 |
| 0.177879 | 30 |
| 0.170188 | 29 |
| -0.120581 | 28 |
| -0.192337 | 27 |
| 0.140926 | 26 |
| 0.110447 | 25 |
| 0.202733 | 24 |
| -0.0500417 | 23 |
| 0 | 22 |
| -0.0902442 | 21 |
| 0.0400214 | 20 |
| 0.161387 | 19 |
| 0.13074 | 18 |
| 0.140926 | 17 |
| 0.0600722 | 16 |
| 0.120581 | 15 |
| 0.677666 | 14 |
| 0.53606 | 13 |
| 0.4847 | 12 |
| 4.22 | 11 |
| 3.61 | 10 |
| 4.35 | 9 |
| 0.435611 | 8 |
| 0.320545 | 7 |
| 0.354093 | 6 |
| 3.66 | 5 |
| 3.51 | 4 |
| 0.244774 | 3 |
| 0.354093 | 2 |
| 0.502397 | 1 |
| 0.669613 | -2 |

|  | studies |
| --- | --- |
| -3 | 118 |
| -3 | 117 |
| -3 | 116 |
| -3 | 115 |
| -3 | 114 |
| -3 | 113 |
| -3 | 112 |
| -3 | 111 |
| -3 | 110 |
| -3 | 109 |
| -3 | 108 |
| -3 | 107 |
| -3 | 106 |
| -3 | 105 |
| -3 | 104 |
| -3 | 103 |
| -3 | 102 |
| -3 | 101 |
| -3 | 100 |
| -3 | 99 |
| -3 | 98 |
| -3 | 97 |
| -3 | 96 |
| -3 | 95 |
| -3 | 94 |
| -3 | 93 |
| -3 | 92 |
| -3 | 91 |
| -3 | 90 |
| -3 | 89 |
| -3 | 88 |
| -3 | 87 |
| -3 | 86 |
| -3 | 85 |
| -3 | 84 |
| -3 | 83 |
| -3 | 82 |
| -3 | 81 |
| -3 | 80 |
| -3 | 79 |
| -3 | 78 |
| -3 | 77 |
| -3 | 76 |
| -3 | 75 |
| -3 | 74 |
| -3 | 73 |
| -3 | 72 |
| -3 | 71 |
| -3 | 70 |
| -3 | 69 |
| -3 | 68 |
| -3 | 67 |
| -3 | 66 |
| -3 | 65 |
| -3 | 64 |
| -3 | 63 |
| -3 | 62 |
| -3 | 61 |
| -3 | 60 |
| -3 | 59 |
| -3 | 58 |
| -3 | 57 |
| -3 | 56 |
| -3 | 55 |
| -3 | 54 |
| -3 | 53 |
| -3 | 52 |
| -3 | 51 |
| -3 | 50 |
| -3 | 49 |
| -3 | 48 |
| -3 | 47 |
| -3 | 46 |
| -3 | 45 |
| -3 | 44 |
| -3 | 43 |
| -3 | 42 |
| -3 | 41 |
| -3 | 40 |
| -3 | 39 |
| -3 | 38 |
| -3 | 37 |
| -3 | 36 |
| -3 | 35 |
| -3 | 34 |
| -3 | 33 |
| -3 | 32 |
| -3 | 31 |
| -3 | 30 |
| -3 | 29 |
| -3 | 28 |
| -3 | 27 |
| -3 | 26 |
| -3 | 25 |
| -3 | 24 |
| -3 | 23 |
| -3 | 22 |
| -3 | 21 |
| -3 | 20 |
| -3 | 19 |
| -3 | 18 |
| -3 | 17 |
| -3 | 16 |
| -3 | 15 |
| -3 | 14 |
| -3 | 13 |
| -3 | 12 |
| -3 | 11 |
| -3 | 10 |
| -3 | 9 |
| -3 | 8 |
| -3 | 7 |
| -3 | 6 |
| -3 | 5 |
| -3 | 4 |
| -3 | 3 |
| -3 | 2 |
| -3 | 1 |
| -3 | -2 |

|  | regions |
| --- | --- |
| -1.7 | 118 |
| -1.7 | 117 |
| -1.7 | 116 |
| -1.7 | 115 |
| -1.7 | 114 |
| -1.7 | 113 |
| -1.7 | 112 |
| -1.7 | 111 |
| -1.7 | 110 |
| -1.7 | 109 |
| -1.7 | 108 |
| -1.7 | 107 |
| -1.7 | 106 |
| -1.7 | 105 |
| -1.7 | 104 |
| -1.7 | 103 |
| -1.7 | 102 |
| -1.7 | 101 |
| -1.7 | 100 |
| -1.7 | 99 |
| -1.7 | 98 |
| -1.7 | 97 |
| -1.7 | 96 |
| -1.7 | 95 |
| -1.7 | 94 |
| -1.7 | 93 |
| -1.7 | 92 |
| -1.7 | 91 |
| -1.7 | 90 |
| -1.7 | 89 |
| -1.7 | 88 |
| -1.7 | 87 |
| -1.7 | 86 |
| -1.7 | 85 |
| -1.7 | 84 |
| -1.7 | 83 |
| -1.7 | 82 |
| -1.7 | 81 |
| -1.7 | 80 |
| -1.7 | 79 |
| -1.7 | 78 |
| -1.7 | 77 |
| -1.7 | 76 |
| -1.7 | 75 |
| -1.7 | 74 |
| -1.7 | 73 |
| -1.7 | 72 |
| -1.7 | 71 |
| -1.7 | 70 |
| -1.7 | 69 |
| -1.7 | 68 |
| -1.7 | 67 |
| -1.7 | 66 |
| -1.7 | 65 |
| -1.7 | 64 |
| -1.7 | 63 |
| -1.7 | 62 |
| -1.7 | 61 |
| -1.7 | 60 |
| -1.7 | 59 |
| -1.7 | 58 |
| -1.7 | 57 |
| -1.7 | 56 |
| -1.7 | 55 |
| -1.7 | 54 |
| -1.7 | 53 |
| -1.7 | 52 |
| -1.7 | 51 |
| -1.7 | 50 |
| -1.7 | 49 |
| -1.7 | 48 |
| -1.7 | 47 |
| -1.7 | 46 |
| -1.7 | 45 |
| -1.7 | 44 |
| -1.7 | 43 |
| -1.7 | 42 |
| -1.7 | 41 |
| -1.7 | 40 |
| -1.7 | 39 |
| -1.7 | 38 |
| -1.7 | 37 |
| -1.7 | 36 |
| -1.7 | 35 |
| -1.7 | 34 |
| -1.7 | 33 |
| -1.7 | 32 |
| -1.7 | 31 |
| -1.7 | 30 |
| -1.7 | 29 |
| -1.7 | 28 |
| -1.7 | 27 |
| -1.7 | 26 |
| -1.7 | 25 |
| -1.7 | 24 |
| -1.7 | 23 |
| -1.7 | 22 |
| -1.7 | 21 |
| -1.7 | 20 |
| -1.7 | 19 |
| -1.7 | 18 |
| -1.7 | 17 |
| -1.7 | 16 |
| -1.7 | 15 |
| -1.7 | 14 |
| -1.7 | 13 |
| -1.7 | 12 |
| -1.7 | 11 |
| -1.7 | 10 |
| -1.7 | 9 |
| -1.7 | 8 |
| -1.7 | 7 |
| -1.7 | 6 |
| -1.7 | 5 |
| -1.7 | 4 |
| -1.7 | 3 |
| -1.7 | 2 |
| -1.7 | 1 |
| -1.7 | -2 |

|  | value |
| --- | --- |
| 2 | 118 |
| 2 | 117 |
| 2 | 116 |
| 2 | 115 |
| 2 | 114 |
| 2 | 113 |
| 2 | 112 |
| 2 | 111 |
| 2 | 110 |
| 2 | 109 |
| 2 | 108 |
| 2 | 107 |
| 2 | 106 |
| 2 | 105 |
| 2 | 104 |
| 2 | 103 |
| 2 | 102 |
| 2 | 101 |
| 2 | 100 |
| 2 | 99 |
| 2 | 98 |
| 2 | 97 |
| 2 | 96 |
| 2 | 95 |
| 2 | 94 |
| 2 | 93 |
| 2 | 92 |
| 2 | 91 |
| 2 | 90 |
| 2 | 89 |
| 2 | 88 |
| 2 | 87 |
| 2 | 86 |
| 2 | 85 |
| 2 | 84 |
| 2 | 83 |
| 2 | 82 |
| 2 | 81 |
| 2 | 80 |
| 2 | 79 |
| 2 | 78 |
| 2 | 77 |
| 2 | 76 |
| 2 | 75 |
| 2 | 74 |
| 2 | 73 |
| 2 | 72 |
| 2 | 71 |
| 2 | 70 |
| 2 | 69 |
| 2 | 68 |
| 2 | 67 |
| 2 | 66 |
| 2 | 65 |
| 2 | 64 |
| 2 | 63 |
| 2 | 62 |
| 2 | 61 |
| 2 | 60 |
| 2 | 59 |
| 2 | 58 |
| 2 | 57 |
| 2 | 56 |
| 2 | 55 |
| 2 | 54 |
| 2 | 53 |
| 2 | 52 |
| 2 | 51 |
| 2 | 50 |
| 2 | 49 |
| 2 | 48 |
| 2 | 47 |
| 2 | 46 |
| 2 | 45 |
| 2 | 44 |
| 2 | 43 |
| 2 | 42 |
| 2 | 41 |
| 2 | 40 |
| 2 | 39 |
| 2 | 38 |
| 2 | 37 |
| 2 | 36 |
| 2 | 35 |
| 2 | 34 |
| 2 | 33 |
| 2 | 32 |
| 2 | 31 |
| 2 | 30 |
| 2 | 29 |
| 2 | 28 |
| 2 | 27 |
| 2 | 26 |
| 2 | 25 |
| 2 | 24 |
| 2 | 23 |
| 2 | 22 |
| 2 | 21 |
| 2 | 20 |
| 2 | 19 |
| 2 | 18 |
| 2 | 17 |
| 2 | 16 |
| 2 | 15 |
| 2 | 14 |
| 2 | 13 |
| 2 | 12 |
| 2 | 11 |
| 2 | 10 |
| 2 | 9 |
| 2 | 8 |
| 2 | 7 |
| 2 | 6 |
| 2 | 5 |
| 2 | 4 |
| 2 | 3 |
| 2 | 2 |
| 2 | 1 |
| 2 | -2 |

### **Figure S9.** Forest plot of the correlation between verbal fluency and all brain structures reported in the included studies

-1	0	1

**Fisher’s z [95% CI]**

**Brain region**

**Study**

|  | data |
| --- | --- |
| 0.384952 | 72 |
| 0.758174 | 71 |
| 0.0600722 | 70 |
| -0.0200027 | 69 |
| -0.0400214 | 68 |
| -0.0902442 | 67 |
| -0.0500417 | 66 |
| -0.0600722 | 65 |
| -0.13074 | 64 |
| -0.0701147 | 63 |
| 0.120581 | 62 |
| 0.0801713 | 61 |
| -0.001 | 60 |
| -0.001 | 59 |
| -0.331647 | 58 |
| -0.202733 | 57 |
| 0.276864 | 56 |
| 0.342828 | 55 |
| 0.677666 | 54 |
| 0.30952 | 53 |
| 0.213171 | 52 |
| 0.255413 | 51 |
| -0.181983 | 50 |
| 0.0701147 | 49 |
| 0.0200027 | 48 |
| 0.244774 | 47 |
| 0.365444 | 46 |
| 0.0400214 | 45 |
| 0.120581 | 44 |
| 0.15114 | 43 |
| 0.202733 | 42 |
| 0.320545 | 41 |
| 0.13074 | 40 |
| 0.13074 | 39 |
| 0.0500417 | 38 |
| 0 | 37 |
| 0.140926 | 36 |
| 0.4118 | 35 |
| 0.40006 | 34 |
| 0.15114 | 33 |
| 0.266108 | 32 |
| 0.213171 | 31 |
| 0.56273 | 30 |
| 0.459897 | 29 |
| 0.100335 | 28 |
| -0.0701147 | 27 |
| -0.0500417 | 26 |
| 0.56273 | 25 |
| 0.435611 | 24 |
| -0.00300001 | 23 |
| -0.140926 | 22 |
| -0.181983 | 21 |
| -0.244774 | 20 |
| -0.0701147 | 19 |
| -0.244774 | 18 |
| -0.140926 | 17 |
| 0.100335 | 16 |
| 0.0500417 | 15 |
| -0.0902442 | 14 |
| -0.192337 | 13 |
| -0.0701147 | 12 |
| -0.030009 | 11 |
| -0.15114 | 10 |
| -0.100335 | 9 |
| -0.100335 | 8 |
| -0.030009 | 7 |
| 0.315023 | 6 |
| 0.258615 | 5 |
| 0.15114 | 4 |
| 0.522984 | 3 |
| 0.522984 | 2 |
| 0.301845 | 1 |
| 0.0869188 | -2 |

|  | studies |
| --- | --- |
| -3 | 72 |
| -3 | 71 |
| -3 | 70 |
| -3 | 69 |
| -3 | 68 |
| -3 | 67 |
| -3 | 66 |
| -3 | 65 |
| -3 | 64 |
| -3 | 63 |
| -3 | 62 |
| -3 | 61 |
| -3 | 60 |
| -3 | 59 |
| -3 | 58 |
| -3 | 57 |
| -3 | 56 |
| -3 | 55 |
| -3 | 54 |
| -3 | 53 |
| -3 | 52 |
| -3 | 51 |
| -3 | 50 |
| -3 | 49 |
| -3 | 48 |
| -3 | 47 |
| -3 | 46 |
| -3 | 45 |
| -3 | 44 |
| -3 | 43 |
| -3 | 42 |
| -3 | 41 |
| -3 | 40 |
| -3 | 39 |
| -3 | 38 |
| -3 | 37 |
| -3 | 36 |
| -3 | 35 |
| -3 | 34 |
| -3 | 33 |
| -3 | 32 |
| -3 | 31 |
| -3 | 30 |
| -3 | 29 |
| -3 | 28 |
| -3 | 27 |
| -3 | 26 |
| -3 | 25 |
| -3 | 24 |
| -3 | 23 |
| -3 | 22 |
| -3 | 21 |
| -3 | 20 |
| -3 | 19 |
| -3 | 18 |
| -3 | 17 |
| -3 | 16 |
| -3 | 15 |
| -3 | 14 |
| -3 | 13 |
| -3 | 12 |
| -3 | 11 |
| -3 | 10 |
| -3 | 9 |
| -3 | 8 |
| -3 | 7 |
| -3 | 6 |
| -3 | 5 |
| -3 | 4 |
| -3 | 3 |
| -3 | 2 |
| -3 | 1 |
| -3 | -2 |

|  | regions |
| --- | --- |
| -2 | 72 |
| -2 | 71 |
| -2 | 70 |
| -2 | 69 |
| -2 | 68 |
| -2 | 67 |
| -2 | 66 |
| -2 | 65 |
| -2 | 64 |
| -2 | 63 |
| -2 | 62 |
| -2 | 61 |
| -2 | 60 |
| -2 | 59 |
| -2 | 58 |
| -2 | 57 |
| -2 | 56 |
| -2 | 55 |
| -2 | 54 |
| -2 | 53 |
| -2 | 52 |
| -2 | 51 |
| -2 | 50 |
| -2 | 49 |
| -2 | 48 |
| -2 | 47 |
| -2 | 46 |
| -2 | 45 |
| -2 | 44 |
| -2 | 43 |
| -2 | 42 |
| -2 | 41 |
| -2 | 40 |
| -2 | 39 |
| -2 | 38 |
| -2 | 37 |
| -2 | 36 |
| -2 | 35 |
| -2 | 34 |
| -2 | 33 |
| -2 | 32 |
| -2 | 31 |
| -2 | 30 |
| -2 | 29 |
| -2 | 28 |
| -2 | 27 |
| -2 | 26 |
| -2 | 25 |
| -2 | 24 |
| -2 | 23 |
| -2 | 22 |
| -2 | 21 |
| -2 | 20 |
| -2 | 19 |
| -2 | 18 |
| -2 | 17 |
| -2 | 16 |
| -2 | 15 |
| -2 | 14 |
| -2 | 13 |
| -2 | 12 |
| -2 | 11 |
| -2 | 10 |
| -2 | 9 |
| -2 | 8 |
| -2 | 7 |
| -2 | 6 |
| -2 | 5 |
| -2 | 4 |
| -2 | 3 |
| -2 | 2 |
| -2 | 1 |
| -2 | -2 |

|  | value |
| --- | --- |
| 2 | 72 |
| 2 | 71 |
| 2 | 70 |
| 2 | 69 |
| 2 | 68 |
| 2 | 67 |
| 2 | 66 |
| 2 | 65 |
| 2 | 64 |
| 2 | 63 |
| 2 | 62 |
| 2 | 61 |
| 2 | 60 |
| 2 | 59 |
| 2 | 58 |
| 2 | 57 |
| 2 | 56 |
| 2 | 55 |
| 2 | 54 |
| 2 | 53 |
| 2 | 52 |
| 2 | 51 |
| 2 | 50 |
| 2 | 49 |
| 2 | 48 |
| 2 | 47 |
| 2 | 46 |
| 2 | 45 |
| 2 | 44 |
| 2 | 43 |
| 2 | 42 |
| 2 | 41 |
| 2 | 40 |
| 2 | 39 |
| 2 | 38 |
| 2 | 37 |
| 2 | 36 |
| 2 | 35 |
| 2 | 34 |
| 2 | 33 |
| 2 | 32 |
| 2 | 31 |
| 2 | 30 |
| 2 | 29 |
| 2 | 28 |
| 2 | 27 |
| 2 | 26 |
| 2 | 25 |
| 2 | 24 |
| 2 | 23 |
| 2 | 22 |
| 2 | 21 |
| 2 | 20 |
| 2 | 19 |
| 2 | 18 |
| 2 | 17 |
| 2 | 16 |
| 2 | 15 |
| 2 | 14 |
| 2 | 13 |
| 2 | 12 |
| 2 | 11 |
| 2 | 10 |
| 2 | 9 |
| 2 | 8 |
| 2 | 7 |
| 2 | 6 |
| 2 | 5 |
| 2 | 4 |
| 2 | 3 |
| 2 | 2 |
| 2 | 1 |
| 2 | -2 |

### **Figure S10.** Forest plot of the correlation between emotional processing tasks of social cognition and all brain structures reported in included studies

**Study**

**Brain region**

**Fisher’s z [95% CI]**

|  | data |
| --- | --- |
| 0.00474577 | 49 |
| -0.079645 | 48 |
| -0.018033 | 47 |
| 0.175639 | 46 |
| 0.0142369 | 45 |
| -0.0265733 | 44 |
| 0.0862663 | 43 |
| -0.0682863 | 42 |
| 0.0578663 | 41 |
| 0.0455438 | 40 |
| 0.712113 | 39 |
| 0.561743 | 38 |
| 0.422459 | 37 |
| 0.463583 | 36 |
| 0.441636 | 35 |
| 0.522984 | 34 |
| 0.479696 | 33 |
| -0.0500417 | 32 |
| -0.13074 | 31 |
| 0.177879 | 30 |
| 0.170188 | 29 |
| -0.120581 | 28 |
| -0.192337 | 27 |
| 0.140926 | 26 |
| 0.110447 | 25 |
| 0.202733 | 24 |
| -0.0500417 | 23 |
| 0 | 22 |
| -0.0902442 | 21 |
| 0.0400214 | 20 |
| 0.161387 | 19 |
| 0.13074 | 18 |
| 0.140926 | 17 |
| 0.0600722 | 16 |
| 0.120581 | 15 |
| 0.677666 | 14 |
| 0.53606 | 13 |
| 0.4847 | 12 |
| 4.22 | 11 |
| 3.61 | 10 |
| 4.35 | 9 |
| 0.435611 | 8 |
| 0.320545 | 7 |
| 0.354093 | 6 |
| 3.66 | 5 |
| 3.51 | 4 |
| 0.244774 | 3 |
| 0.354093 | 2 |
| 0.502397 | 1 |

|  | studies |
| --- | --- |
| -3 | 49 |
| -3 | 48 |
| -3 | 47 |
| -3 | 46 |
| -3 | 45 |
| -3 | 44 |
| -3 | 43 |
| -3 | 42 |
| -3 | 41 |
| -3 | 40 |
| -3 | 39 |
| -3 | 38 |
| -3 | 37 |
| -3 | 36 |
| -3 | 35 |
| -3 | 34 |
| -3 | 33 |
| -3 | 32 |
| -3 | 31 |
| -3 | 30 |
| -3 | 29 |
| -3 | 28 |
| -3 | 27 |
| -3 | 26 |
| -3 | 25 |
| -3 | 24 |
| -3 | 23 |
| -3 | 22 |
| -3 | 21 |
| -3 | 20 |
| -3 | 19 |
| -3 | 18 |
| -3 | 17 |
| -3 | 16 |
| -3 | 15 |
| -3 | 14 |
| -3 | 13 |
| -3 | 12 |
| -3 | 11 |
| -3 | 10 |
| -3 | 9 |
| -3 | 8 |
| -3 | 7 |
| -3 | 6 |
| -3 | 5 |
| -3 | 4 |
| -3 | 3 |
| -3 | 2 |
| -3 | 1 |

|  | regions |
| --- | --- |
| -1.7 | 49 |
| -1.7 | 48 |
| -1.7 | 47 |
| -1.7 | 46 |
| -1.7 | 45 |
| -1.7 | 44 |
| -1.7 | 43 |
| -1.7 | 42 |
| -1.7 | 41 |
| -1.7 | 40 |
| -1.7 | 39 |
| -1.7 | 38 |
| -1.7 | 37 |
| -1.7 | 36 |
| -1.7 | 35 |
| -1.7 | 34 |
| -1.7 | 33 |
| -1.7 | 32 |
| -1.7 | 31 |
| -1.7 | 30 |
| -1.7 | 29 |
| -1.7 | 28 |
| -1.7 | 27 |
| -1.7 | 26 |
| -1.7 | 25 |
| -1.7 | 24 |
| -1.7 | 23 |
| -1.7 | 22 |
| -1.7 | 21 |
| -1.7 | 20 |
| -1.7 | 19 |
| -1.7 | 18 |
| -1.7 | 17 |
| -1.7 | 16 |
| -1.7 | 15 |
| -1.7 | 14 |
| -1.7 | 13 |
| -1.7 | 12 |
| -1.7 | 11 |
| -1.7 | 10 |
| -1.7 | 9 |
| -1.7 | 8 |
| -1.7 | 7 |
| -1.7 | 6 |
| -1.7 | 5 |
| -1.7 | 4 |
| -1.7 | 3 |
| -1.7 | 2 |
| -1.7 | 1 |

|  | value |
| --- | --- |
| 2 | 49 |
| 2 | 48 |
| 2 | 47 |
| 2 | 46 |
| 2 | 45 |
| 2 | 44 |
| 2 | 43 |
| 2 | 42 |
| 2 | 41 |
| 2 | 40 |
| 2 | 39 |
| 2 | 38 |
| 2 | 37 |
| 2 | 36 |
| 2 | 35 |
| 2 | 34 |
| 2 | 33 |
| 2 | 32 |
| 2 | 31 |
| 2 | 30 |
| 2 | 29 |
| 2 | 28 |
| 2 | 27 |
| 2 | 26 |
| 2 | 25 |
| 2 | 24 |
| 2 | 23 |
| 2 | 22 |
| 2 | 21 |
| 2 | 20 |
| 2 | 19 |
| 2 | 18 |
| 2 | 17 |
| 2 | 16 |
| 2 | 15 |
| 2 | 14 |
| 2 | 13 |
| 2 | 12 |
| 2 | 11 |
| 2 | 10 |
| 2 | 9 |
| 2 | 8 |
| 2 | 7 |
| 2 | 6 |
| 2 | 5 |
| 2 | 4 |
| 2 | 3 |
| 2 | 2 |
| 2 | 1 |

### **Figure S11.** Forest plot of the correlation between theory of mind tasks of social cognition and all brain structures reported in included studies

**Fisher’s z [95% CI]**

**Brain region**

**Study**

|  | data |
| --- | --- |
| -0.0957187 | 69 |
| 0.0237267 | 68 |
| 0.0132878 | 67 |
| -0.00379662 | 66 |
| 0.142837 | 65 |
| -0.0360602 | 64 |
| 0.0654452 | 63 |
| -0.016135 | 62 |
| 0.0777526 | 61 |
| -0.0947739 | 60 |
| 0.266108 | 59 |
| 0.522984 | 58 |
| 0.192337 | 57 |
| 0.244774 | 56 |
| 0.13074 | 55 |
| 0.140926 | 54 |
| 0.879929 | 53 |
| 0.892861 | 52 |
| 0.779072 | 51 |
| 0.816859 | 50 |
| 1.09836 | 49 |
| 1.00407 | 48 |
| 1.2387 | 47 |
| 0.847077 | 46 |
| 0.84375 | 45 |
| 1.26298 | 44 |
| 0.966486 | 43 |
| 0.871787 | 42 |
| 0.937199 | 41 |
| 0.937199 | 40 |
| 0.881552 | 39 |
| 0.837073 | 38 |
| 0.875049 | 37 |
| 0.946516 | 36 |
| 1.04061 | 35 |
| 0.868516 | 34 |
| 0.808354 | 33 |
| 1.08451 | 32 |
| 0.773846 | 31 |
| 0.889639 | 30 |
| 1.37294 | 29 |
| 0.888025 | 28 |
| 1.01293 | 27 |
| 0.842083 | 26 |
| 0.861954 | 25 |
| 0.961904 | 24 |
| 0.830366 | 23 |
| 0.918371 | 22 |
| 0.918371 | 21 |
| 0.969532 | 20 |
| 1.17055 | 19 |
| 0.811761 | 18 |
| 0.894469 | 17 |
| 0.879929 | 16 |
| 1.00407 | 15 |
| 0.873419 | 14 |
| 1.03048 | 13 |
| 1.18987 | 12 |
| 0.870152 | 11 |
| 0.902482 | 10 |
| 0.929385 | 9 |
| 0.987664 | 8 |
| 0.938756 | 7 |
| 1.06487 | 6 |
| 0.95577 | 5 |
| 1.00851 | 4 |
| 0.848737 | 3 |
| 0.951151 | 2 |
| 0.865239 | 1 |

|  | studies |
| --- | --- |
| -3 | 69 |
| -3 | 68 |
| -3 | 67 |
| -3 | 66 |
| -3 | 65 |
| -3 | 64 |
| -3 | 63 |
| -3 | 62 |
| -3 | 61 |
| -3 | 60 |
| -3 | 59 |
| -3 | 58 |
| -3 | 57 |
| -3 | 56 |
| -3 | 55 |
| -3 | 54 |
| -3 | 53 |
| -3 | 52 |
| -3 | 51 |
| -3 | 50 |
| -3 | 49 |
| -3 | 48 |
| -3 | 47 |
| -3 | 46 |
| -3 | 45 |
| -3 | 44 |
| -3 | 43 |
| -3 | 42 |
| -3 | 41 |
| -3 | 40 |
| -3 | 39 |
| -3 | 38 |
| -3 | 37 |
| -3 | 36 |
| -3 | 35 |
| -3 | 34 |
| -3 | 33 |
| -3 | 32 |
| -3 | 31 |
| -3 | 30 |
| -3 | 29 |
| -3 | 28 |
| -3 | 27 |
| -3 | 26 |
| -3 | 25 |
| -3 | 24 |
| -3 | 23 |
| -3 | 22 |
| -3 | 21 |
| -3 | 20 |
| -3 | 19 |
| -3 | 18 |
| -3 | 17 |
| -3 | 16 |
| -3 | 15 |
| -3 | 14 |
| -3 | 13 |
| -3 | 12 |
| -3 | 11 |
| -3 | 10 |
| -3 | 9 |
| -3 | 8 |
| -3 | 7 |
| -3 | 6 |
| -3 | 5 |
| -3 | 4 |
| -3 | 3 |
| -3 | 2 |
| -3 | 1 |

|  | regions |
| --- | --- |
| -1.7 | 69 |
| -1.7 | 68 |
| -1.7 | 67 |
| -1.7 | 66 |
| -1.7 | 65 |
| -1.7 | 64 |
| -1.7 | 63 |
| -1.7 | 62 |
| -1.7 | 61 |
| -1.7 | 60 |
| -1.7 | 59 |
| -1.7 | 58 |
| -1.7 | 57 |
| -1.7 | 56 |
| -1.7 | 55 |
| -1.7 | 54 |
| -1.7 | 53 |
| -1.7 | 52 |
| -1.7 | 51 |
| -1.7 | 50 |
| -1.7 | 49 |
| -1.7 | 48 |
| -1.7 | 47 |
| -1.7 | 46 |
| -1.7 | 45 |
| -1.7 | 44 |
| -1.7 | 43 |
| -1.7 | 42 |
| -1.7 | 41 |
| -1.7 | 40 |
| -1.7 | 39 |
| -1.7 | 38 |
| -1.7 | 37 |
| -1.7 | 36 |
| -1.7 | 35 |
| -1.7 | 34 |
| -1.7 | 33 |
| -1.7 | 32 |
| -1.7 | 31 |
| -1.7 | 30 |
| -1.7 | 29 |
| -1.7 | 28 |
| -1.7 | 27 |
| -1.7 | 26 |
| -1.7 | 25 |
| -1.7 | 24 |
| -1.7 | 23 |
| -1.7 | 22 |
| -1.7 | 21 |
| -1.7 | 20 |
| -1.7 | 19 |
| -1.7 | 18 |
| -1.7 | 17 |
| -1.7 | 16 |
| -1.7 | 15 |
| -1.7 | 14 |
| -1.7 | 13 |
| -1.7 | 12 |
| -1.7 | 11 |
| -1.7 | 10 |
| -1.7 | 9 |
| -1.7 | 8 |
| -1.7 | 7 |
| -1.7 | 6 |
| -1.7 | 5 |
| -1.7 | 4 |
| -1.7 | 3 |
| -1.7 | 2 |
| -1.7 | 1 |

|  | value |
| --- | --- |
| 2 | 69 |
| 2 | 68 |
| 2 | 67 |
| 2 | 66 |
| 2 | 65 |
| 2 | 64 |
| 2 | 63 |
| 2 | 62 |
| 2 | 61 |
| 2 | 60 |
| 2 | 59 |
| 2 | 58 |
| 2 | 57 |
| 2 | 56 |
| 2 | 55 |
| 2 | 54 |
| 2 | 53 |
| 2 | 52 |
| 2 | 51 |
| 2 | 50 |
| 2 | 49 |
| 2 | 48 |
| 2 | 47 |
| 2 | 46 |
| 2 | 45 |
| 2 | 44 |
| 2 | 43 |
| 2 | 42 |
| 2 | 41 |
| 2 | 40 |
| 2 | 39 |
| 2 | 38 |
| 2 | 37 |
| 2 | 36 |
| 2 | 35 |
| 2 | 34 |
| 2 | 33 |
| 2 | 32 |
| 2 | 31 |
| 2 | 30 |
| 2 | 29 |
| 2 | 28 |
| 2 | 27 |
| 2 | 26 |
| 2 | 25 |
| 2 | 24 |
| 2 | 23 |
| 2 | 22 |
| 2 | 21 |
| 2 | 20 |
| 2 | 19 |
| 2 | 18 |
| 2 | 17 |
| 2 | 16 |
| 2 | 15 |
| 2 | 14 |
| 2 | 13 |
| 2 | 12 |
| 2 | 11 |
| 2 | 10 |
| 2 | 9 |
| 2 | 8 |
| 2 | 7 |
| 2 | 6 |
| 2 | 5 |
| 2 | 4 |
| 2 | 3 |
| 2 | 2 |
| 2 | 1 |

### **Table S4.** Values of effects, corrected with FDR, observed between the eight cognitive domains and the seven networks as well as brain regions categorized in the networks.

| **#** | **Network or**  **Brain structure** | **Cognitive Domain** | **S, C, N** | **Fisher’s z [95% CI] random model** | **p-value** |
| --- | --- | --- | --- | --- | --- |
| **DMN** | | SP | **2, 5, 135** | **0.508 [0.280; 0.737]**  0.704 [0.346; 1.062]  0.423 [0.156; 0.690] | ***p < 0.001*** |
| 4 | Inferior temporal gyrus |  | 1, 2, 18 |  | *p < 0.001* |
| 8 | Temporal pole |  | 2, 3, 135 |  | *p = 0.002* |
| **DMN** | | VM | **3, 4, 161**  2, 3, 116  1, 1, 45 | **0.470 [-0.05; 0.946]** | **p = 0.052**  p = 0.165  *p = 0.007* |
| 3 | Inferior parietal lobule |  |  | 0.495 [-0.204; 1.194]  0.418 [0.115; 0.720] |  |
| 7 | Precuneus |  |  |  |  |
| **DMN** | | VisM | **1, 4, 22** | **-0.094 [-0.400; 0.212]**  -0.094 [-0.400; 0.212] | **p = 0.548**  p = 0.548 |
| 8 | Temporal Pole |  | 1, 4, 22 |  |  |
| **DMN** | | R&EF | **4, 69, 140**  2, 13, 43  1, 12, 18  2, 13, 93  1, 12, 18  2, 3, 100  1, 12, 18  1, 4, 22 | **0.620 [0.542; 0.699]**  0.694 [0.556; 0.832]  0.681 [0.535; 0.827]  0.593 [0.290; 0.897]  0.638 [0.492; 0.784]  0.571 [0.179; 0.963]  0.666 [0.520; 0.812]  0.330 [0.106; 0.555] | ***p < 0.001***  *p < 0.001*  *p < 0.001*  *p < 0.001*  *p < 0.001*  *p = 0.004*  *p < 0.001*  *p = 0.004* |
| 2 | Anterior cingulate gyrus |  |  |  |  |
| 3 | Inferior parietal lobule |  |  |  |  |
| 4 | Inferior temporal gyrus |  |  |  |  |
| 5 | Middle temporal gyrus |  |  |  |  |
| 6 | Posterior cingulate gyrus |  |  |  |  |
| 7 | Precuneus |  |  |  |  |
| 8 | Temporal pole |  |  |  |  |
| **DMN** | | SC | **5, 31, 157**  1, 1, 21  3, 18, 86  1, 1, 20  2, 5, 72  2, 5, 60  1, 1, 21 | **0.804 [0.356; 1.253]**  0.880 [0.418; 1.342]  0.569 [0.372; 0.765]  0.523 [0.048; 0.998]  1.722 [0.134; 3.309]  0.592 [0.153; 1.032]  0.951 [0.489; 1.413] | ***p < 0.001***  *p < 0.001*  *p < 0.001*  p = 0.031  p = 0.034  *p = 0.008*  *p < 0.001* |
| 1 | Angular gyrus |  |  |  |  |
| 2 | Anterior cingulate gyrus |  |  |  |  |
| 34 | IFG, pars orbitalis |  |  |  |  |
| 6 | Posterior cingulate gyrus |  |  |  |  |
| 7 | Precuneus |  |  |  |  |
| 8 | Temporal pole |  |  |  |  |
| **DMN** | | VF | **2, 2, 50**  1, 1, 18  1, 1, 32 | **0.447 [0.088; 0.806]**  0.678 [0.172; 1.184]  0.302 [-0.062; 0.666] | ***p = 0.015***  *p = 0.009*  p = 0.104 |
| 4 | Inferior temporal gyrus |  |  |  |  |
| 7 | Precuneus |  |  |  |  |
| **DAN** | | SP | **1, 1, 32**  1, 1, 32 | **0.576 [0.212; 0.940]**  0.576 [0.212; 0.940] | ***p = 0.002***  *p = 0.002* |
| 9 | IFG, pars opercularis |  |  |  |  |
| **DAN** | | ATT | **1, 2, 51**  1, 2, 51 | **0.352 [0.018; 0.687]**  0.352 [0.018; 0.687] | **p = 0.039**  p = 0.039 |
| 9 | IFG, pars opercularis |  |  |  |  |
| **DAN** | | WM | **2, 2, 135**  2, 2, 135 | **0.221 [0.048; 0.394]**  0.221 [0.048; 0.394] | ***p = 0.012***  *p = 0.012* |
| 9 | IFG, pars opercularis |  |  |  |  |
| **DAN** | | R&EF | **2, 13, 50**  2, 13, 50 | **0.729 [0.593; 0.864]**  0.729 [0.593; 0.864] | *p < 0.001*  *p < 0.001* |
| 9 | IFG, pars opercularis |  |  |  |  |
| **DAN** | | SC | **2, 3, 41**  2, 3, 41 | **0.929 [0.467; 1.391]**  0.929 [0.467; 1.391] | ***p < 0.001***  *p < 0.001* |
| 10 | Superior parietal lobule |  |  |  |  |
| **FPN** | | SP | **4, 7, 146**  2, 5, 57  2, 2, 89 | **0.337 [0.106; 0.568]**  0.017 [-0.199; 0.234]  0.616 [0.400; 0.831] | ***p = 0.004***  p = 0.875  *p < 0.001* |
| 11 | Dorsolateral prefrontal |  |  |  |  |
| 15 | Middle frontal gyrus |  |  |  |  |
| **FPN** | | ATT | **3, 10, 92**  2, 6, 41  1, 2, 51  1, 2, 51 | **0.123 [-0.061; 0.307]**  -0.114 [-0.386; 0.158]  0.288 [0.088; 0.488]  0.326 [0.126; 0.526] | **p = 0.190**  p = 0.411  *p = 0.005*  *p = 0.001* |
| 11 | Dorsolateral prefrontal |  |  |  |  |
| 13 | IFG, pars triangularis |  |  |  |  |
| 15 | Middle frontal gyrus |  |  |  |  |
| **FPN** | | WM | **8, 21, 168**  4, 14, 98  1, 4, 17  1, 1, 18  1, 1, 22  1, 1, 13 | **0.154 [0.012; 0.295]**  0.088 [-0.094; 0.271]  0.357 [0.095; 0.619]  -0.020 [-0.526; 0.486]  0.310 [-0.140; 0.759]  0.497 [-0.122; 1.117] | **p = 0.034**  p = 0.344  *p = 0.008*  p = 0.938  p = 0.177  p = 0.116 |
| 11 | Dorsolateral prefrontal |  |  |  |  |
| 12 | Inferior frontal gyrus |  |  |  |  |
| 13 | IFG, pars triangularis |  |  |  |  |
| 14 | Medial frontal gyrus |  |  |  |  |
| 15 | Middle frontal gyrus |  |  |  |  |
| **FPN** | | VM | **4, 12, 130**  2, 10, 41  2, 2, 89 | **0.440 [0.290; 0.590]**  0.373 [0.211; 0.535]  0.576 [0.182; 0.970] | ***p < 0.001***  *p < 0.001*  *p = 0.005* |
| 11 | Dorsolateral prefrontal |  |  |  |  |
|  | Middle frontal gyrus |  |  |  |  |
| **FPN** | | VisM | **4, 13, 142**  2, 10, 41  2, 3, 101 | **0.322 [0.202; 0.443]**  0.216 [0.054; 0.378]  0.465 [0.286; 0.645] | ***p < 0.001***  *p = 0.009*  *p < 0.001* |
| 11 | Dorsolateral prefrontal |  |  |  |  |
| 12 | Inferior frontal gyrus |  |  |  |  |
| **FPN** | | R&EF | **13, 51, 472**  6, 16, 194  3, 3, 149  1, 6, 18  4, 23, 86  1, 3, 57 | **0.411 [0.298; 0.523]**  0.240 [0.105; 0.375]  0.113 [-0.141; 0.368]  0.694 [0.556; 0.832]  0.643 [0.537; 0.749]  0.741 [0.517; 0.966] | ***p < 0.001***  *p < 0.001*  p = 0.383  *p < 0.001*  *p < 0.001*  *p < 0.001* |
| 11 | Dorsolateral prefrontal |  |  |  |  |
| 12 | Inferior frontal gyrus |  |  |  |  |
| 13 | IFG, pars triangularis |  |  |  |  |
| 14 | Medial frontal gyrus |  |  |  |  |
| 15 | Middle frontal gyrus |  |  |  |  |
| **FPN** | | SC | **5, 15, 266**  1, 2, 20  2, 3, 205  2, 3, 187  3, 7, 80 | **0.411 [0.237; 0.585]**  0.219 [-0.118; 0.555]  0.157 [0.029; 0.284]  0.640 [0.056; 1.224]  0.543 [0.178; 0.908] | ***p < 0.001***  p = 0.203  p = 0.016  p = 0.032  *p = 0.004* |
| 11 | Dorsolateral prefrontal |  |  |  |  |
| 12 | Inferior frontal gyrus |  |  |  |  |
| 13 | IFG, pars triangularis |  |  |  |  |
| 14 | Medial frontal gyrus |  |  |  |  |
| **LIM** | | SP | **5, 9, 159**  1, 2, 18  3, 5, 80  1, 2, 61 | **0.013 [-0.151; 0.177]**  0.563 [0.205; 0.921]  -0.041 [-0.236; 0.155]  -0.085 [-0.267; 0.097] | **p = 0.879**  *p = 0.002*  p = 0.684  p = 0.359 |
| 16 | Entorhinal cortex |  |  |  |  |
| 17 | Orbitofrontal cortex |  |  |  |  |
| 18 | Parahippocampal cortex |  |  |  |  |
| **LIM** | | ATT | **3, 5, 119**  3, 5, 119 | **0.150 [-0.272; 0.573]**  0.150 [-0.272; 0.573] | **p = 0.486**  p = 0.486 |
| 17 | Orbitofrontal cortex |  |  |  |  |
| **LIM** | | WM | **3, 5, 80**  3, 5, 80 | **0.025 [-0.252; 0.301]**  0.025 [-0.252; 0.301] | **p = 0.862**  p = 0.862 |
| 17 | Orbitofrontal cortex |  |  |  |  |
| **LIM** | | VM | **7, 22, 223**  1, 1, 32  4, 13, 85  2, 8, 106 | **0.129 [0.012; 0.245]**  -0.288 [-0.652; 0.076]  0.029 [-0.155; 0.212]  0.267 [0.165; 0.369] | **p = 0.031**  p = 0.121  p = 0.759  *p < 0.001* |
| 35 | Gyrus rectus |  |  |  |  |
| 17 | Orbitofrontal cortex |  |  |  |  |
| 18 | Parahippocampal cortex |  |  |  |  |
| **LIM** | | VisM | **5, 17, 160**  1, 1, 12  2, 10, 42  2, 6, 106 | **-0.049 [-0.139; 0.042]**  -0.460 [-1.113; 0.193]  0.028 [-0.119; 0.176]  -0.084 [-0.200; 0.032] | **p = 0.289**  p = 0.168  p = 0.707  p = 0.158 |
| 35 | Gyrus rectus |  |  |  |  |
| 17 | Orbitofrontal cortex |  |  |  |  |
| 18 | Parahippocampal cortex |  |  |  |  |
| **LIM** | | R&EF | **10, 33, 338**  5, 23, 166  5, 10, 172 | **0.365 [0.216; 0.513]**  0.423 [0.269; 0.577]  0.110 [-0.113; 0.334] | ***p < 0.001***  *p < 0.001*  p = 0.333 |
| 17 | Orbitofrontal cortex |  |  |  |  |
| 18 | Parahippocampal cortex |  |  |  |  |
| **LIM** | | SC | **2, 8, 72**  2, 2, 21  1, 6, 51 | **1.892 [0.547; 3.236]**  0.912 [0.585; 1.239]  2.215 [0.585; 3.845] | ***p = 0.006***  *p < 0.001*  *p = 0.007* |
| 17 | Orbitofrontal cortex |  |  |  |  |
| 18 | Parahippocampal cortex |  |  |  |  |
| **LIM** | | VF | **2, 4, 106**  2, 4, 106 | **-0.119 [-0.257; 0.020]**  -0.119 [-0.257; 0.020] | **p = 0.093**  p = 0.093 |
| 18 | Parahippocampal cortex |  |  |  |  |
| **SOM** | | SP | **2, 3, 178**  1, 1, 117  1, 2, 61 | **0.010 [-0.301; 0.321]**  0.288 [0.104; 0.471]  -0.111 [-0.293; 0.071] | **p = 0.951**  *p = 0.002*  p = 0.234 |
| 20 | Heschl’s gyrus |  |  |  |  |
| 23 | Superior temporal gyrus |  |  |  |  |
| **SOM** | | ATT | **1, 6, 51**  1, 2, 51  1, 4, 51 | **0.358 [0.205; 0.511]**  0.551 [0.351; 0.751]  0.365 [0.165; 0.566] | ***p < 0.001***  *p < 0.001*  *p < 0.001* |
| 20 | Heschl’s gyrus |  |  |  |  |
| 21 | Planum temporale |  |  |  |  |
| **SOM** | | WM | **2, 2, 157**  2, 2, 157 | **0.436 [0.276; 0.595]**  **0.436 [0.276; 0.595]** | ***p < 0.001***  *p < 0.001* |
| 23 | Superior temporal gyrus |  |  |  |  |
| **SOM** | | VM | **2, 3, 76**  2, 3, 76 | **0.147 [-0.127; 0.421]**  0.147 [-0.127; 0.421] | **p = 0.292**  p = 0.292 |
| 23 | Superior temporal gyrus |  |  |  |  |
| **SOM** | | VisM | **2, 3, 89**  2, 3, 89 | **0.358 [-0.271; 0.987]**  0.358 [-0.271; 0.987] | **p = 0.264**  p = 0.264 |
| 23 | Superior temporal gyrus |  |  |  |  |
| **SOM** | | R&EF | **6, 34, 310**  1, 1, 25  2, 6, 135  2, 3, 43  1, 12, 18  4, 12, 168 | **0.537 [0.384; 0.690]**  0.769 [0.351; 1.186]  0.636 [0.410; 0.863]  0.123 [-0.665; 0.911]  0.757 [0.611; 0.903]  0.455 [0.263; 0.647] | ***p < 0.001***  *p < 0.001*  *p < 0.001*  p = 0.759  *p < 0.001* |
| 19 | Claustrum |  |  |  |  |
| 20 | Heschl’s gyrus |  |  |  |  |
| 21 | Planum temporale |  |  |  |  |
| 22 | Precentral gyrus |  |  |  |  |
| 23 | Superior temporal gyrus |  |  |  |  |
| **SOM** | | SC | **2, 11, 60**  1, 3, 21  1, 2, 21  2, 6, 60 | **0.784 [0.545; 1.024]**  0.935 [0.669; 1.202]  0.963 [0.637; 1.290]  0.667 [0.292; 1.043] | ***p < 0.001***  *p < 0.001*  *p < 0.001*  *p < 0.001* |
| 22 | Precentral gyrus |  |  |  |  |
| 23 | Superior temporal gyrus |  |  |  |  |
| 24 | Supplementary motor area |  |  |  |  |
| **SOM** | | VF | **2, 4, 80**  2, 4, 80 | **0.130 [-0.209; 0.470]**  0.130 [-0.209; 0.470] | **p = 0.452**  p = 0.452 |
| 23 | Superior temporal gyrus |  |  |  |  |
| **VAN** | | SP | **1, 1, 32**  1, 1, 32 | **0.352 [-0.012; 0.716]**  0.352 [-0.012; 0.716] | **p = 0.058**  p = 0.058 |
| 26 | Insula |  |  |  |  |
| **VAN** | | WM | **2, 3, 49**  1, 1, 32  1, 2, 17 | **0.443 [0.183; 0.702]**  0.317 [-0.047; 0.681]  0.572 [0.202; 0.943] | ***p < 0.001***  p = 0.088  *p = 0.002* |
| 26 | Insula |  |  |  |  |
| 28 | Superior frontal gyrus |  |  |  |  |
| **VAN** | | VM | **3, 3, 117**  2, 2, 60  1, 1, 57 | **0.622 [0.253; 0.992]**  0.654 [-0.057; 1.366]  0.576 [0.310; 0.843] | ***p < 0.001***  p = 0.071  *p < 0.001* |
| 26 | Insula |  |  |  |  |
| 28 | Superior frontal gyrus |  |  |  |  |
| **VAN** | | VisM | **1, 2, 113**  1, 2, 113 | **-0.006 [-0.138; 0.126]**  -0.006 [-0.138; 0.126] | **p = 0.927**  p = 0.927 |
| 26 | Insula |  |  |  |  |
| **VAN** | | R&EF | **2, 13, 93**  1, 6, 18  3, 7, 142 | **0.604 [0.511; 0.697]**  0.775 [0.569; 0.982]  0.538 [0.046; 1.031] | ***p < 0.001***  *p < 0.001*  p = 0.032 |
| 25 | BANKSTS |  |  |  |  |
| 28 | Superior frontal gyrus |  |  |  |  |
| **VAN** | | SC | **4, 41, 193**  3, 29, 173  1, 3, 21  1, 7, 21  1, 2, 39 | **0.315 [0.214; 0.415]**  0.142 [0.060; 0.225]  1.131 [0.658; 1.605]  0.964 [0.790; 1.139]  0.090 [-0.141; 0.321] | ***p < 0.001***  *p < 0.001*  *p < 0.001*  *p < 0.001*  p = 0.443 |
| 26 | Insula |  |  |  |  |
| 27 | Middle cingulate gyrus |  |  |  |  |
| 28 | Superior frontal gyrus |  |  |  |  |
| 36 | Temporoparietal junction |  |  |  |  |
| **VAN** | | VF | **1, 1, 26**  1, 1, 26 | **0.151 [-0.258; 0.560]**  0.151 [-0.258; 0.560] | **p = 0.469**  p = 0.469 |
| 28 | Superior frontal gyrus |  |  |  |  |
| **VIS** | | SP | **1, 1, 32**  1, 1, 32 | **0.535 [0.171; 0.899]**  0.535 [0.171; 0.899] | **p = 0.004**  p = 0.004 |
| 29 | Calcarine cortex |  |  |  |  |
| **VIS** | | VM | **1, 1, 32**  1, 1, 32 | **-0.302 [-0.666; 0.062]**  -0.302 [-0.666; 0.062] | **p = 0.104**  p = 0.104 |
| 33 | Middle occipital gyrus |  |  |  |  |
| **VIS** | | VisM | **2, 12, 60**  2, 12, 60 | **0.291 [0.072; 0.509]**  0.291 [0.072; 0.509] | ***p = 0.009***  *p = 0.009* |
| 30 | Fusiform gyrus |  |  |  |  |
| **VIS** | | R&EF | **4, 24, 182**  4, 18, 182  1, 1, 25  1, 1, 25  1, 4, 18 | **0.592 [0.448; 0.734]**  0.490 [0.241; 0.738]  0.723 [0.306; 1.141]  0.803 [0.385; 1.221]  0.695 [0.189; 1.202] | ***p < 0.001***  *p < 0.001*  *p < 0.001*  *p < 0.001*  p = 0.014 |
| 30 | Fusiform gyrus |  |  |  |  |
| 32 | Lingual gyrus |  |  |  |  |
| 31 | Inferior occipital |  |  |  |  |
|  | Occipital Lobe |  |  |  |  |
| **VIS** | | SC | **1, 3, 21**  1, 3, 21 | **0.914 [0.648; 1.181]**  0.914 [0.648; 1.181] | ***p < 0.001***  *p < 0.001* |
| 33 | Middle occipital gyrus |  |  |  |  |

*Significant results* after FDR correction are in Figure 3A for **networks** and 3B for brain areas.

Both significant (color) and nonsignificant (grey) results are visualized in Figure S12A for **networks** and S12B for brain regions.

S: number of studies; C: number of correlations; N: sample size

### **Table S5.** Values of effects, corrected with FDR, observed between the eight cognitive domains and the three structures selected.

| **Structure** | **Cognitive Domain** | **S, C, N** | **Fisher’s z [95% CI] random model** | **p-value** |
| --- | --- | --- | --- | --- |
| Amygdala | SP | 1, 6, 163 | 0.204 [0.141; 0.267] | *p < 0.001* |
|  | ATT | 1, 2, 174 | 0.100 [-0.006; 0.206] | p = 0.065 |
|  | WM | 1, 8, 163 | 0.136 [0.081; 0.191] | *p < 0.001* |
|  | VM | 3, 6, 213 | 0.129 [-0.161; 0.419] | p = 0.384 |
|  | VisM | 3, 6, 213 | 0.307 [0.000; 0.614] | p = 0.050 |
|  | R&EF | 2, 9, 192 | 0.169 [0.075; 0.263] | *p < 0.001* |
|  | SC | 2, 4, 36 | 0.596 [0.351; 0.841] | *p < 0.001* |
| Hippocampus | SP | 7, 22, 346 | 0.130 [0.080; 0.180] | *p < 0.001* |
|  | ATT | 3, 14, 213 | -0.015 [-0.161; 0.131] | p = 0.841 |
|  | WM | 7, 48, 512 | 0.159 [0.115; 0.202] | *p < 0.001* |
|  | VM | 17, 75, 967 | 0.234 [0.181; 0.287] | *p < 0.001* |
|  | VisM | 11, 35, 653 | 0.141 [0.063; 0.219] | *p < 0.001* |
|  | R&EF | 8, 36, 396 | 0.190 [0.111; 0.268] | *p < 0.001* |
|  | SC | 1, 1, 21 | 1.263 [0.801; 1.725] | *p < 0.001* |
|  | VF | 3, 5, 128 | -0.134 [-0.276; 0.009] | p = 0.066 |
| Cerebellum | ATT | 1, 20, 40 | 0.109 [0.030; 0.188] | *p = 0.07* |
|  | WM | 1, 2, 28 | 0.800 [0.141; 1.078] | *p < 0.001* |
|  | VM | 2, 21, 55 | -0.004 [-0.112; 0.105] | p = 0.948 |
|  | VisM | 1, 3, 28 | 1.042 [0.816; 1.268] | *p < 0.001* |
|  | R&EF | 4, 37, 142 | 0.078 [-0.015; 0.170] | p = 0.099 |
|  | VF | 1, 30, 40 | 0.201 [0.135; 0.267] | *p < 0.001* |

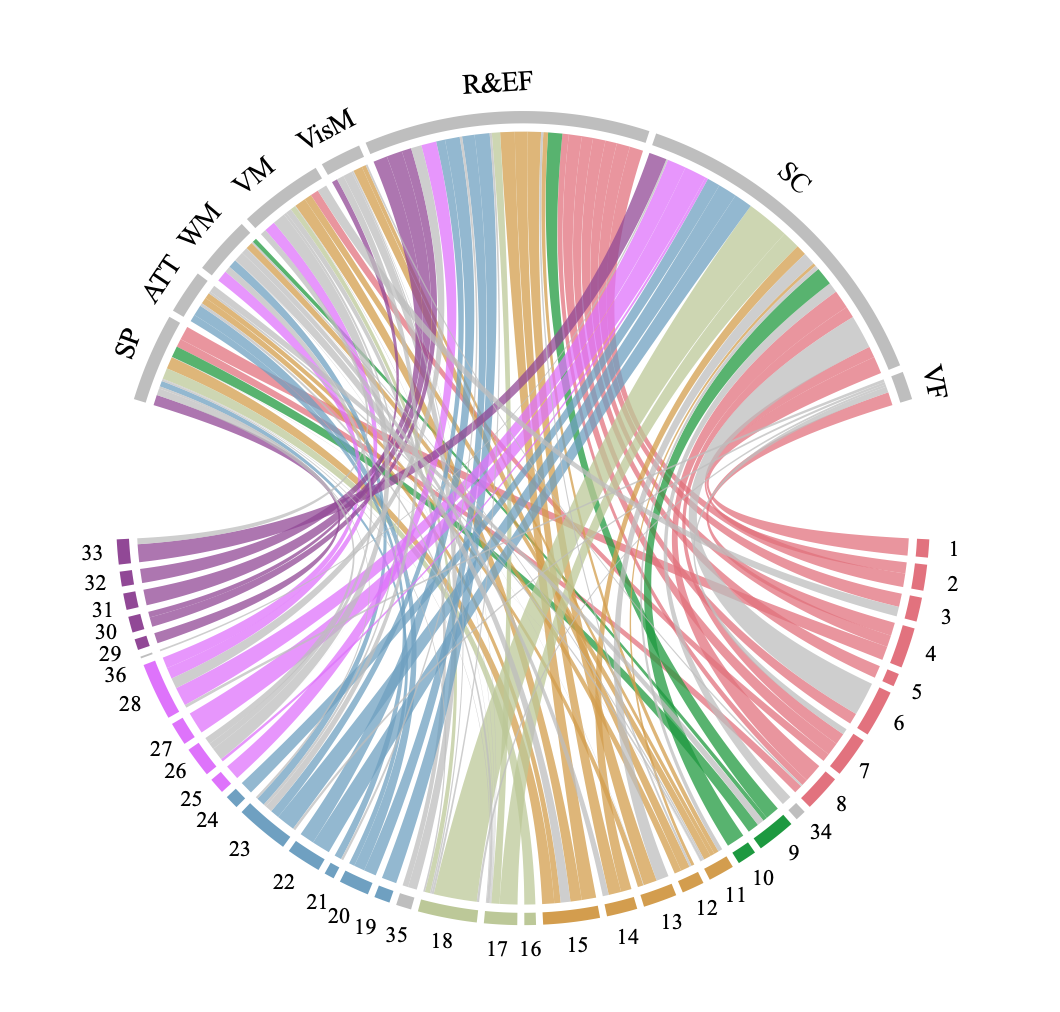

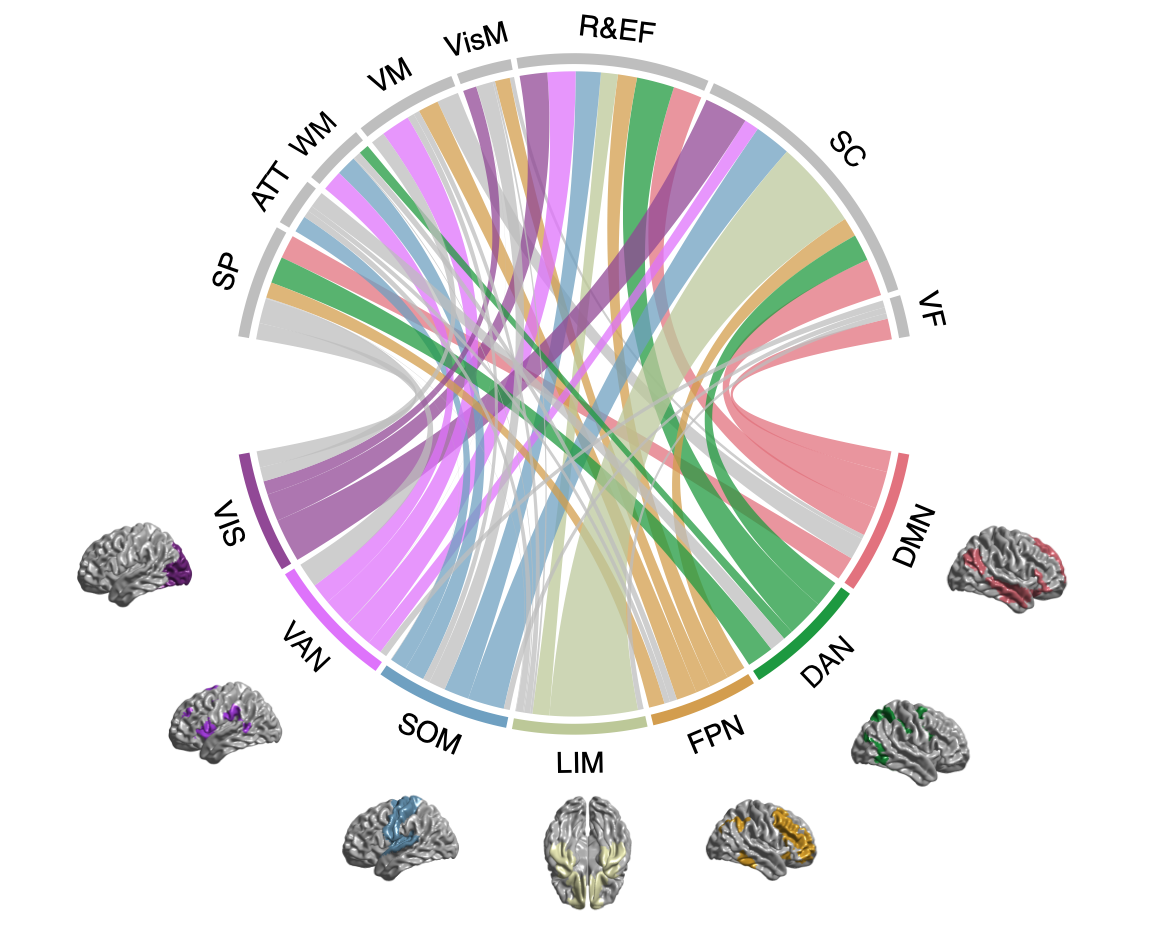

### **Figure S12.** A) Circle plot of the FDR significant correlations between the seven brain networks and the eight cognitive domains. B) Circle plot of the correlations between the brain regions categorized in the seven brain networks and the eight cognitive domains.

**B)**

**A)**

The thickness of the link is proportional to the correlation strength. The colors of the brain regions correspond to the respective network. The grey links are nonsignificant with FDR. The legend and the values of the effect are in Table S4.
